# Hematopoietic mosaic chromosomal alterations are pleiotropic drivers of inflammaging, multimorbidity, and mortality

**DOI:** 10.64898/2026.06.24.26356446

**Authors:** Nicole D. Vincelette, Qianxing Mo, Chia-Ho Cheng, Junyoung Park, Andrew T. Kuykendall, Ling Zhang, Jungwon Moon, Tiffany N. Razabdouski, Erika A. Eksioglu, Felyschia M. Lledo, Peter R. Papenhausen, Zhuoer Xie, Onyee Chan, David A. Sallman, Javier Pinilla-Ibarz, Daniel J. Murphy, Rami S. Komrokji, John L. Cleveland, Xiaoqing Yu, Seongseok Yun

**Author notes:** **Correspondence to:** Nicole D. Vincelette PhD, Malignant Hematology Department, H. Lee Moffitt Cancer Center and Research Institute, 12902 USF Magnolia Drive, Tampa, FL 33612, Seongseok Yun MD PhD, Malignant Hematology Department, H. Lee Moffitt Cancer Center and Research Institute, 12902 USF Magnolia Drive, Tampa, FL 33612. N.D.V. and S.Y. contributed equally as co-first authors and share corresponding authorship.

## Abstract

Mosaic chromosomal alterations (mCAs) are a prevalent but poorly understood form of clonal hematopoiesis (CH). Whether mCAs contribute to disease independently of CHIP, and whether their large-scale genomic effects can be resolved to actionable targets, remain unknown. In 452,594 UK Biobank participants, we show that mCAs confer multimorbidity and mortality risk independent of CHIP. Notably, mCA-CHIP co-occurrence defines a very high-risk clonal state with synergistically elevated mortality, identifying a population not captured by CHIP screening alone. To resolve large mCAs to specific disease mechanisms, a cytoband-level mapping framework was developed that links mCAs to discrete genomic loci and candidate effector genes. Functional validation using single-cell transcriptomics and mouse models prioritized MYC (chr8 gain) and S100A9 (chr1 gain) as key drivers of systemic inflammation and multiorgan pathology. These findings establish mCAs as independent, synergistic, and genetically-resolvable drivers of age-related disease, with immediate implications for screening, risk stratification, and therapeutic development.

**SIGNIFICANCE:** Hematopoietic mCAs drive age-related multimorbidity and mortality independently of CHIP, while co-occurrence defines a synergistically high-risk clonal state undetectable by standard screening. Integrative cytoband-level mapping and functional validation resolve large chromosomal alterations to discrete effector genes, enabling mechanistic risk stratification and informing precision surveillance and targeted therapeutic strategies.

## INTRODUCTION

Clonal hematopoiesis (CH) is an age-related expansion of hematopoietic stem cells (HSCs) and progenitors driven by somatic genetic alterations. Clonal hematopoiesis of indeterminate potential (CHIP), the most extensively studied form of CH, is defined by somatic mutations in myeloid malignancy-associated genes—most commonly *DNMT3A, ASXL1*, and *TET2*—at variant allele fractions (VAF) ≥2% without overt hematologic disorders^1–4^. CHIP prevalence increases markedly with age and confers risk for hematologic malignancies and multiple age-related diseases through inflammation-driven mechanisms^2–17^.

Mosaic chromosomal alterations (mCAs), including gains, losses, and copy number neutral loss of heterozygosity (CNLOH), represent a distinct yet comparatively understudied form of CH that also affects 10-40% of individuals over the age of 70^18–21^. Like CHIP, mCA prevalence increases with age and is associated with hematologic malignancy risk^18–21^. However, their broader clinical impact beyond hematologic malignancies remains poorly defined. Loss of Y chromosome, one of the most studied mCAs, has been linked to solid tumors, Alzheimer’s disease, and cardiovascular disease (CVD)^22–25^, but the effects of mCAs affecting other chromosomes on disease incidence, multimorbidity, and mortality remain poorly characterized. A major challenge lies in mapping large-scale chromosomal alterations, which can span hundreds of genes, to specific disease phenotypes and in identifying key cytogenetic loci and effector genes that drive systemic disease risk.

Here, we systematically characterized associations between mCAs and disease incidence and mortality through genomic and epidemiological analyses in 452,594 UK Biobank (UKBB) participants combined with single-cell RNA-seq (scRNA-seq) of mCA-positive bone marrow (BM) mononuclear cells from patients with myeloid diseases (**Supplementary Fig. 1**). These analyses established dose-response relationships between mosaic cell fraction (MCF) and disease risk. We applied regularized regression to identify disease-associated cytogenetic loci. Focusing on chr8 and chr1 gains as representative mCAs with broad disease associations, we validated computational predictions via scRNA-seq of patient samples and provided evidence for causality using genetically engineered mouse models. Collectively, these findings establish mCAs as pleiotropic drivers of age-related multimorbidity and provide a platform for translating large-scale structural mosaicism into actionable biological insight.

## RESULTS

### Population landscape and baseline associations of mosaic chromosomal alterations

To characterize the population-level landscape of mCAs in hematopoiesis, we leveraged mCA calls from the SNP array-based analysis of UKBB participants^20^. Among 452,594 participants, 254,879 (56.3%) were female, with median age of 57 years (IQR 50-63) at recruitment and 58 years (IQR 50-63) at mCA assessment (**Supplementary Fig. 2a-c** and **Table S1**). A total of 42,390 participants (9.4%) harbored ≥1 mCA (**Fig. 1a**), comprising 45,536 events: 2,541 (5.6%) gains (+), 34,686 (76.2%) losses (-), and 8,309 (18.2%) CNLOH (=) (**Supplementary Fig. 2d**). Most carriers harbored a single mCA, with a subset harboring multiple events (range: 2-22) (**Supplementary Fig. 2e**). The most commonly affected chromosomes were Y, X, 1, 11, 14, and 13, with frequent mCA including Y-, X-, 1=, 11=, 9=, and 14= (**Supplementary Fig. 2f**). Notably, mCA prevalence increased with age, with heterogeneous expansion kinetics and sex-specific variations (**Fig. 1b-e**). The MCFs were generally higher for gains and losses than for CNLOH and varied substantially across mCA types, consistent with mCA-specific fitness effects^26,27^ (**Supplementary Fig. 2g-j**).

**Fig. 1.**
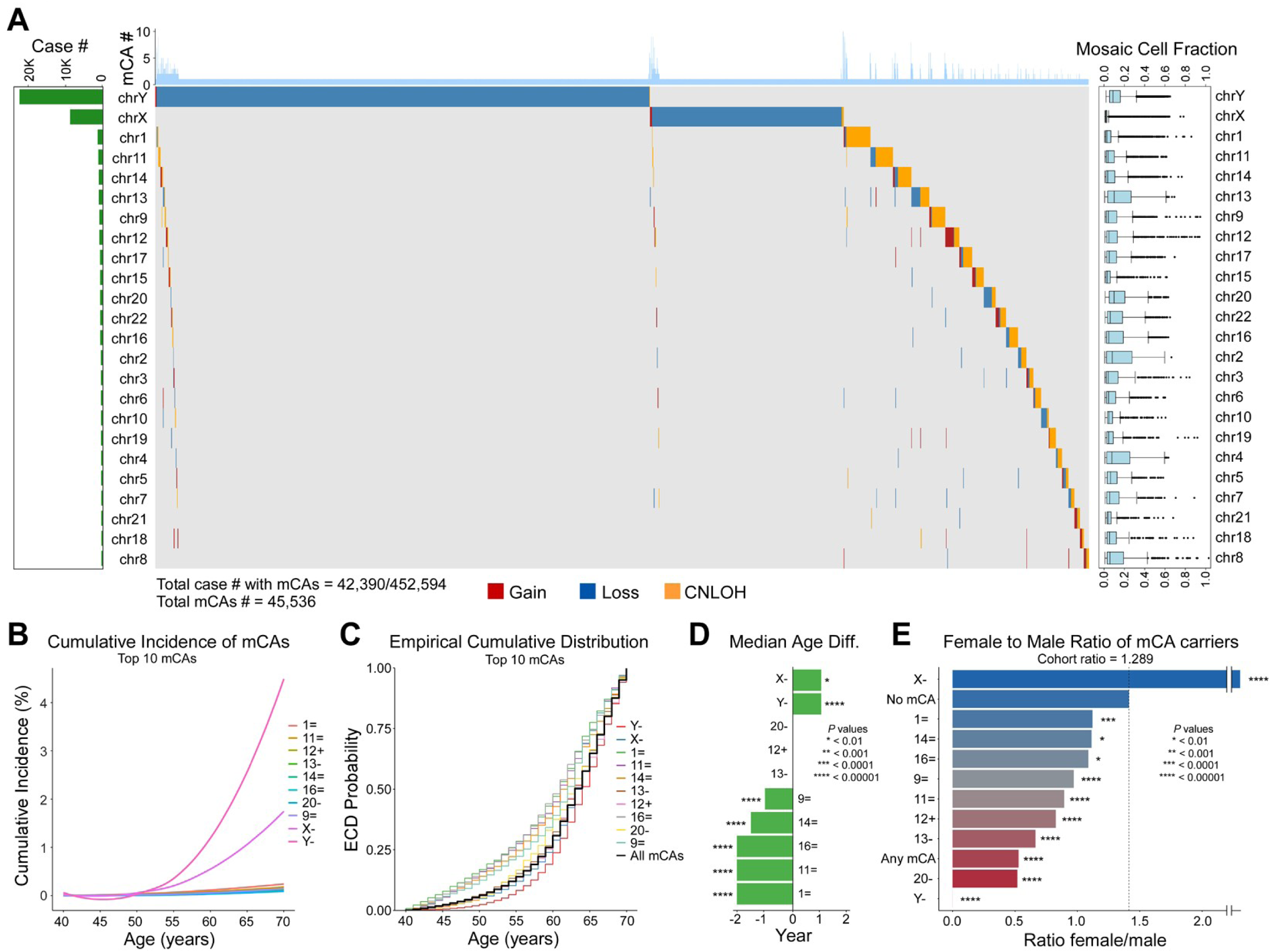
Landscape and age-related dynamics of mCAs in the UK Biobank. **a**, Distribution of mCAs across 452,594 participants (mCA, n=42,390; no mCA, n=410,204). Left, number of carriers per chromosome (horizontal bars). Centre, individual mCA events displayed as an oncoplot; each column represents a participant and rows represent chromosomes (red, gain; blue, loss; orange, CNLOH). Top histogram shows mCA count per individual. Right, mosaic cell fraction (MCF) distribution per chromosome (box plots show median and interquartile range). **b**, Cumulative incidence of the ten most frequent mCAs by age. **c**, Empirical cumulative distribution of age at mCA detection for the top ten subtypes. **d**, Median age difference (years) between each mCA subtype and all mCA carriers combined. P values were calculated using the Wilcoxon rank-sum test. **e**, Female-to-male ratio among mCA subtypes, all mCA carriers combined, and non-carriers (control). P values were calculated using the chi-square test comparing each group’s sex distribution to the non-carrier control group. Dashed line indicates female-to-male ratio in the non-carrier control group (1.413). The overall cohort female-to-male ratio was 1.289.

Cross-sectional logistic regression analysis revealed associations between mCAs and older age (OR=1.12, p<0.0001), male sex (OR=2.67, p<0.0001), and smoking or alcohol use; BMI and obesity showed no association (**Supplementary Fig. 3a**). In addition, adjusted CBC analyses revealed positive associations with white blood cell (WBC) count (OR=2.25, p<0.0001), consistent with findings in CHIP^12^, and negative associations with red blood cell count (OR=0.35, p<0.0001), while platelet associations were heterogeneous (**Supplementary Fig. 3b**). Biochemical profiling showed reduced urate and hemoglobin A1c (HbA1c) levels among mCA carriers, whereas pro-inflammatory cytokines and alarmins were elevated (**Supplementary Fig. 3c-f**). Collectively, these findings establish mCAs as common, clonally heterogeneous features of aging hematopoiesis, which are associated with pro-inflammatory profiles that may contribute to systemic disease.

### mCAs confer broad risk of multimorbidity

Given the positive association of mCAs with elevated WBC count, pro-inflammatory cytokines, and alarmins, we performed a phenome-wide analysis of incident disease risk using baseline mCA status, excluding pre-existing conditions prior to mCA assessment and accounting for death as a competing event. At baseline, mCA carriers had slightly more pre-existing conditions than non-carriers (3.37 vs. 2.87, p<0.001) (**Fig. 2a** and **Supplementary Fig. 4a-b**). mCA carriers had significantly higher cumulative incidence of new disease (p<0.0001), developed more incident diseases (HR=1.33, p<0.0001), and experienced more frequent hospital admissions (HR=1.39, p<0.0001) (**Fig. 2b-c**), indicating an accelerated medical burden with aging.

**Fig. 2.**
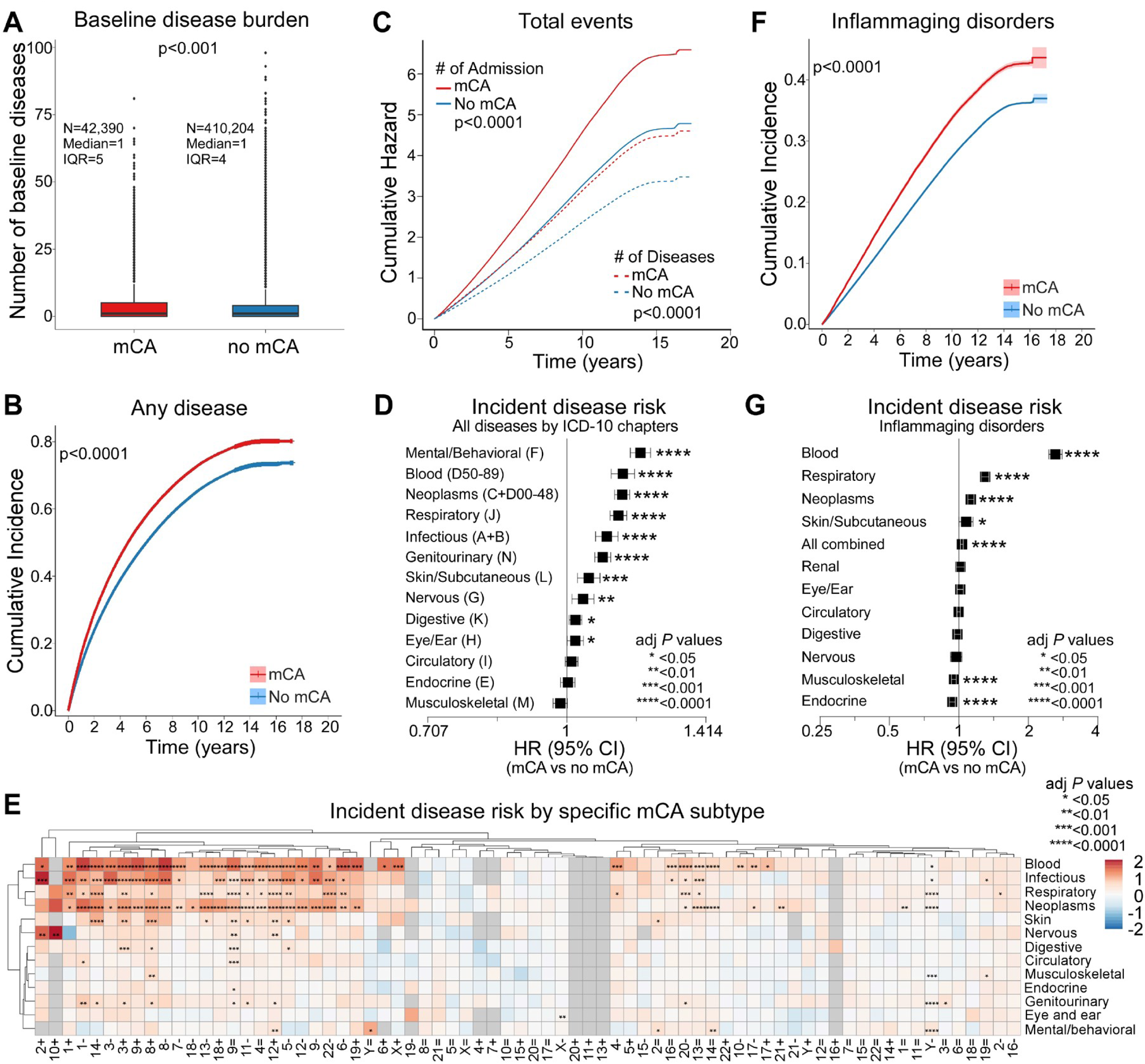
Association between mCAs and incident disease risk. **a**, Baseline disease burden at mCA assessment. Box plots show median and interquartile range (IQR) of total number of diseases diagnosed prior to mCA assessment. P value was calculated using Wilcoxon rank-sum test. **b**, Cumulative incidence of any new disease diagnosis comparing mCA carriers (red) vs. non-carriers (blue). **c**, Cumulative hazard of total hospital admissions (solid lines) and total incident diseases (dashed lines) over time comparing mCA carriers (red) vs. non-carriers (blue). **d**, Subdistribution HRs with 95% CIs for incident diseases across ICD-10 chapters, adjusted for age, sex, and baseline disease burden. **e**, Heatmap of incident disease risk by specific mCA subtype across ICD-10 chapters. Color scale represents log₂-transformed HRs; red indicates increased risk and blue indicates decreased risk. Hierarchical clustering was applied to rows and columns using complete linkage with Euclidean distance. **f**, Cumulative incidence of inflammaging-related disorders comparing mCA carriers (red) vs. non-carriers (blue). **g**, Adjusted HRs with 95% CIs for inflammaging-related disorders by organ system. For **b** and **f**, P values were calculated using Gray’s test. For **d**, **e**, and **g**, Fine-Gray competing risk regression was used with death as a competing event, and multiple testing was corrected using the Benjamini-Hochberg method.

When stratified by ICD-10 category, mCA carriers showed significantly increased incidence risk across nearly all disease chapters (**Supplementary Fig. 4c-o**). Fine-Gray competing risk regression analysis confirmed independent associations between mCAs and incident diseases (HRs=1.02-1.20, all p<0.05) (**Fig. 2d** and **Table S2**). Further, consistent with prior reports^19,20^, analysis of individual mCA subtypes revealed that several alterations are significantly associated with hematologic disorders (**Fig. 2e, Supplementary Fig. 5a-m**, and **Table S3**). Notably, 3+, 8+, 9=, 12+, and Y- exhibit the broadest spectrum of associations, conferring increased risk across multiple ICD-10 chapters. Thus, specific mCAs confer broad susceptibility to multimorbidity, with effects that extend well beyond hematologic disease.

### mCAs associate with inflammaging-related diseases

The broad, multi-system disease associations observed for mCAs, along with elevated pro-inflammatory cytokines and alarmins, suggested shared inflammatory pathways^2,5,28,29^, prompting evaluation of inflammaging-related disorders. Notably, mCA carriers had higher cumulative incidence of inflammaging-related conditions (p<0.0001) (**Fig. 2f**). Further, after adjusting for baseline health, mCAs were associated with increased risk of hematologic, respiratory, neoplastic, skin and subcutaneous disorders (HRs=1.07-2.62, all p<0.05) (**Fig. 2g** and **Table S4**). In contrast, musculoskeletal (HR=0.95) and endocrine/metabolic disorders (HR=0.94) were reduced in mCA carriers (both p<0.0001), and this was primarily driven by lower incidence of gout and type 2 diabetes among Y- carriers (**Fig. 2g** and **Supplementary Fig. 6a-b**), consistent with decreased urate and HbA1c (**Supplementary Fig. 3c**). Finally, individual mCAs exhibited heterogeneous associations across organ systems, including cardiovascular, respiratory, and digestive disorders (**Supplementary Fig. 6a-l**, and **Table S5**). Collectively, these findings support a role for mCAs in systemic inflammation driving age-related multimorbidity.

### Disease risk scales with mCA clonal burden

Prior analyses have treated mCA status as binary, despite mCAs spanning a spectrum of clonal burden (**Supplementary Fig. 2g-j**). Analogous to the dose-response relationship between VAF and disease risk for somatic mutations^2,5,12,30^, we tested if higher MCF confers proportionally greater risk. This was evaluated using complementary binning and spline regression models adjusted for key covariates.

For 9=, 5%-binning revealed a clear dose-response gradient for hematologic disorders, confirmed by spline regression analysis (**Fig. 3a-b** and **Table S6**). Similar patterns were observed for 12+, 13-, and Y-, with MCF-dependent increases in hematologic disease risk across both modeling approaches (**Fig. 3c-d** and **Supplementary Fig. 7a-d**). These graded associations extended across multiple organ systems (**Supplementary Fig. 7e-l**). Thus, there are dose-dependent relationships between mCAs and multimorbidity.

**Fig. 3.**
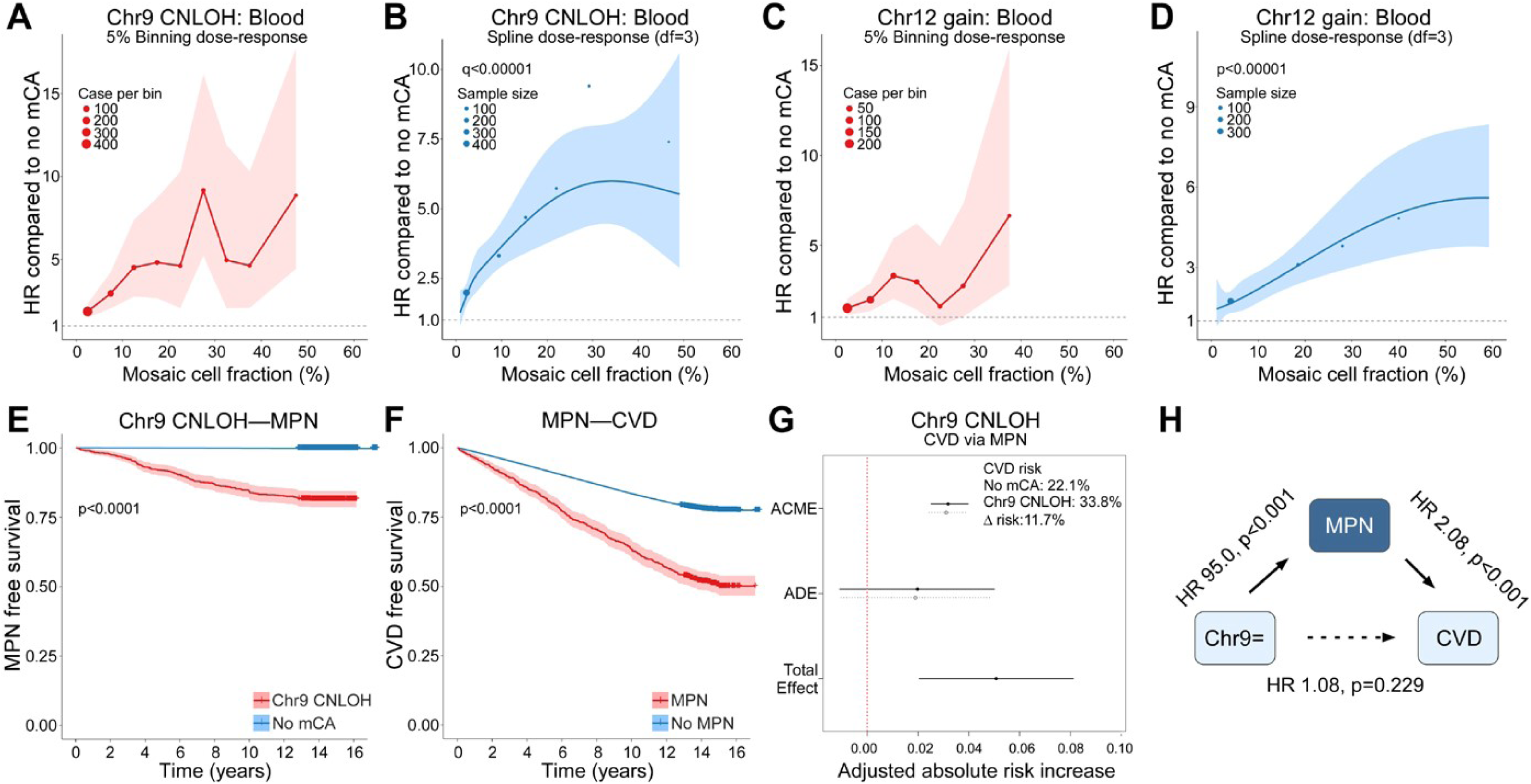
Dose-response relationships between mosaic cell fraction and disease risk, and mediation analyses. **a-b**, Dose-response relationship between chromosome 9 CNLOH (9=) mosaic cell fraction (MCF) and hematologic disease risk using 5% binning (**a**) and natural cubic spline regression (df=3) (**b**). **c-d**, Dose-response relationship between chromosome 12 gain (12+) MCF and hematologic disease risk using 5% binning (**c**) and natural cubic spline regression (df=3) (**d**). In **a** and **c**, dot size reflects sample size per bin, and shaded areas represent 95% CIs; in **b** and **d**, shaded areas represent 95% CIs. Dashed horizontal lines indicate HR=1. **e-h**, Mediation analysis of 9= on CVD risk via myeloproliferative neoplasm (MPN). MPN-free survival comparing 9= carriers vs. non-carriers (**e**) and CVD-free survival comparing individuals with vs. without MPN (**f**); shaded areas represent 95% CIs and P values were calculated using log-rank test. Causal mediation analysis showing average causal mediation effect (ACME), average direct effect (ADE), and total effect with 95% CIs (**g**); mediation models were adjusted for age, sex, and baseline disease burden. Path diagram summarizing statistically inferred direct and mediated effects (**h**); HRs were estimated using Cox proportional hazards regression adjusted for age, sex, and baseline disease burden; solid arrows indicate significant associations, and dashed arrow indicates non-significant direct effect.

### Hematologic-dependent and -independent pathways link mCAs to systemic disease

Given dose-dependent associations of mCAs with both hematologic and non-hematologic diseases, we investigated if effects on non-hematologic diseases were direct or mediated via hematologic malignancies. We focused on mCA-disease pairs with known hematologic intermediates: myeloproliferative neoplasms (MPN) as a mediator between 9= and CVD, and chronic lymphocytic leukemia (CLL) as a mediator between 12+ or 13- and chronic bronchitis or actinic keratosis^31–34^.

For 9= and CVD, absolute risk increased from 22.1% in individuals without mCAs to 33.8% in 9= carriers (Δ=11.7%) (**Fig. 3e-h**). Similarly, 13- increased actinic keratosis risk from 0.95% to 3.53% (Δ=2.58%) (**Supplementary Fig. 7m-p**). Both mCAs strongly predicted their respective hematologic malignancies (9=→MPN: HR=95.0, 13-→CLL: HR=206.2, both p<0.001), which in turn predicted the outcomes (MPN→CVD: HR=2.08; CLL→actinic keratosis: HR=3.12, both p<0.001). Notably, direct effects were not significant (9=→CVD: HR=1.08, p=0.229; 13-→actinic keratosis: HR=1.48, p=0.115), where mediation analysis estimated that 64% of the 9= effect and 43% of the 13- effect were mediated via MPN and CLL, respectively (both ACME p<0.001). In contrast, 12+ exhibited both direct and mediated effects on chronic bronchitis, with risk rising from 3.85% to 8.68% (Δ=4.83%) (**Supplementary Fig. 7q-t**). 12+ strongly predicted CLL (HR=117.6, p<0.001), which in turn predicted chronic bronchitis (HR=1.45, p=0.017), yet after adjusting for CLL, the direct effect of 12+ remained significant (HR=1.39, p=0.028); thus 12+ influences chronic bronchitis risk through both hematologic and non-hematologic pathways. Therefore, mCAs contribute to multimorbidity through diverse mechanisms, from predominantly mediated to mixed direct and indirect effects.

### mCAs are associated with increased systemic mortality risk

Given the pleiotropic disease associations, effects of mCAs on mortality were assessed. Overall survival (OS) was shorter among mCA carriers (p<0.0001) (**Fig. 4a**) and mCAs were independently associated with higher all-cause mortality (HR=1.21, p<0.0001) (**Fig. 4b** and **Table S7**). Further, risk was highest for complex mCA (HR=3.11), followed by gain-only (HR=1.44), CNLOH-only (HR=1.30), and loss-only (HR=1.16) mCAs (all p<0.0001). High MCF conferred greater risk than low MCF (HR=1.40 vs. 1.10, both p<0.0001), supporting dose-response relationships for both mosaic complexity and clonal burden.

**Fig. 4.**
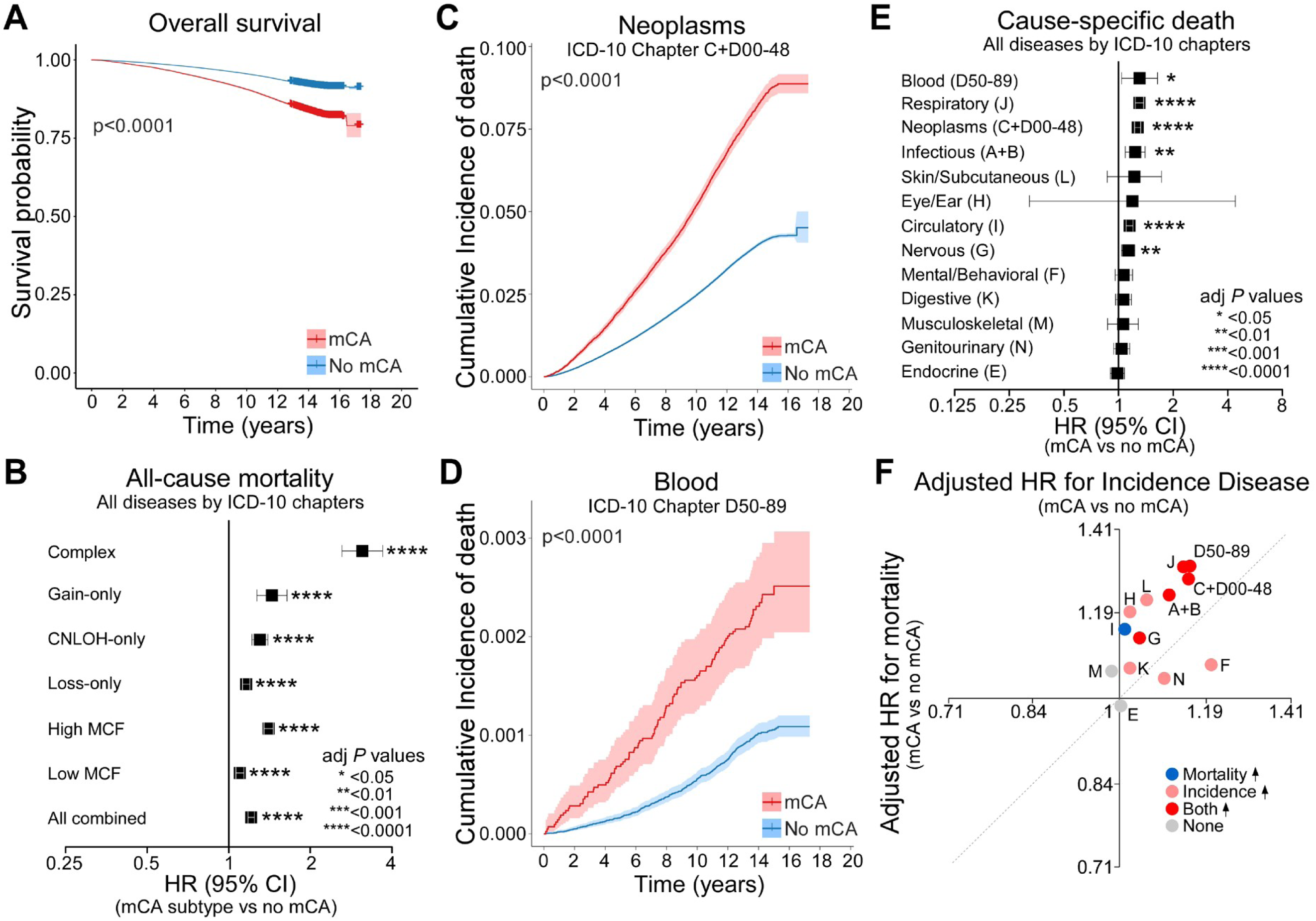
Mortality risk associated with mCAs and mCA subtypes. **a**, Overall survival comparing mCA carriers (red) vs. non-carriers (blue); P value calculated using log-rank test. **b**, All-cause mortality HRs with 95% CIs by mCA subtype using Cox proportional hazards regression, adjusted for age, sex, and baseline disease burden. mCA subtypes include: any mCA, complex mCA (≥3 chromosomes affected), low MCF (<10%), high MCF (≥10%), and by copy number change type (gain-only, loss-only, CNLOH-only). **c-d**, Cumulative incidence of death from neoplasms (ICD-10 chapters C+D00-48) (**c**) and hematologic disorders (ICD-10 chapter D50-89) (**d**) comparing mCA carriers (red) vs. non-carriers (blue); shaded areas represent 95% CIs and P values were calculated using Gray’s test. **e**, Cause-specific mortality HRs with 95% CIs across ICD-10 chapters using Fine-Gray competing risk regression with deaths from other causes treated as competing events. **f**, Comparison of mCA-associated effect sizes on disease incidence (x-axis) vs. mortality (y-axis) across ICD-10 chapters; each point represents one ICD-10 chapter (labeled by chapter code). Colors indicate increased mortality only (blue), increased incidence only (pink), both increased (red), or neither (gray). The dashed line denotes equal effect size for incidence and mortality. Visualization highlights heterogeneity in how mCAs relate to disease onset vs. survival. Multiple testing was corrected using the Benjamini-Hochberg method.

Cause-specific analyses revealed higher mortality among mCA carriers across multiple organ systems (**Fig. 4c-e, Supplementary Fig. 8a-k** and **Table S8**). Importantly, mCAs differentially contributed to disease incidence vs. mortality: many disease categories showed increases in both, whereas CVD showed increased mortality without change in incidence, and mental/behavioral, digestive, and other disorders showed the opposite pattern (**Fig. 4f**). Thus, mCAs exert heterogeneous effects on disease onset and survival across organ systems.

### mCAs and CHIP define a high-risk clonal state with synergistic effects on mortality

Beyond organ-specific heterogeneity, mCA carriers also have higher rates of somatic mutations (**Fig. 5a-d**), raising the possibility that some of the observed effects could be attributable to CHIP. To assess if mCAs confer disease and mortality risk independently, we restricted analysis to 404,468 participants without CHIP or missing mutation data (36,725 mCA+; 367,743 mCA-). Most disease associations remained significant (**Fig. 5e-f** and **Table S9**), and indeed mCA carriers had worse OS (HR=1.17, p<0.0001) with subtype-specific patterns consistent with the primary analyses (**Fig. 5g-h** and **Table S10**). Thus, mCA effects are largely CHIP-independent.

**Fig. 5.**
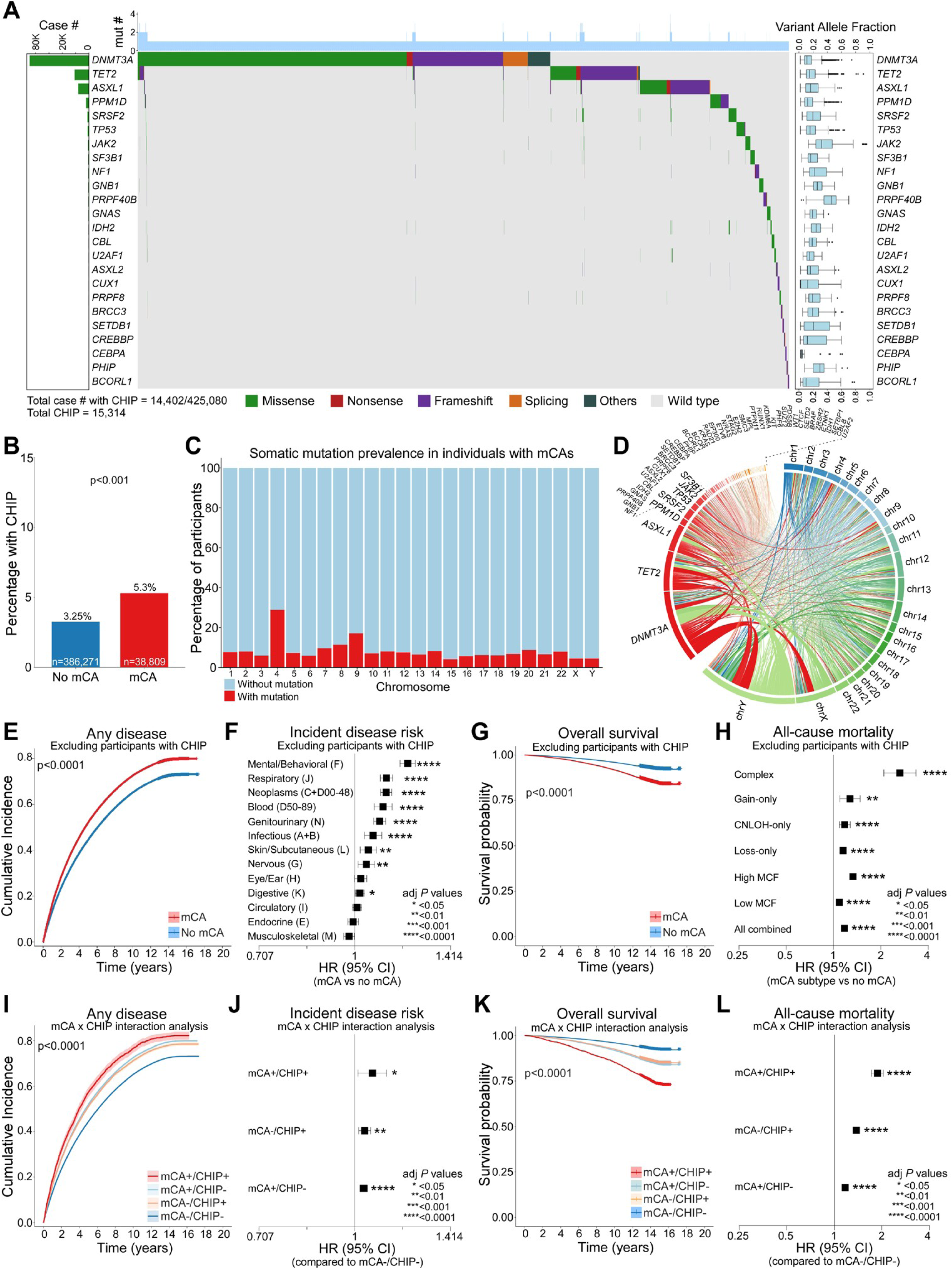
Independent and synergistic effects of mCAs and CHIP on disease risk and mortality. **a**, Landscape of somatic mutations in the UK Biobank cohort (top 24 most frequently mutated genes). Left, number of cases per gene. Centre, oncoplot showing mutation types (green, missense; red, nonsense; purple, frameshift; orange, splicing; dark green, others; gray, wild type). Right, variant allele fraction (VAF) distribution per gene. **b**, Prevalence of somatic mutations comparing mCA carriers (n=38,809) vs. non-carriers (n=386,271); 425,080 participants were assessed for both mCA and somatic mutations, with 15,314 somatic mutations identified in 14,402 participants; P value calculated using chi-square test. **c**, Prevalence of somatic mutations by chromosome among mCA carriers. **d**, Circos plot showing co-occurrence of somatic mutations and mCA-affected chromosomes. **e-h**, Sensitivity analyses excluding participants with CHIP: cumulative incidence of any disease (**e**), incident disease risk by ICD-10 chapter (**f**), overall survival (**g**), and all-cause mortality by mCA subtype (**h**). **i-l**, Joint effect and interaction analysis for mCA and CHIP: cumulative incidence of any disease (**i**), subdistribution HRs for incident disease risk compared to mCA-/CHIP- reference group (**j**), overall survival (**k**), and HRs for all-cause mortality compared to mCA-/CHIP- reference group (**l**). For incident disease, interaction was evaluated on both multiplicative and additive scales (see Methods). For **e** and **i**, P values were calculated using Gray’s test. For **g** and **k**, P values were calculated using log-rank test. For **f** and **j**, Fine-Gray competing risk regression was used with death as a competing event. For **h** and **l**, Cox proportional hazards regression was used. All regression models were adjusted for age, sex, and baseline disease burden; multiple testing was corrected using the Benjamini-Hochberg method.

To examine joint effects of mCA and CHIP, participants were stratified into four groups by mCA and CHIP status. For disease incidence, all exposed groups showed elevated risk compared to double-negative individuals (HRs=1.03-1.07, all p<0.05) (**Fig. 5i-l**), but no interaction was detected on either the multiplicative (interaction HR=0.996, p=0.88) or additive scale (RERI=-0.004, p=0.91; S=0.95, p=0.91), indicating independent contributions to disease incidence. In contrast, for mortality, mCA+/CHIP- (HR=1.17), mCA-/CHIP+ (HR=1.38), and mCA+/CHIP+ (HR=1.87) all showed elevated risk (all p<0.0001), with a significant positive interaction (interaction HR=1.16, p=0.0045). Thus, while mCAs and CHIP independently promote disease development, their co-occurrence synergistically compromises OS.

### Cytogenetic mapping resolves mCAs to disease-associated loci and candidate genes

mCAs can span hundreds of genes, complicating the localization of specific genomic regions underlying associations with multimorbidity and mortality. To assess if discrete sub-chromosomal regions are preferentially associated with disease risk, we developed a cytoband-level mapping approach to localize disease-relevant loci. mCAs were classified as homogeneous or heterogeneous according to predefined criteria (see Methods). LASSO-penalized Cox regression was then applied to prioritize disease-associated cytobands within heterogeneous mCAs (**Supplementary Fig. 9a-d**), followed by refitting selected loci in standard Cox models to estimate HRs and assess statistical significance. To evaluate the validity of this approach, bootstrap stability analysis was performed with 1,000 iterations. LASSO coefficients showed strong concordance with Cox HRs (r=0.561, 94% direction agreement), supporting the robustness of the variable selection pipeline (**Supplementary Fig. 10a-d**). Homogeneous mCA events (12+, 14+, 15+, 18+, and X-) were excluded from these analyses given limited resolution for whole-chromosome events with uniform genomic boundaries.

These analyses identified key cytogenetic loci associated with increased risk of hematologic and neoplastic diseases, including gains at 1q21-23, 3q11-26, 8q21-24 and 9p24; losses at 5q13-33, 7q21-36, 13q13-14, 17p11-13, and 20q13; and CNLOH at 1p34-36, 4q21-31, 9p21- 24, and 13q12-22 (**Fig. 6a-b**, **Supplementary Fig. 11-12**, and **Table S11**). Many of these loci correspond to established cytogenetic lesions in hematologic malignancies^1^, validating our mapping approach. Notably, several loci were also associated with non-neoplastic, non-hematologic diseases; for example, gains at 1q21.1-23.2 and 8q21.3-24.23, as well as CNLOH at 9p21.3-24.2, were linked to cardiovascular, infectious and digestive disorders.

**Fig. 6.**
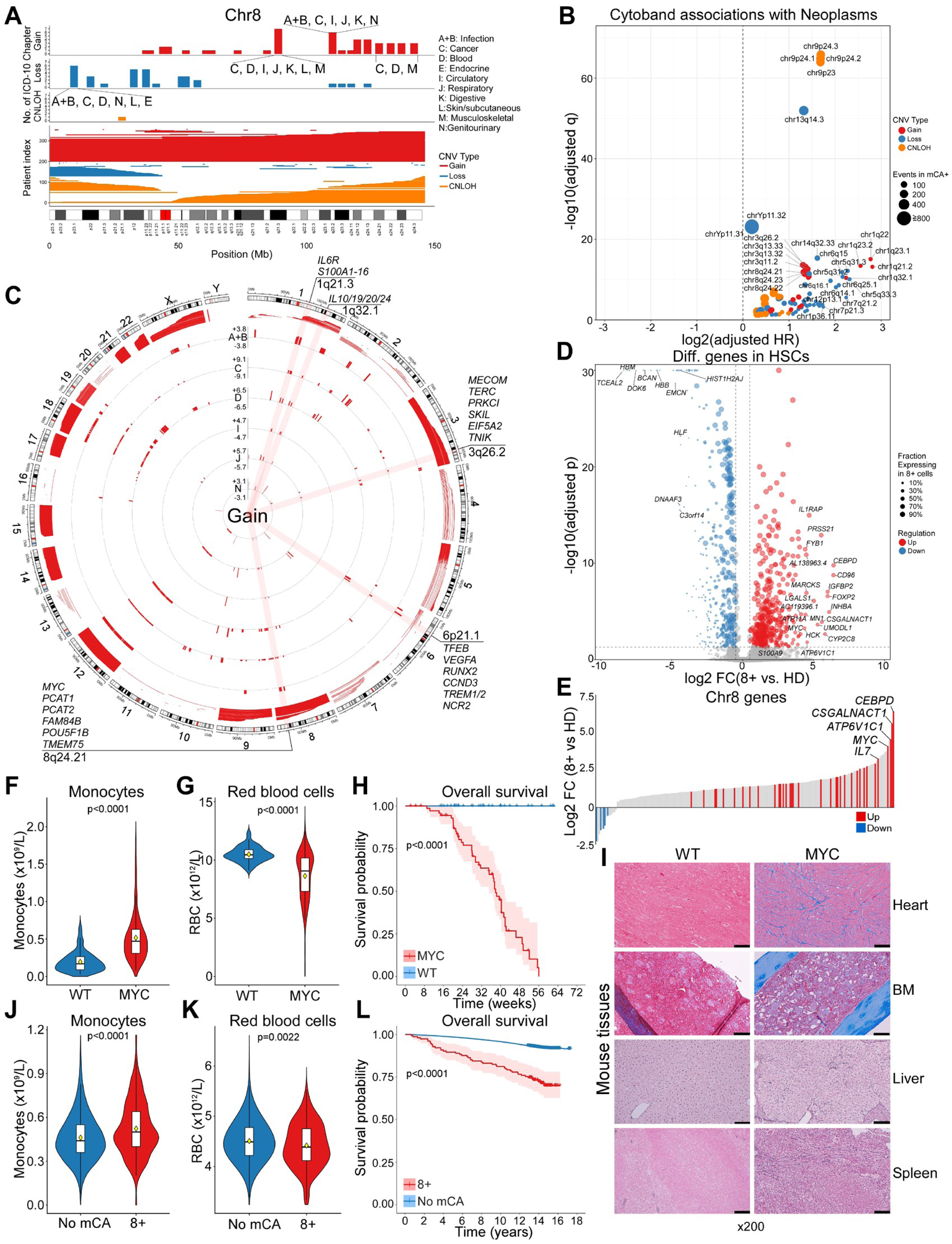
Cytoband-level mapping of disease-associated loci and functional validation of candidate gene in chr8 gain. **a**, Chr8 cytoband-level disease associations. Cytobands were selected by LASSO-penalized Cox regression and refitted using Cox models (see Methods). Top tracks show cytobands significantly associated with ICD-10 disease chapters (q<0.05) for gain, loss, and CNLOH. Bottom track displays mCA distribution across chr8; colors indicate CNV type (red, gain; blue, loss; orange, CNLOH). **b**, Volcano plot of cytoband associations with neoplasms. The x-axis shows log2-transformed HRs, and the y-axis shows -log10(q-values). Dot size reflects number of incident disease events among mCA carriers; colors indicate CNV type. **c**, Circos plot of genome-wide gain-associated cytobands. Outer track shows chromosomal ideogram; inner tracks display log₂-transformed HRs for cytobands significantly associated with incident disease risk (q<0.05). Selected candidate genes within these loci are annotated. **d**, Differentially expressed genes in HSCs from patients with 8+ compared to healthy donors (HD). scRNA-seq was performed on 71,090 cells from 13 samples (6 patients); 8+ (n=24,942) and disomy 8 (n=46,148) cells were classified using Numbat (see Methods). HD included 16,504 cells from 3 donors. Dot size reflects fraction of 8+ cells expressing each gene; colors indicate upregulation (red) or downregulation (blue). **e**, Chr8 genes ranked by log₂ fold change in 8+ HSCs vs. HD. Colors indicate upregulation (red, q<0.05) or downregulation (blue, q<0.05); gray bars represent nominally significant genes (p<0.05, q≥0.05). **f–h**, Monocyte counts (**f**), RBC counts (**g**), and overall survival (**h**) in Mx1-Cre^+/-^;Rosa26^LSL-MYC/LSL-MYC^ mice (MYC; n=77) vs. wild-type (WT; n=62) following pIpC induction. *MYC* expression levels are comparable to those in 8+ human HSCs (panel **e** and **Supplementary Fig. 17e**). **i**, Trichrome staining of the heart and BM, and reticulin staining of the liver and spleen from MYC overexpressing vs. WT mice (scale bar, 50 μm). **j–l**, Monocyte counts (**j**), RBC counts (**k**), and overall survival (**l**) in UKBB participants with 8+ (n=154) vs. non-carriers (n=410,204). For **f**, **g**, **j**, and **k**, P values were calculated using the Wilcoxon rank-sum test; box plots show median and interquartile range. For **h** and **l**, P values were calculated using log-rank test; shaded areas represent 95% CIs.

At the gene level, gain-associated loci were enriched for oncogenes and inflammatory mediators, including *S100A1-16, IL6R,* and *MCL1* (1q21-32); *MECOM* (3q26); *TFEB* (6q21); *MYC, GSDMC,* and *GSDMD* (8q21-24); and *JAK2* (9p24) (**Fig. 6c, Supplementary Fig. 13a**, and **Table S12**). Loss-associated regions harbor tumor suppressor genes and negative regulators of inflammation, including *RPS14* (5q33); *EZH2* (7q36); *DUSP4* and *DUSP26* (8p11-12); *CDKN2A/B* (9p21); *BRCA2, RB1,* and *miR-15a/16-1* (13q12-14); and *TP53* (17p13) (**Supplementary Fig. 13b**). CNLOH-associated cytobands linked to increased disease risk also harbored numerous oncogenes, tumor suppressors, and pro- and anti-inflammatory genes (**Supplementary Fig. 13c**). Thus, disease-associated mCAs localize to cytogenetic regions enriched for genes involved in oncogenesis and inflammatory signaling.

### Single-cell transcriptomics prioritizes candidate effector genes within mCA loci

To move from association towards mechanism, single-cell transcriptomics was used to prioritize candidate genes within disease-associated loci. We focused on 8+ as a representative example, given its (i) established clinical relevance in hematologic malignancies and demonstrated role in alarmin-mediated inflammation^35^, (ii) broad phenotypic associations across multiple disease categories (**Fig. 2e** and **6a**), and (iii) relatively high prevalence in hematologic malignancies, facilitating sample acquisition.

We performed scRNA-seq on BM mononuclear cells from patients with 8+ (MPN, MDS, MDS/MPN or AML; confirmed by karyotyping) and healthy donors (**Supplementary Fig. 14-15**). 8+ cells were distinguished from disomy 8 cells using Numbat^36^, and gene expression in 8+ cells was compared with that in healthy donor cells within each cluster. InferCNV^37^ was also used to identify chromosomal segments with altered transcriptional activity relative to healthy donor cells. Given the rarity of 8+ in the general population, obtaining CH-stage BM samples is impractical. Therefore, we analyzed serial samples collected before and after treatment to approximate CH biology. In patients achieving complete remission, post-treatment samples contained residual 8+ HSCs without malignant blasts, representing a clonal but non-malignant state analogous to CH.

By intersecting 723 differentially expressed genes in 8+ HSCs (adjusted p<0.05) with genes located on chr8, 40 overlapping genes were identified (**Fig. 6d-e** and **Table S13**). Of these, *MYC* was the highest-ranked gene within the disease-associated 8q21.3-24.23 locus identified by LASSO-Cox regression **(Fig. 6c**). Although all patients analyzed had whole chr8 gain, regions of increased transcriptional activity were focally enriched within 8q21.3-24.23 in the HSPC clusters, consistent with the locus identified in our LASSO-Cox analyses (**Supplementary Fig. 16a-e**). Further, patients with 8+ had significantly higher levels of MYC protein expression across myeloid diseases (**Supplementary Fig. 17a-c**). Together, these findings prioritize *MYC* as a candidate gene within the 8+ locus associated with disease risk.

### Functional validation of mCA effector genes driving systemic disease

To test causality, we assessed inflammation and pathology in the Mx1-Cre^+/-^;Rosa26^LSL-MYC/LSL-MYC^ mouse model, in which *MYC* expression is induced by polyinosinic-polycytidylic acid (pIpC) in the HSC compartment (**Supplementary Fig. 17d-f**)^35,38^. Mice with *MYC* overexpression developed monocytosis, anemia, and HSPC expansion, with significantly reduced OS (**Fig. 6f-h** and **Supplementary Fig. 17g-i**). *MYC* overexpression also provoked marked fibrosis in multiple organs, including BM, spleen, liver, and heart (**Fig. 6i**). These findings are consistent with observations in 8+ carriers, who showed monocytosis, anemia, reduced OS, and elevated CRP in high-MCF carriers, with trends toward higher TNFα and IL6, compared to non-carriers (**Fig. 6j-l** and **Supplementary Fig. 17j-n**). Thus, programming murine HSCs to overexpress *MYC* at levels comparable to those observed in HSCs of 8+ carriers is sufficient to provoke systemic inflammation and multiorgan pathology.

To assess if this integrative platform can be generalized to other mCAs, we extended our analysis to 1+. In the HSC cluster, 588 genes were differentially expressed in 1+ cells vs. healthy donor cells (adjusted p<0.05); of these, 80 genes mapped to chr1 (**Supplementary Fig. 18a-c** and **Table S14**). Among the top-ranked candidates, *S100A9* emerged as the highest-ranked inflammatory gene within this disease-associated locus (**Supplementary Fig. 18d**). As observed in 8+, InferCNV analysis revealed that transcriptional activation in 1+ HSPCs was focally enriched within 1q21.1-23.2 (**Supplementary Fig. 16a-e**), a region that harbors *S100A9* and is associated with significantly increased risk of multiple diseases (**Supplementary Fig. 11a**). In mice, enforced overexpression of *S100A9* in HSCs at levels seen in 1+ HSPCs was sufficient to provoke multiorgan fibrosis and reduced OS in vivo (**Supplementary Fig. 18e-m**), and 1+ carriers showed elevated circulating pro-inflammatory cytokines and alarmins (**Supplementary Fig. 19a-f**).

Thus, this integrated epidemiological, bioinformatics, and experimental framework enables identification and functional validation of key genes driving mCA-associated disease risk across distinct aneuploidies. Notably, the convergence of 8+ (via MYC) and 1+ (via S100A9) on inflammatory signaling and multiorgan pathology highlights inflammation as a shared mechanism by which at least some mCAs drive systemic disease.

## DISCUSSION

CH has emerged as a systemic phenomenon with broad implications across multiple organ systems and represents a critical intersection between aging biology and disease susceptibility^1–4^. While CHIP has been extensively characterized as a driver of diverse age-related pathologies through inflammatory signaling^5–12^, mCAs have remained understudied beyond hematologic malignancies^18–21^. Here, we addressed this gap through comprehensive phenome-wide analyses integrated with cytogenetic mapping, single-cell transcriptomics, and in vivo modeling.

Integrated analyses of 452,594 UKBB participants, coupled with scRNA-seq and mouse model studies, established that mCAs are pleiotropic drivers of multimorbidity and mortality, with effect sizes comparable to CHIP (all-cause mortality HR=1.21 vs. HR=1.34 for CHIP^39^). These associations increase with MCF and are amplified in individuals harboring complex mCAs. Select alterations, including 3+, 8+, 9=, 12+, and Y-, have broad disease associations. Notably, many mCAs are linked to inflammaging-related disorders, implicating chronic inflammation as an underlying mechanism analogous to CHIP-mediated effects^2,5,29^. Thus, together with CHIP, mCAs are major contributors to age-related disease burden.

While mCAs collectively are associated with both increased incidence and mortality for hematologic, respiratory, neoplastic, infectious, and nervous system disorders, CVD showed increased mortality, consistent with a prior report^40^, in the absence of increased incidence. This pattern suggests mCAs may modulate disease progression or case-fatality rather than initiation, though alternative explanations, including baseline lethality, differential diagnostic intensity, and first-presentation fatal events, cannot be excluded. Conversely, some mCA-associated organ system disorders (e.g., mental/behavioral, sensory, and digestive) have elevated incidence without effects on mortality. This highlights the need for different clinical strategies for individuals with mCAs, where those having organ system disorders with elevated mortality warrant aggressive management of established diseases, whereas those having only elevated incidence may benefit from preventive strategies. Importantly, these patterns persist after exclusion of individuals with CHIP, establishing mCAs as independent modulators of both disease incidence and survival.

Aggregate analyses, however, mask substantial heterogeneity at the level of individual mCAs. Previous studies have treated mCA status as binary (present vs. absent)^40–42^, precluding identification of alteration-specific disease associations. Analysis of individual mCAs established that specific alterations confer distinct risk profiles: for instance, 8+ and 9= are associated with increased CVD risk despite no aggregate signal. These findings emphasize the importance of mCA-specific risk assessment and indicate that clinical management should be tailored to the specific alteration(s) carried by each individual.

A key clinical challenge in managing CH individuals is determining the optimal timing and scope of screening and intervention. Currently, incidentally detected mCAs on clinical genomic testing lack standardized management guidelines, and most individuals receive no systematic follow-up. The mediation analyses reported here revealed that mCAs can influence non-hematologic diseases via direct pathways and/or indirect mechanisms mediated by hematologic malignancies, which has important implications for surveillance. For mCAs with predominantly mediated effects, such as 9= (associated with increased CVD risk primarily through MPN development), early detection and treatment of the hematologic malignancy may substantially reduce downstream CVD burden. In contrast, mCAs with mixed direct and mediated effects, such as 12+ on chronic bronchitis, may require broader monitoring strategies beyond hematologic assessment. Individuals with such alterations may benefit from baseline screening and preventive intervention (e.g., smoking cessation) independent of hematologic malignancy status. Finally, as higher MCF and complex mCAs are associated with particularly elevated mortality risk, intensified surveillance in these subgroups should be considered. Collectively, these results support mCA-specific surveillance strategies informed by disease pathways, clone burden, and genomic complexity.

Mapping large-scale chromosomal alterations to specific disease phenotypes has been a major challenge in mCA research. We addressed this challenge by developing a cytoband-level mapping approach using LASSO-penalized Cox regression to prioritize disease-associated loci within heterogeneous mCAs. This method identified cytobands harboring oncogenes and inflammatory activators within gain-associated regions, and tumor suppressors or negative regulators of inflammation within loss-associated regions. CNLOH regions similarly revealed candidate genes with potential pathogenic roles through biallelic changes. Because mCAs encompass large genomic regions, direct experimental modeling of chromosomal dosage effects is infeasible. We therefore adopted a reductionist, sufficiency-based strategy, testing whether candidate genes prioritized from mCA-associated loci could recapitulate key disease-relevant phenotypes without implying that they fully explain mCA biology. scRNA-seq analysis of BM cells from patients with 8+ and 1+ demonstrated focal transcriptional activation within 8q21.3-24.23 and 1q21.1-23.2 loci in HSCs, with *MYC* and *S100A9* emerging as candidate effectors. Using genetically engineered mouse models, enforced expression of *MYC* or *S100A9* in HSCs was sufficient to induce systemic inflammatory and fibrotic phenotypes, supporting their causal contribution to selected disease manifestations associated with 8+ and 1+. This integrated approach is generalizable to other mCAs, providing a platform for identifying gene-level effectors. Analogous to emerging efforts targeting recurrent gene mutations in CH (e.g., IDH1/2 inhibitors in *IDH1/2*-mutant clonal cytopenia of undetermined significance; NCT05030441, NCT06566742, NCT05102370)^43^, our framework may enable mechanism-informed risk stratification and precision interventions for mCA carriers based on perturbed genes.

Several important areas merit further investigation. First, the cytoband mapping and functional validation approach outlined here should be expanded beyond 8+ and 1+ to systematically identify driver loci and genes across all disease-associated mCAs, ideally with validation in CH-stage samples. Loss-associated mCAs represent equally important targets, potentially addressable via CRISPR screening or in vivo knockout models. Second, although whole-chromosome gains increase the dosage of all genes, we observed preferential transcriptional activation confined to specific cytobands. If shared mechanisms underlie this selective activation across distinct mCAs, they may represent convergent, targetable vulnerabilities. Third, as MYC drives S100A9-mediated inflammatory signaling^35^, and S100A9 appears causative in incidence and mortality associated with 1+ and other mCAs (e.g., del(5q) haploinsufficiency in MDS^44^), dissecting the inflammatory cascades linking mCAs to multiorgan pathology is an important priority. Fourth, although no interaction between mCAs and CHIP was observed for disease incidence at the aggregate level, synergistic effects on mortality were evident. Thus, analyses of specific mCA-mutation combinations may reveal heterogeneous effects on disease incidence and survival that are obscured in aggregate analyses, enabling more refined risk stratification. Fifth, prospective studies are needed to determine if mCA-informed surveillance strategies improve clinical outcomes and are cost-effective for population-level implementation. Finally, the unexpected protective associations observed for certain mCAs (e.g., Y- with reduced risk of gout, diabetes, hypertension) highlight context-dependent biology requiring mechanistic studies.

Several limitations of this work should be acknowledged. First, homogeneous mCAs affecting entire chromosomes could not be resolved at the cytoband level, limiting candidate gene identification. Second, the extreme rarity of 8+ and 1+ limits the ability to obtain BM samples at the CH stage; we therefore used post-treatment remission samples containing residual trisomy-positive HSCs to approximate a CH-like state. Validation using bona fide CH-stage samples would strengthen these findings. Third, mCA assessment on chrY was limited to the distal p arm, precluding analysis of other chrY loci. Fourth, mCA-disease associations were not replicated in external cohorts, as our primary aim was to establish causality through functional validation. Fifth, although analyses were restricted to incident diseases following mCA assessment and were adjusted for disease burden, subclinical conditions at baseline may have influenced both mCA expansion and disease diagnosis. Finally, the UKBB is predominantly European, which limits generalizing these findings to other ethnic groups, particularly for population-specific mCAs.

In conclusion, our study provides the first systematic characterization of individual mCAs as pleiotropic drivers of age-related multimorbidity and mortality independent of CHIP and provides a generalizable platform for linking mCAs to disease phenotypes and candidate driver genes. These findings have implications for risk stratification, surveillance, and therapeutic development in individuals with CH.

## METHODS

### Study population and data used in analysis

From an initial pool of 482,615 UKBB participants with previously completed mCA assessment (Loh et al. ^20^), we applied a systematic filtering criteria to ensure data quality and definitive mCA classification. First, 309 participants were excluded as having invalid or missing mCA assessment dates, which are defined as dates that were missing (n=32), equal to 1900-01-01 (n=277), or on or before 1901-01-01 (n=0), as these represented data entry errors or placeholder values. This filtering step yielded 482,306 participants with valid assessment dates spanning March 13, 2006, to October 1, 2010. Next, mCA status was classified based on CNV analysis. Participants were categorized as mCA positive if any CNV with a known copy change (i.e., gain, loss, or CNLOH) was detected, or mCA negative if no CNVs with known copy changes were detected and at least one non-unknown CNV was present. To ensure definitive classification and avoid misclassification bias, we excluded 19,712 (6.2%) participants who had only CNVs with unknown copy change status, as their true mCA status could not be reliably determined. After these exclusions, our final analytical cohort comprised 452,594 participants, including 42,390 (9.4%) mCA positive and 410,204 (90.6%) mCA negative individuals. All subsequent analyses, including disease incidence and mortality analyses, were restricted to this well-characterized cohort with definitive mCA classification and valid temporal data.

### Cross-sectional association analyses of demographic, lifestyle, and hematologic parameters with mCA status

To identify factors associated with mCA prevalence, we performed cross-sectional logistic regression analyses comparing mCA-positive participants against mCA-negative controls. We examined associations across three categories of variables: (i) demographic factors (age at mCA assessment, sex); (ii) lifestyle factors (ever smoking, current smoking, ever alcohol use, current alcohol use, body mass index, and obesity defined by ICD-10 code E66.0); and (iii) hematologic and biochemistry parameters.

For demographic and lifestyle factors, we fitted univariate logistic regression models with mCA status as the binary outcome. For CBC and biochemistry parameters, values were natural log-transformed using log(1+x) to normalize distributions, and models were adjusted for age at mCA assessment and sex. Odds ratios (ORs) with 95% confidence intervals (CIs) were estimated by exponentiating model coefficients, and p-values were calculated using Wald tests on the regression coefficients. These analyses were extended to the six mCA subtypes described above, comparing participants harboring each specific alteration against mCA-negative controls. To account for multiple testing, the Benjamini-Hochberg FDR procedure was applied separately within each mCA subtype, with significance defined as FDR-adjusted q<0.05. To compare CBC parameters between participants harboring specific chromosomal gains (e.g., 8+) and mCA-negative controls, we used the Wilcoxon rank-sum test, a non-parametric method appropriate for comparing distributions between two independent groups. Distributions were visualized using violin plots with overlaid boxplots showing median and interquartile range; mean values were indicated by diamond symbols.

### Competing risk analysis of disease incidence

Disease incidence was ascertained from hospital inpatient admission records linked to the UKBB. Diagnoses were recorded using ICD-10 codes, analyzed across 13 mutually exclusive disease categories. This categorization approach was chosen to enable comprehensive disease spectrum analysis and to ensure consistent multiple testing correction across both incidence and mortality analyses. Follow-up began at the date of mCA assessment (defined by first blood sample collection date) and continued until first incident disease diagnosis, death, loss to follow-up, or July 14, 2023, whichever occurred first. Participants who died before their mCA assessment date were excluded from analysis to prevent immortal time bias. Follow-up time was calculated in years as the difference between the event date (or censoring date) and the mCA assessment date.

To identify truly incident (new onset) disease, we employed a rigorous exclusion strategy for pre-existing conditions. For each participant and disease category, we catalogued all ICD-10 diagnosis codes recorded in hospital admissions on or before the mCA assessment date. Post-mCA admissions were then examined, and any diagnoses matching pre-existing codes for that participant were excluded. This code-level approach ensures that we captured only genuinely new disease diagnoses rather than recurrent admissions for chronic conditions. Incident disease was defined as the first occurrence of any qualifying ICD-10 code after mCA assessment that had not been previously recorded.

Death prior to disease diagnosis was treated as a competing risk rather than censoring, as deceased participants cannot experience incident diseases. For each disease category, participants were classified as: (i) incident disease (event of interest, coded as 1); (ii) death before disease diagnosis (competing event, coded as 2); or (iii) alive and disease-free at the end of follow-up (censored, coded as 0). We employed competing risk methods to assess mCA-disease incidence associations. Cumulative incidence functions (CIFs) were compared between mCA-positive vs. mCA-negative groups using Gray’s test. We estimated subdistribution hazard ratios (HRs) using Fine-Gray regression, which appropriately accounts for competing risks by modeling the subdistribution hazard (the instantaneous rate of occurrence of the event of interest in the presence of competing events). We fitted two models per disease category: (i) unadjusted, with only mCA status; and (ii) adjusted for age at mCA assessment, sex, and baseline disease burden (number of distinct ICD-10 diagnoses prior to mCA assessment). The adjusted model accounts for potential confounding by demographic factors and pre-existing health status. The subdistribution HR represents the ratio of the instantaneous rate of the event in mCA vs. no mCA groups, accounting for the competing risk. P-values for the adjusted models were calculated using Wald tests on the coefficient for mCA status. For the 13 ICD-10 disease category analyses (mCA vs. no mCA), we required more than 10 total events with events present in both comparison groups; analyses not meeting these thresholds were excluded from reporting.

To assess if specific mCA subtypes were differentially associated with disease risk, we extended the analysis to 72 individual mCA subtypes defined by chromosome (1-22, X, Y) and CNV type (gain, loss, or CNLOH). For each mCA subtype, participants harboring that specific alteration were compared against those with no detectable mCA. We first analyzed associations between the 72 mCA subtypes and each of the 13 ICD-10 disease categories (936 tests). We then extended this analysis to 87 individual inflammaging-related diseases grouped into 11 pathophysiological categories, with combined endpoints created for each category as well as an overall inflammaging endpoint (6,264 tests). For both the mCA subtype x ICD-10 disease category analyses and the mCA subtype x inflammaging disease analyses, we applied stringent event thresholds to ensure statistical stability given the larger number of comparisons: unadjusted analyses (Gray’s test and unadjusted Fine-Gray regression) required a minimum of 10 total events with at least 5 events in each comparison group, while adjusted Fine-Gray regression models required a minimum of 30 total events with at least 5 events per group to ensure adequate power for estimating coefficients for mCA status and three additional covariates. Results were visualized using heatmaps displaying log2 transformed subdistribution HRs, with hierarchical clustering applied to identify patterns of mCA-disease associations.

To account for multiple testing, we applied the Benjamini-Hochberg false discovery rate (FDR) procedure separately within each analysis: mCA status vs. 13 ICD-10 categories, 72 mCA subtypes vs. 13 ICD-10 categories, and 72 mCA subtypes vs. inflammaging diseases. The FDR threshold was set at 0.05, and FDR-adjusted q-values were reported alongside raw p-values. This procedure controls the expected proportion of false positives among rejected hypotheses. All analyses were performed using R version 4.5.1. Competing risk analyses were conducted using the cmprsk package for Gray’s test and Fine-Gray regression, and the tidycmprsk package for estimating and visualizing cumulative incidence functions. Data manipulation was performed using the dplyr and data.table packages. All statistical tests were two-sided.

### All-cause mortality in mCA subtypes

To evaluate mortality risk across different mCA subtypes, we performed all-cause mortality analyses comparing each subtype against mCA negative participants. From the 42,390 mCA positive participants in our analytical cohort, we defined seven mutually non-exclusive subtypes based on genomic characteristics: (i) all mCA (n=42,390); (ii) complex mCA, defined as mCAs affecting ≥3 unique chromosomes (n=370); (iii) high MCF, defined as any mCA with cell fraction ≥10% (n=13,686); (iv) low MCF, defined as all mCAs with cell fraction <10% (n=28,704); (v) gain-only, participants with only copy number gains and no losses or CNLOH (n=1,466); (vi) loss-only, participants with only copy number losses and no gains or CNLOH (n=32,187); (vii) neutral-only, participants with only CNLOH and no gains or losses (n=7,093). Follow-up methods and time calculations were identical to those described for the competing risk analyses. For each subtype, we created a binary comparison dataset comprising subtype-positive participants and all mCA negative controls (n=410,204). Unlike the cause-specific mortality analysis which analyzed deaths within 13 specific disease categories using competing risk methods, all-cause mortality analysis included deaths from any cause and used standard survival methods as there were no competing events. We fitted Cox proportional hazards models to estimate HRs for each subtype. Two models were constructed per subtype: (i) an unadjusted model with subtype status only; and (ii) an adjusted model including age at mCA assessment, sex, and baseline disease burden. The HR represents the ratio of the instantaneous mortality rate in each subtype compared to mCA-negative participants. P-values were calculated using Wald tests on the coefficient for subtype status. To account for multiple testing across the seven subtypes, we applied the

Benjamini-Hochberg FDR procedure to both unadjusted and adjusted model p-values, setting the FDR threshold at 0.05. Statistical analyses were conducted using R with the survival package for Cox regression.

### Competing risk analysis of cause-specific mortality

Mortality data were obtained from death registry records through the UKBB data portal. Cause of death was determined using ICD-10 codes from death certificates, analyzed across the same 13 mutually exclusive disease categories as the incidence analysis. Follow-up methods, time calculations, and exclusion criteria were identical to those described for the incidence analysis. The median follow-up time was 14.34 years (IQR 13.6-15.06 years), during which 38,282 deaths occurred (6,759 and 31,523 in mCA positive and negative groups, respectively).

For each disease category, we constructed cause-specific event indicators: death from the specified cause (event of interest, coded as 1), death from any other cause (competing event, coded as 2), or censored (alive at end of follow-up, coded as 0). We applied the same competing risk framework as described for disease incidence, using Gray’s test to compare CIFs between mCA groups and Fine-Gray regression to estimate subdistribution HRs. Models were fitted with identical covariate adjustment strategies: an unadjusted model with mCA status only, and an adjusted model including age at mCA assessment, sex, and baseline disease burden. Multiple testing correction was performed using the Benjamini-Hochberg FDR procedure with a threshold of 0.05, identical to our incidence analysis. Statistical analyses were conducted using the same R packages and methods as described above.

### Sensitivity analysis excluding participants with somatic mutations and interaction analysis

To assess if the association between mCAs and disease outcomes are independent of somatic mutations, we performed sensitivity analyses excluding participants with clonal hematopoiesis of indeterminate potential (CHIP). CHIP status was obtained from data returned to UK Biobank by Vlasschaert et al.^45^. In this study, CHIP was annotated from whole-exome sequencing data using Mutect2 across 74 canonical CHIP driver genes. Participants were classified as CHIP-positive if they harbored at least one somatic mutation and CHIP-negative if no mutations were detected. Of the 452,594 participants in the primary analysis, 15,620 (3.5%) harbored somatic mutations and were excluded. An additional 32,506 participants lacking somatic mutation assessment data were also excluded to avoid misclassification, yielding a final cohort of 404,468 individuals. Among these, 36,725 (9.1%) carried mCAs and 367,743 (90.9%) did not.

For disease incidence, we repeated the competing risk analyses as described above on the CHIP-excluded cohort, comparing cumulative incidence and estimating subdistribution HRs for each ICD-10 disease category. For mortality, Kaplan-Meier survival analysis was performed comparing OS between mCA carriers and non-carriers using the log-rank test, followed by Cox proportional hazards regression adjusted for age, sex, and baseline disease burden.

To formally assess the joint effects of mCA and CHIP on disease incidence and mortality, we performed an interaction analysis among 418,870 participants with available data for both mCA and CHIP status. Participants were stratified into four mutually exclusive groups: (1) mCA-/CHIP- (reference group, n=367,743), (2) mCA+/CHIP- (n=36,725), (3) mCA-/CHIP+ (n=12,347), and (4) mCA+/CHIP+ (n=2,055). For disease incidence, cumulative incidence curves accounting for death as a competing risk were generated for each group, with statistical significance assessed using Gray’s test. Fine-Gray subdistribution hazard regression was used to estimate subdistribution HRs for each group compared to the double-negative reference, adjusted for age, sex, and baseline disease burden. For mortality, Kaplan-Meier survival curves were generated with statistical significance assessed using the log-rank test, and pairwise comparisons were performed with Benjamini-Hochberg correction. Cox proportional hazards regression was used to estimate adjusted HRs for each group compared to the double-negative reference. For both outcomes, statistical interaction on the multiplicative scale was tested by fitting models including main effects for mCA status, CHIP status, and their product term (mCA × CHIP), along with covariates. For disease incidence, interaction on the additive scale was also assessed by calculating the relative excess risk due to interaction (RERI), attributable proportion (AP), and synergy index (S), with 95% CIs estimated using the delta method. A significant positive interaction term (p<0.05) would indicate a synergistic effect, whereby the combined presence of mCA and CHIP confers excess risk beyond that expected from multiplying their individual effects.

### Dose-response analysis of mosaic cell fraction

To assess if disease risk varies with mCA burden beyond simple presence/absence, we performed dose-response analyses examining the relationship between MCF and incident disease risk. These analyses were conducted using the same analytical cohort of 452,594 participants with definitive mCA classification described above, applying identical follow-up methods, time calculations, and covariate adjustments.

For each mCA subtype-disease combination, we fitted Cox proportional hazards models with natural cubic splines (df=3) to flexibly model the dose-response relationship between MCF and disease risk. This approach allows for potentially non-linear relationships, such as threshold effects where risk increases only above a certain clone size, or saturation effects where risk plateaus at high cell fractions. All models were adjusted for age at mCA assessment, sex, and baseline disease burden. Death prior to disease diagnosis was treated as a censoring event in this cause-specific hazard analysis, as our primary interest was the direct relationship between MCF and disease incidence rather than the subdistribution incorporating competing mortality. We assessed overall significance of the dose-response relationship using a joint Wald test for the spline basis coefficients. To ensure stable parameter estimation, we required a minimum of 30 disease events overall, at least 5 events in both exposed and unexposed groups, and at least 50 mCA positive participants for spline model fitting. Analyses not meeting these thresholds were excluded. To account for multiple testing across all mCA subtype-disease combinations, we applied Benjamini-Hochberg FDR correction, with significance defined as FDR-adjusted q<0.05.

To visualize dose-response relationships at high resolution, we performed binned analyses for mCA-disease pairs showing significant spline effects. MCF values were divided into 5-percentage-point bins, and HRs were estimated for each bin compared to mCA negative controls, adjusted for age, sex, and baseline disease burden.

### Mediation analyses of mCAs and disease pathways

To assess if specific mCAs confer disease risk directly or through intermediate hematologic malignancies, we performed formal mediation analyses using both time-to-event and binary outcome approaches. We focused on three mCA-disease pairs with prior evidence suggesting hematologic intermediates: (i) 9= and CVD potentially mediated through MPN; (ii) 13- and actinic keratosis potentially mediated through CLL; and (iii) 12+ and chronic bronchitis potentially mediated through CLL. For each pathway, analyses compared participants carrying the specific mCA of interest (regardless of other concurrent mCAs) to mCA negative controls (participants without any detectable mCAs).

We employed a sequential Cox proportional hazards modeling approach to decompose the total effect of each mCA on disease outcomes into direct and indirect (mediated) components. For each mCA-mediator-outcome pathway, we fitted three Cox models, all adjusted for age at mCA assessment, sex, and baseline disease burden. First, we modeled time from mCA assessment to first incident hematologic malignancy as a function of mCA status (pathway A: exposure → mediator). Second, we modeled time to incident disease outcome as a function of mCA status without including the mediator, representing the total (combined direct plus indirect) effect. Third, we modeled time to incident disease outcome as a function of both mCA status and mediator status (binary indicator of incident hematologic malignancy), representing the direct effect after accounting for mediation. This model also quantified pathway B (mediator → outcome). Mediator status was coded as a binary time-fixed indicator (0/1) representing whether a participant ever developed hematologic malignancy during follow-up. Evidence for mediation was indicated by significant associations in pathways A and B, and attenuation of the mCA effect when comparing total to direct effect models. Full mediation was concluded when the direct effect became non-significant (p≥0.05) after adjusting for the mediator, while partial mediation was concluded when the direct effect remained significant but was attenuated.

To quantify the proportion of total effect explained by mediation, we performed binary mediation analysis using the mediation package in R. We fitted two generalized linear models with binomial family: first, regressing mediator status on mCA status, and second, regressing outcome status on both mCA status and mediator status, using the same covariates as the Cox models (age, sex, and baseline disease burden). We estimated the Average Causal Mediation Effect (ACME, the indirect effect through the hematologic malignancy), the Average Direct Effect (ADE, the direct effect not mediated through the hematologic malignancy), the Total Effect (the sum of ACME and ADE), and the Proportion Mediated (ACME/Total Effect) via quasi-Bayesian Monte Carlo simulation with 1,000 iterations and robust standard errors. Results were reported as absolute risk differences (in percentage points) with 95% CIs. Because the binary mediation analysis treats both mediator and outcome as fixed binary variables, it complements, but does not duplicate, the time-to-event Cox models used for the primary mediation interpretation.

Given the combination of rare mCA exposures and strong effect sizes for mCA-to-malignancy associations, we performed comprehensive diagnostics to ensure model reliability and distinguish genuine biological effects from statistical artifacts. For each mediation pathway, we examined 2×2 contingency tables for exposure-mediator, exposure-outcome, and mediator-outcome relationships to detect potential complete separation (cells with zero observations) or quasi-separation (cells with <5 observations), both of which can produce unstable parameter estimates. We verified logistic regression model convergence by confirming that Fisher scoring iterations were ≤10 and examined standard errors to ensure no coefficients exceeded 2.0, which would indicate quasi-separation. For Cox proportional hazards models, we assessed model fit using concordance statistics and likelihood ratio tests. These analyses demonstrated adequate model fit with no evidence of complete separation (minimum cell count ≥7 across all contingency tables), appropriate convergence (4-10 iterations for all models), and reasonable standard errors (maximum 0.25, all others <0.17). The very large HRs observed for mCA-to-malignancy pathways are consistent with established biological relationships where these specific mCAs represent characteristic genetic features of their respective malignancies.

### Classification of homogeneous vs. heterogeneous mCAs

To characterize the genomic consistency of mCAs across individuals, we developed a bin-based approach to quantify whether mCAs of a given type (i.e., gain, loss, or CNLOH) affect similar chromosomal regions across carriers or exhibit variable breakpoints and genomic footprints. For each chromosome, we divided the genomic span into 1 Mb bins (0.5 Mb for chrY due to its smaller size) and constructed a binary matrix indicating the presence or absence of mCA coverage in each bin for every affected individual. Using this matrix, we derived four complementary metrics to quantify across-sample similarity.

First, we computed a homogeneity index, defined as the proportion of bins with any mCA coverage in which more than 50% of samples shared overlapping alterations. This metric captures cross-sample consistency in affected regions. Second, we calculated the Gini coefficient of bin coverage to determine if mCA footprints were uniformly distributed across the chromosome or concentrated in focal regions, with lower values indicating more uniform coverage. Third, we computed normalized coverage entropy by discretizing bin coverage into quintiles and calculating Shannon entropy, which quantifies the dispersion of mCA signal across genomic bins; lower entropy indicates that coverage is concentrated in specific regions rather than scattered. Fourth, we determined the mean coverage, representing the average proportion of samples covering each bin, with higher values indicating that most genomic bins are consistently affected.

A mCA subtype was classified as homogeneous if it satisfied all of the following criteria: (i) homogeneity index >0.9, indicating strong cross-sample consistency; (ii) Gini coefficient <0.25, indicating coverage is not focal or clustered; (iii) normalized coverage entropy <0.4, indicating the CNV signal is concentrated rather than dispersed; and (iv) mean coverage >0.75, indicating that the majority of bins are affected across samples. mCA subtypes failing to meet all four criteria were classified as heterogeneous. These thresholds distinguish mCAs with stereotyped genomic boundaries (e.g., whole-chromosome or whole-arm events) from those with variable breakpoints across individuals. To assess the stability of these classification metrics, we performed bootstrap resampling (B=1,000 iterations) by resampling individuals with replacement and recalculating all four metrics for each iteration. We report 95% CIs derived from the 2.5^th^ and 97.5^th^ percentiles of the bootstrap distributions.

### Cytoband-level feature selection and hazard estimation for mCA-specific disease risk

To identify candidate cytogenetic bands associated with disease incidence, we performed a two-stage analytical approach combining LASSO-penalized Cox regression for feature selection followed by standard Cox proportional hazards modeling for HR estimation. We chose LASSO (L1 penalty) over ridge regression (L2 penalty) because our goal was feature selection rather than prediction; LASSO produces sparse solutions by shrinking coefficients to exactly zero, enabling identification of specific disease-associated cytobands. The analysis was restricted to heterogeneous mCAs, as these exhibit variable breakpoints across individuals and may harbor distinct disease-associated regions within the same chromosome; homogeneous mCA events (i.e., 12+, 14+, 15+, 18+, and X-) have uniform genomic boundaries across carriers, making cytoband-level resolution uninformative. Participants harboring both homogeneous and heterogeneous mCAs were retained and analyzed for their heterogeneous events. CNVs were analyzed at the cytoband level rather than the individual gene level, as gene-level analysis would be severely underpowered given the sparse distribution of mCAs across thousands of genes. CNVs were mapped to cytogenetic bands using the UCSC hg19 cytoband annotations. For each chromosome, overlapping cytobands were identified for all CNV segments using the GenomicRanges package in R. Binary indicator variables were created for each unique combination of cytoband and CNV type (gain, loss, or CNLOH), resulting in features such as “1p36.33_loss” or “12q24.31_CNLOH”. The analysis compared participants harboring mCAs on the chromosome of interest against participants with no detectable mCA on any chromosome.

This penalized regression approach efficiently handles the high-dimensional and correlated features arising from overlapping CNV segments. The optimal penalty parameter (λ) was determined using 10-fold cross-validation with a fixed random seed for reproducibility, by selecting the value that minimized the cross-validated partial likelihood deviance (lambda.min), and cytobands with non-zero coefficients at this optimal λ were retained for subsequent analysis. We chose lambda.min over the more conservative lambda.1se to allow for more inclusive screening. Cytobands with non-zero coefficients at the optimal λ were refitted into standard Cox proportional hazards models to obtain unbiased HR estimates with 95% CIs. Both unadjusted and adjusted models were constructed, with adjusted models including age at mCA assessment, sex, and baseline disease burden as covariates. Death prior to disease diagnosis was treated as a censoring event. Incident cohort construction and follow-up definitions followed the procedures described for the competing risk analysis of disease incidence. Benjamini-Hochberg FDR correction was applied globally across all cytoband-disease category combinations.

To ensure stable model estimation, minimum event thresholds were applied at both the chromosome and cytoband levels. At the chromosome level, analysis proceeded only when (i) the total number of incident disease events exceeded 10, and (ii) events were observed in both the mCA carrier and non-carrier groups. At the cytoband level, Cox regression was performed only for cytobands meeting the following criteria: (i) variation in marker status (i.e., at least one participant with and at least one without the cytoband-specific CNV), (ii) at least one event in both the marker-positive and marker-negative groups, and (iii) a combined total of more than 10 events across both groups. Chromosome–disease category combinations and individual cytobands failing to meet these thresholds were excluded from analysis.

To assess the robustness of LASSO-selected cytobands, we performed bootstrap stability analysis with 1,000 iterations. To improve computational efficiency, controls (no mCA) were subsampled at a 4:1 ratio relative to mCA cases prior to bootstrap analysis. For each iteration, we resampled participants with replacement and refitted the LASSO-penalized Cox regression model using identical procedures as described above. Bootstrap selection frequency was calculated as the proportion of iterations in which each cytoband was selected (i.e., received a non-zero coefficient). Cytobands with selection frequency ≥70% were classified as highly stable. To validate the concordance between LASSO and Cox regression estimates, we compared LASSO coefficients with log-transformed HRs from standard Cox models. Direction concordance was defined as the proportion of cytobands for which LASSO coefficients and Cox log(HR) values shared the same sign (both positive or both negative). Pearson correlation was computed to quantify the strength of the linear relationship between LASSO coefficients and Cox log(HR) estimates. The results were visualized using scatter plots displaying bootstrap selection frequency vs. Cox FDR significance, and LASSO coefficients vs. Cox log(HR), with points colored by bootstrap stability category (<50%, 50-70%, ≥70%).

### Circos plot visualization

To provide a genome-wide overview of mCA-disease associations, we generated circos plots displaying cytoband-level HRs across all chromosomes using the circlize package in R. For each CNV subtype, separate plots were constructed comprising multiple concentric tracks. The outermost track displayed the chromosomal ideogram with cytogenetic band annotations based on UCSC hg19 coordinates. The second track depicted the distribution of CNV segments across the genome, with segments colored by CNV subtype (red for gains, blue for losses, orange for CNLOH). The inner six tracks displayed log2-transformed adjusted HRs for six ICD-10 disease categories selected based on the number of significant cytoband associations.

For heterogeneous mCAs, HRs were derived from standard Cox regression analysis described above, with cytobands meeting FDR<0.05 displayed as vertical bars at their genomic midpoint positions. For homogeneous mCAs (12+, 14+, 15+, 18+, and X-), which have uniform genomic boundaries across carriers precluding cytoband-level resolution, we performed standard Cox proportional hazards regression comparing carriers of each homogeneous mCA against participants with no detectable mCA, adjusting for age at mCA assessment, sex, and baseline disease burden. The resulting chromosome-wide HR was then applied uniformly across all cytobands of the affected chromosome, with FDR correction performed separately for these 65 tests (5 homogeneous mCAs × 13 disease categories). Within each disease category track, positive log2(HR) values (increased risk) were displayed in red and negative values (decreased risk) in blue. Each track employed category-specific y-axis scaling to optimize visualization of effect sizes, with axis ranges indicated in the figure legend. Bar width was set to 2 Mb to ensure visibility of individual cytoband associations.

### Gene annotation of disease-associated cytobands

To characterize the potential functional consequences of significant mCA-disease associations identified through LASSO-Cox regression, we systematically catalogued all genes and microRNAs (miRNAs) located within cytobands that demonstrated significant associations with disease incidence. Cytobands meeting the significance threshold (FDR-adjusted q<0.05) from the cytoband-level LASSO-Cox analysis were selected for gene annotation. This analysis was restricted to heterogeneous mCAs, as homogeneous mCAs affect entire chromosomes or chromosome arms uniformly across carriers, precluding meaningful cytoband-level gene prioritization.

Gene annotations were retrieved from Ensembl (GRCh37/hg19 assembly) using the biomaRt package in R. For each significant cytoband, we queried all genes with genomic coordinates overlapping the cytoband boundaries as defined by UCSC hg19 cytoband annotations. Retrieved attributes included Ensembl gene identifier, gene symbol, gene biotype, chromosomal coordinates, and cytogenetic band assignment. Genes were classified into two categories: protein-coding genes (including all non-miRNA biotypes such as protein-coding, long non-coding RNA, and pseudogenes) and miRNAs. Genes lacking annotated gene symbols were excluded from downstream analyses.

### Patient selection for single-cell analysis

Using the Moffitt Cancer Center (MCC) Total Cancer Care protocol, a prospective biorepository and clinical data registry, we identified cases diagnosed with MDS, MPN, MDS/MPN, or AML. All patients provided written informed consent, and our study was approved by the MCC IRB (MCC#21675, MCC#22471, MCC#23050). Healthy donors (HDs) BM aspirates were commercially obtained from Lonza (Cat#1M-105). Given the extreme rarity of 8+ and 1+ in the general population (<0.05%), prospective collection of BM samples from individuals at the CH stage is not feasible. To address this limitation, we employed a serial sampling strategy. For each patient, we collected BM samples at diagnosis (pre-treatment) and following therapy (post-treatment). Patients achieving complete remission contain residual 8+ or 1+ HSCs and progenitors without the confounding presence of malignant blasts. This approach approximates a “CH-like” state in which aneuploid cells persist without overt malignancy, enabling analysis of the transcriptional consequences of aneuploidy in a non-malignant context.

### Single-cell library production and sequencing

BM mononuclear cells (MNCs) from MPN, MDS, MDS/MPN, and AML patients and HDs were prepared following the protocol provided by 10x Genomics (CG000392, Rev A). For single cell encapsulation and library production, we used the Chromium Single Cell 5’ Reagent Kits (v2) per user guideline (CG000331). Sequencing was performed on a NovaSeq 6000 platform (Illumina) aiming for a minimum of 50,000 reads/cell. In parallel, somatic mutations were assessed from the same BM MNCs using a 98-gene myeloid NGS panel, as previously described^46^.

### scRNA-seq data processing, filtering, batch effect correction, and clustering

Raw sequencing reads from single cells were aligned against the customized human reference genome and processed using the *count* module of Cell Ranger. Gene-barcode matrices containing only barcodes with UMI counts passing threshold were imported to Seurat V5 for further analysis. Genes detected in fewer than 3 cells were excluded; cells with less than 200 genes detected or greater than 10% mitochondrial UMIs were also filtered out. Doublets were detected using Scrublet, DoubletFinder, scDblFinder, and doubletCells implemented in scran, assuming 0.08% doublet rate for every 1,000 sequenced cells. Cells identified as doublets by at least two algorithms were removed from further analysis. Raw UMI counts were log normalized and the top 5,000 variable genes were identified using the “vst” method implemented in the *FindVariableFeatures* function in Seurat. T cell receptor and immunoglobulin genes were removed from the variable genes to prevent clustering based on V(D)J transcripts. S and G2/M cell cycle phase scores were assigned to cells using the *CellCycleScoring* function.

The Seurat layer data were further scaled using the *ScaleData* function by regressing against total read count, percentages of mitochondrial UMIs, and cell cycle phase scores (S and G2/M) and principal component analysis was performed on the combined dataset to obtain the top 40 principal components. To further remove batch effects, the *HarmonyIntegration* algorithm in Seurat was applied to correct for batch effects, and the resulting corrected embeddings were used for downstream analyses. A shared nearest neighbor (SNN) based graph was constructed using the top 40 Harmony principal components, and clusters were identified using the Louvain algorithm with *FindCluster* function at resolution = 1 (33 clusters). Uniform Manifold Approximation and Projection (UMAP) were generated by the *RunUMAP* function and used for visualization.

### Human BM cluster annotation

Human bone marrow (BM) scRNA-seq data were analyzed as described above. A shared nearest neighbor (SNN) based graph was constructed using the top 40 Harmony principal components, and clusters were identified using the Louvain algorithm using *FindCluster* function at resolution = 1 (22 cell types). Clusters were annotated by comparing the cluster-specific genes to canonical markers for hematopoietic stem cells (HSCs) (*SPINK2, MEIS1, MLLT3*), erythroblasts (*AHSP, HBB, ALAS2*), macrophages (*CD68, CD163*), neutrophils (*CXCL8, CXCR2*), megakaryocytes (*PPBP, PF4, GP9*), common myeloid progenitors (CMPs) (*FLT3, CEBPA*), CD14+ monocytes (*CSF3, CD36, S100A12*), CD16+ monocytes (*LST1, LILRB2*), multi-lymphoid progenitors (MLPs) (*DNTT, CXCR4*), fibroblasts (*COL6A1, COL1A2, DCN*), granulocyte-monocyte progenitors (GMPs) (*MPO, KIT, CD34*), mast cells (*KIT, TPSAB1, CPA3*), megakaryo-erythroid progenitors (MEPs) (*GATA1, HBD, TFRC*), T-cells (*CD3E, CD3D*), CD4+ T-cells (*CD4*), CD8+ T-cells (*CD8A, CD8B*), NK cells (*NCAM1*, *KLRB1*, *KLRD1)*, Naive B-cells (*IL4R, IGHD*), Pro/Pre-B cells (*SOX4, RAG2, LEF1*), Plasma cells (*IGHA1, IGHG1, IGHG2, IGHG4*), plasmacytoid dendritic cells (pDCs) (*LILRA4, IL3RA*), and conventional dendritic cells (cDC) (*CST3, CD1A*).

### Numbat analysis

To infer large-scale CNVs from scRNA-seq data, the Numbat algorithm (v1.3.2.1)^36^ was applied to the 10x Genomics scRNA-seq dataset. Numbat integrates gene expression, allele-specific, and haplotype information to comprehensively characterize CNV profiles at the single-cell level. We followed the guidelines provided in the Numbat GitHub documentation (https://kharchenkolab.github.io/numbat/articles/numbat.html). Allele-specific data was obtained using the pileup_and_phase.R script with default parameters. CNV profiles were then inferred using the run_numbat function with default settings. Samples collected from different conditions of the same patient were analyzed and visualized jointly, with major cell type annotations and genotype information incorporated into the CNV visualization.

### Differential gene expression comparing 8+ or 1+ vs disomy mononuclear cells

To assess transcriptional differences associated with CNV, we compared cells with copy number gains on chr8 or chr1 with disomy MNCs within each major cell type. Cell classification was based on copy number states inferred by Numbat. Cells with an inferred CNV value ≥1 were classified as CNV-gain cells, while all remaining cells were considered disomy MNCs. Differential gene expression analysis between CNV-gain and disomy groups was performed using the *FindMarkers* function in the Seurat package. Across all samples, a total of 24,942 8+ cells and 46,148 disomy 8 cells were included for chr8 analysis, while 2,372 1+ cells and 23,560 disomy 1 cells were included for chr1 analysis.

### InferCNV analysis

To further characterize CNVs in 8+ or 1+ cells compared with disomy MNCs, CNV scores were calculated for each cell using the inferCNV R package (v1.21.0) (GitHub). A raw count matrix, cell annotation file, and gene–chromosome position file were prepared in accordance with the inferCNV data requirements (https://github.com/broadinstitute/inferCNV). For visualization and comparison of cells with copy number gains and disomy cells, all cells were ordered by groups defined by the combination of trisomy status and patient identity. To avoid clustering driven by group-level, the parameter k_obs_groups was set to 1, and three HD samples were selected as reference cells for CNV inference.

### Immunohistochemistry staining

Paraffin-embedded BM trephine biopsies were used for immunohistochemistry (IHC) analyses as described^47^. Tissue blocks were sectioned at 4-µm thickness. Unstained slides were deparaffinized using an automated system with EZ Prep solution (Ventana Medical Systems) and stained with anti-MYC antibody using a Ventana Discovery XT automated system (Ventana Medical Systems) following the manufacturer’s protocols. The percentage of MYC-positive cells was assessed by an independent hematopathologist blinded to clinical data.

### Animal studies

All animal studies were performed in compliance with the National Institutes of Health Guidelines under a protocol approved by the H. Lee Moffitt Cancer Center & Research Institute and the University of South Florida Institutional Animal Care and Use Committee (IACUC). Mouse genotypes from tail biopsies were determined using real time PCR with specific probes designed for each gene (Transnetyx). All mice used in our *in vivo* studies were on a C57BL/6J background.

Mx1-Cre^+/-^;Rosa26^LSL-MYC/LSL-MYC^ mice were generated by crossing Mx1-Cre^+/-^ mice (RRID:IMSR_JAX:003556) with Rosa26^LSL-MYC/LSL-MYC^ mice (RRID:IMSR_JAX:033805). Mice at ages 6-11 weeks in both experimental and control groups were treated with pIpC (250 µg/kg, D1 and D3, prepared in normal saline, i.p.) to induce *MYC* gene expression. S100A9Tg mice (RRID:IMSR_JAX:018055) were obtained from the Jackson Laboratory and backcrossed to a congenic C57BL/6J background. Using peripheral blood (PB) samples, complete blood counts (CBC) with differential were assessed 1 week prior to pIpC treatment (baseline), and then every 4 weeks following pIpC treatment. Mice at pre-defined endpoints were sacrificed and primary tissues were analyzed via the assays described below. Pre-defined endpoints used in our *in vivo* studies include (i) substantial weight loss (≥20% loss), (ii) abnormalities with movement or breathing, (iii) excessive lethargy, tremors, restlessness, failure to groom causing unkempt appearance, teeth grinding, or guarding (protecting the painful area), (iv) failure to show normal patterns of inquisitiveness, and (v) inability to urinate or defecate. Mice were harvested at pre-defined endpoints and primary BM and spleen cells were harvested for RNA extraction and qRT-PCR and for immunoblotting of proteins. BM, spleen, liver, heart, lung, and colon tissues were also harvested and incubated in Neutral Buffered Formalin for 24 hours, then stored in 70% EtOH until used for trichrome staining.

### Immunoblotting

Primary cells isolated from BM and spleen (5×10⁶ cells per sample) were washed with ice-cold PBS and lysed in RIPA buffer (10 mM Tris [pH 7.4], 100 mM NaCl, 1 mM EDTA, 1 mM EGTA, 1% Triton X-100, 10% glycerol, 0.1% SDS, 0.5% deoxycholate) supplemented with protease inhibitor cocktail (complete mini tablet, 1 tablet/10 ml), 1 mM PMSF, 10 mM β-glycerophosphate, 1 mM sodium fluoride, and 1 mM sodium orthovanadate. Lysates were sonicated, cleared by centrifugation (15,000 rpm, 2 min), and protein concentrations were quantified by BCA assay. Equal amounts of protein were resolved by SDS-PAGE, transferred to nitrocellulose membranes, and probed with primary antibodies. Blots were visualized using the Odyssey Fc Imaging System (LI-COR).

### qRT-PCR assays

Total RNA was extracted from primary BM and spleen cells using the RNA extraction kit (Qiagen). Reverse transcription was performed with the iScript cDNA synthesis kit, followed by quantitative real-time PCR on a CFX96 Touch Real-Time PCR Detection System (BioRad). Relative gene expression was calculated using the comparative Ct method (2^-ΔΔCt), with Ubiquitin and Actin as reference genes.

### Flow cytometry analyses

Primary BM and spleen cells were harvested as above. Following RBC lysis using ACK buffer, cells were resuspended in PBS with 2% FBS. To characterize changes in individual hematopoietic lineages, cells were stained with Zombie near IR (ZNIR, viability dye), mouse anti-CD16/32 (Fc block), -Ter119-V450, -B220-PE, -Ly-6G/Ly-6C-APC, -CD11b-BUV737, - CD3-BV786, and -F4/80-BUV395 fluorochrome-conjugated antibodies. To characterize the HSCs and progenitor populations, cells were stained with mouse Lin cocktail (BV421), anti-CD34-PE, -CD117-APC, -Ly-6A/Ly-6E-BB515, -CD150-PE-Dazzle 594, -CD48-Brilliant Violet 711, and -CD16/CD32-BUV395 conjugated antibodies. Cells were fixed with 4% paraformaldehyde for 10 minutes, washed with FACS buffer three times, and then subjected to flow cytometry.

### Reticulin and Trichrome staining

Reticulin and trichrome staining were performed to assess fibrosis in tissues. The reticulin stain was based on the method of Gomori and Snook (Artisan Reticulin-Nuclear Fast Red Stain Kit), and the trichrome stain was based on the original Masson’s procedure (Artisan Masson’s Trichrome Stain Kit). Staining was performed using the Artisan Staining System following protocols provided by the manufacturer.

## Data Availability

The mCA and CHIP calls used in our study are available from UK Biobank, through an application process described at http://www.ukbiobank.ac.uk/using-the-resource. Deidentified clinical and genetic data supporting the findings of this study are available from the corresponding authors upon reasonable request, subject to completion of a data sharing agreement and compliance with applicable ethical and privacy regulations.

## DATA AVAILABILITY

The mCA and CHIP calls used in our study are available from UK Biobank, through an application process described at http://www.ukbiobank.ac.uk/using-the-resource. De-identified clinical and genetic data supporting the findings of this study are available from the corresponding authors upon reasonable request, subject to completion of a data sharing agreement and compliance with applicable ethical and privacy regulations.

## CODE AVAILABILITY

Codes used in this study are available on the Yun-Lab GitHub page (https://github.com/namaska97/Yun-Lab).

## AUTHOR CONTRIBUTIONS

Study conception and design: N.D.V. and S.Y.; performed experiments, collected, and assembled the data: N.D.V., C-H.C., A.T.K., F.M.L., T.N.R., J.M., L.Z., J.P., P.R.P., X.Y., S.Y.; analyzed and interpreted the data: N.D.V., Q.M., C-H.C., J.L.C., X.Y., and S.Y.; writing and/or revision of the manuscript: N.D.V., Q.M., J.L.C., X.Y., and S.Y.; review of manuscript: all authors reviewed the manuscript; administrative, technical or material support: N.D.V., D.J.M., J.L.C., and S.Y.; study supervision: N.D.V. and S.Y.

## ACKNOWLEDGEMENTS

The authors thank Jodi Kroeger, Dr. Neel Chaudhary, and the Moffitt Flow Cytometry Core; Sean Yoder, Dr. Chaomei Zhang, Dr. Lan Zhang, Tania Mesa, and the Moffitt Genomics Core for their help with scRNA-seq analyses; Samara Rivera-Santiago, Yamila Caraballo Perez, Janis De La Iglesia, and the Moffitt Clinical Pathology Lab for their help with trichrome staining; and Dr. Dae-hyun Lee for technical support with UK Biobank access application. This research has been conducted using the UK Biobank Resource under Application Number 92513. This work was supported in part by Deutsche Krebshilfe grant 109220 (D.J.M.), the Cortner-Couch Endowed Chair for Cancer Research from the University of South Florida School of Medicine (J.L.C.), NIH grant K08 CA237627 (S.Y.), R01 CA280116 (S.Y.), K22 CA292451 (N.D.V.), an American Society of Hematology (ASH) Research Training Award for Fellows (RTAF) (S.Y.), an ASH Scholar Award for Fellows (S.Y.), an ASH Scholar Award for Fellow to Faculty (N.D.V.), a Career Development Award from the American Society of Clinical Oncology (ASCO) (S.Y.), a Miles for Moffitt grant from the H. Lee Moffitt Cancer Center and Research Institute (S.Y. and Q.M.), a research grant from the Clinical Science Division at the H. Lee Moffitt Cancer Center & Research Institute (S.Y.), a research fund from the Southeast Trust (S.Y.), and by NCI Comprehensive Cancer Grant P30 CA076292 to the H. Lee Moffitt Cancer Center & Research Institute.

## Supplementary Information

**Supplementary Information includes:**

Supplementary Figures S1-S19

Supplementary Table S1-S14

**Table S1:** Baseline demographic and laboratory profiles in UKBB cohort.

| **Variables** | **mCA**<br>(n=42,390) | **No mCA**<br>(n=410,204) | **Total**<br>(n=452,594) |
| --- | --- | --- | --- |
| **Sex (%)** |  |  |  |
| Female | 14,659 (34.6) | 240,220 (58.6) | 254,879 (56.3) |
| Male | 27,731 (65.4) | 169,984 (41.4) | 197,715 (43.7) |
| **Age, median (IQR)** |  |  |  |
| At recruitment | 63 (59-66) | 57 (49-62) | 57 (50-63) |
| At mCA assessment | 63 (59-67) | 57 (50-63) | 58 (50-63) |
| **Follow up, median (IQR)** | 14.7 (13.9-15.5) | 14.9 (14.1-15.6) | 14.9 (14.1-15.6) |
| **Lifestyle (%)** |  |  |  |
| Smoking history | 23,427 (55.3) | 177,694 (43.3) | 210,121 (44.4) |
| Alcohol use | 40,889 (96.5) | 390,150 (95.1) | 431,039 (95.2) |
| **CBC, median (IQR)** |  |  |  |
| WBC, cells/μL | 6.9 (5.9-8.2) | 6.6 (5.6-7.8) | 6.6 (5.6-7.8) |
| Neutrophil, cells/μL | 4.2 (3.4-5.1) | 4.0 (3.2-4.9) | 4.0 (3.3-5.0) |
| Hemoglobin, g/dL | 14.5 (13.6-15.3) | 14.1 (13.3-15.0) | 14.1 (13.3-15.0) |
| Platelet, K/μL | 242.5 (208.2-282.0) | 249.1 (214.5-288.1) | 248.7 (214.0-287.9) |

**Abbreviations:** mCA (mosaic chromosomal alteration), IQR (interquartile range), CBC (complete blood count), WBC (white blood cells)

**Table S2:** Incident disease risk by ICD-10 chapter based on mCA status.

**Table S3:** Incident disease risk by ICD-10 chapter based on individual mCA subtype.

**Table S4:** Incident inflammaging-related disease risk by organ system based on mCA status.

**Table S5:** Incident inflammaging-related disease risk by organ system based on individual mCA subtype.

**Table S6:** Dose-response analysis of incident disease risk by MCF for individual mCA subtypes.

**Table S7:** All-cause mortality risk by mCA subtype.

**Table S8:** Cause-specific mortality risk by ICD-10 chapter based on mCA status.

**Table S9:** Incident disease risk by ICD-10 chapter based on mCA status excluding CHIP participants.

**Table S10:** All-cause mortality risk by mCA subtype excluding CHIP participants.

**Table S11:** Cytoband-level disease associations identified by LASSO-Cox regression with HR estimates.

**Table S12:** Gene annotations for disease-associated cytobands.

**Table S13:** Differentially expressed genes in 8+ HSCs compared to healthy donor HSCs.

**Table S14:** Differentially expressed genes in 1+ HSCs compared to healthy donor HSCs.

**Supplementary Fig. 1.**
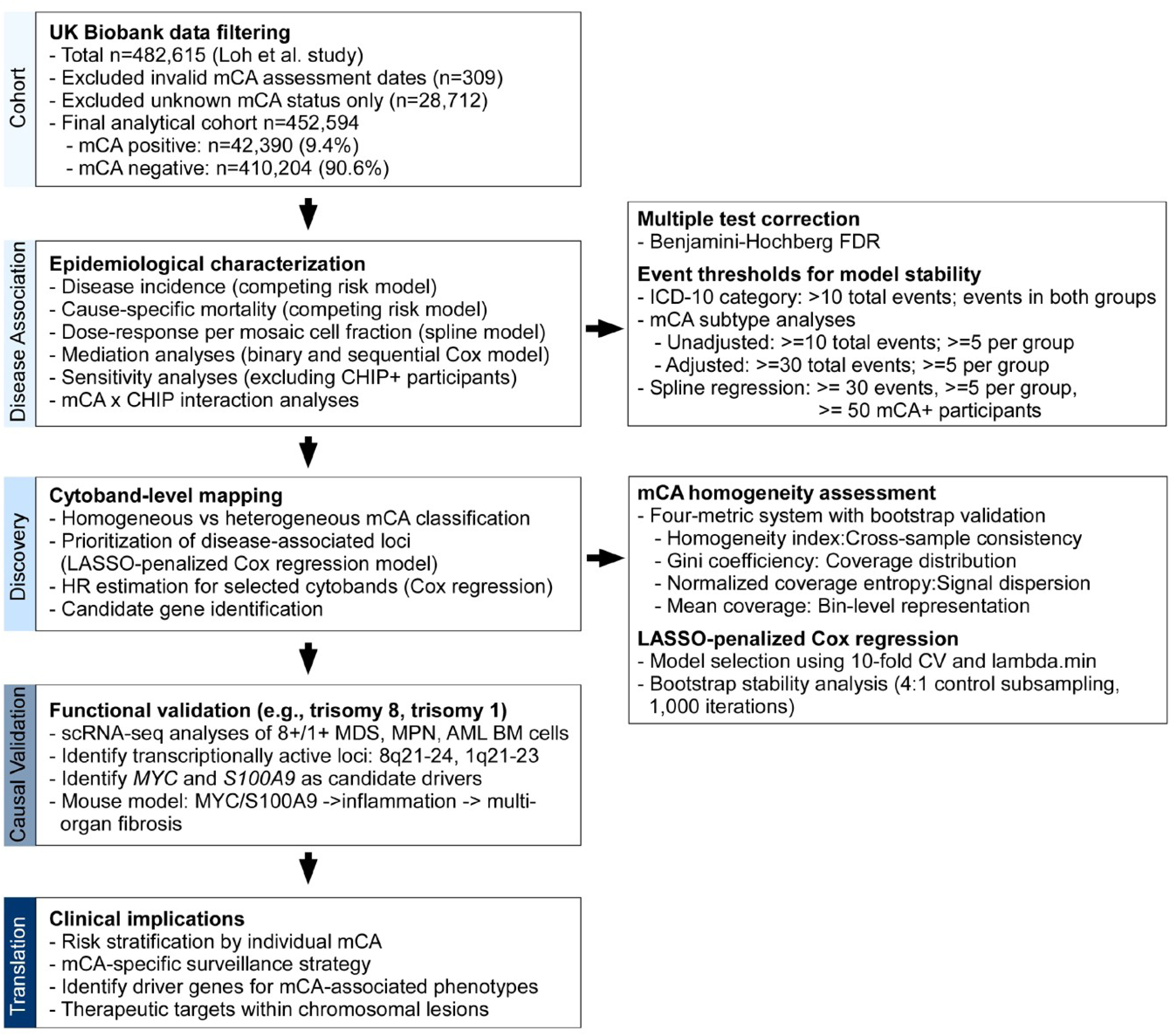
Study design and analytical workflow. Schematic overview of the analytical pipeline. mCA calls were obtained from Loh et al.^20^; somatic mutation (CHIP) data were obtained from the Vlasschaert et al. study^45^. Left panels show the sequential stages of analysis: (i) UK Biobank cohort filtering and mCA classification, (ii) epidemiological characterization of mCA-disease associations, (iii) cytoband-level mapping to prioritize disease-associated loci and to identify candidate genes, (iv) experimental interrogation of selected candidate genes using scRNA-seq and mouse models, and (v) clinical implications. Right panels detail the statistical approaches used at each stage, including multiple testing correction, event thresholds for model stability, mCA homogeneity assessment metrics, and LASSO-penalized Cox regression parameters.

**Supplementary Fig. 2.**
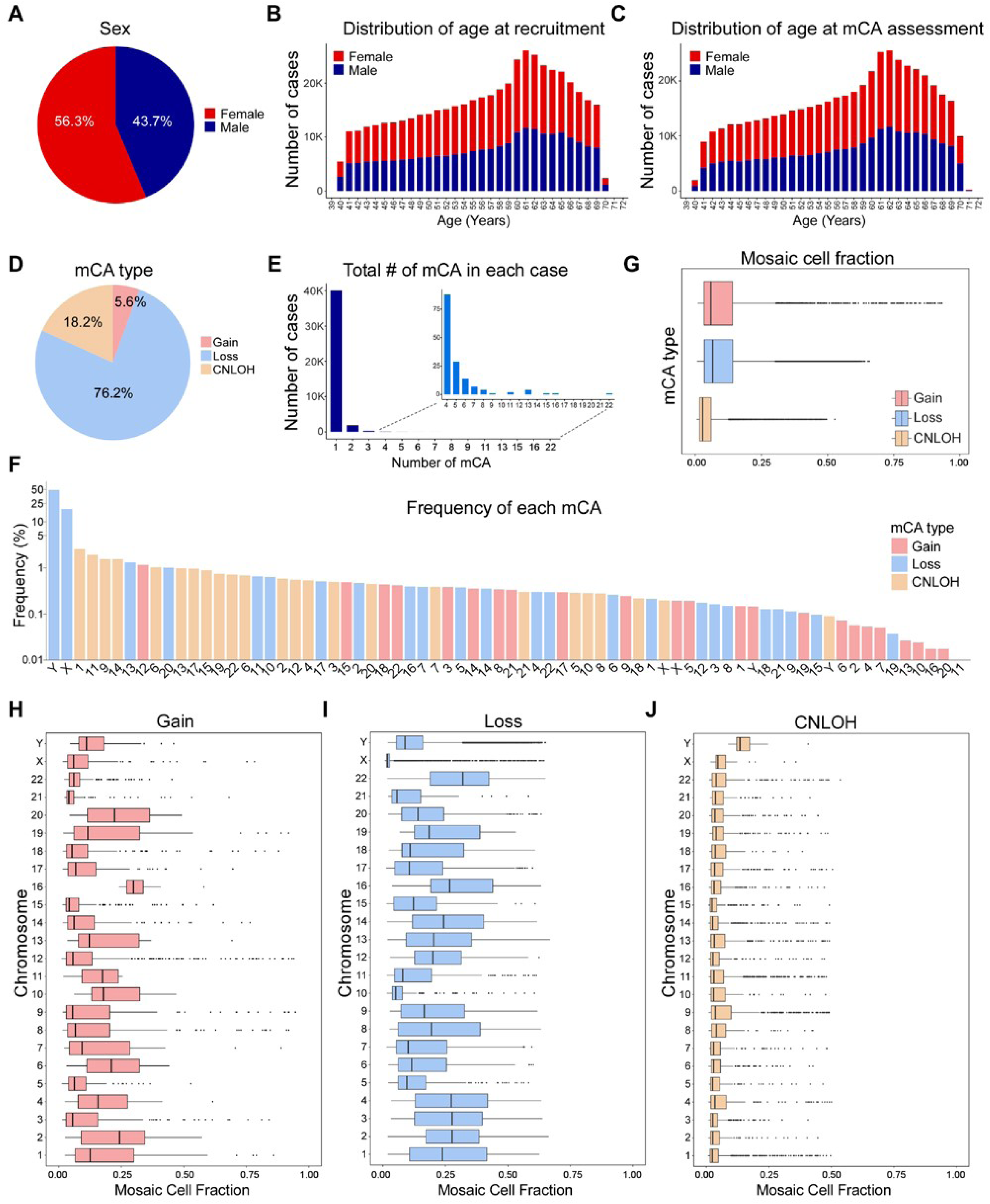
Demographic and mCA characteristics of the UK Biobank cohort. **a**, Sex distribution of the study cohort (n=452,594). **b-c**, Age distribution at recruitment (**b**) and at mCA assessment (**c**) by sex (red, female; blue, male). **d**, Distribution of mCA types among all detected events (n=45,536): loss (n=34,686; 76.2%), CNLOH (n=8,309; 18.2%), and gain (n=2,541; 5.6%). **e**, Number of mCAs per individual among mCA carriers; inset shows distribution for individuals with ≥4 mCAs. **f**, Frequency of individual mCA subtypes (log scale); colors indicate mCA type (red, gain; blue, loss; orange, CNLOH). **g-j**, Mosaic cell fraction (MCF) distribution by mCA type (**g**) and by chromosome for gain (**h**), loss (**i**), and CNLOH (**j**). Box plots show median and interquartile range.

**Supplementary Fig. 3.**
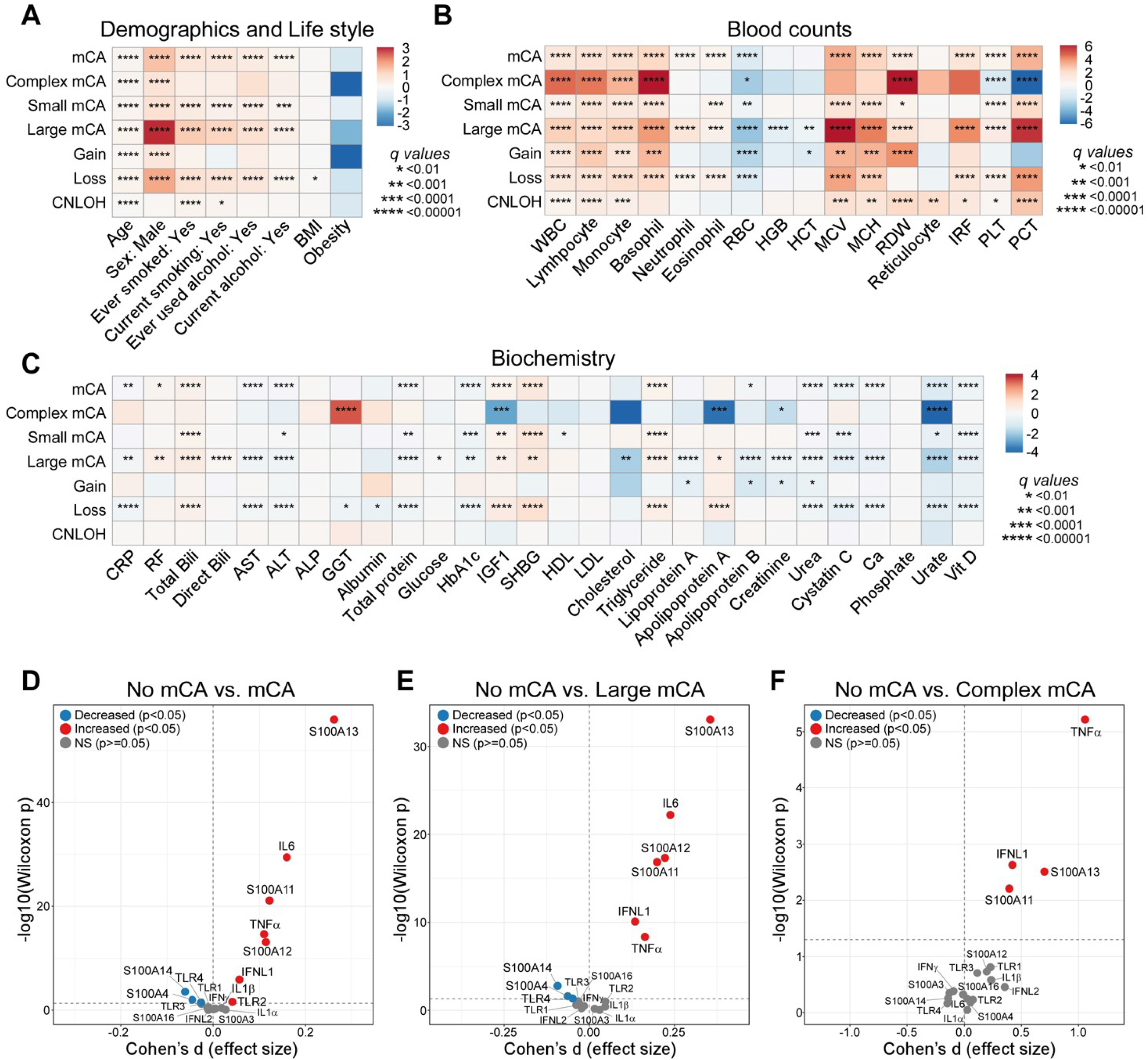
Cross-sectional associations between mCAs and baseline demographic, lifestyle, laboratory, and inflammatory parameters. **a**, Associations between mCA subtypes and demographic and lifestyle factors. **b**, Associations between mCA subtypes and complete blood count parameters. **c**, Associations between mCA subtypes and serum biochemistry parameters. Heatmaps display log₂-transformed odds ratios (ORs) from logistic regression; red indicates positive associations and blue indicates negative associations. For demographic and lifestyle factors, univariate logistic regression was performed. For blood count and biochemistry parameters, models were adjusted for age and sex, with values log-transformed prior to analysis. Rows represent mCA subtypes: all mCA carriers, complex mCA (≥3 chromosomes affected), small mCA (MCF <10%), large mCA (MCF ≥10%), and mCA classified by copy number change (gain, loss, or CNLOH). Multiple testing was corrected using the Benjamini-Hochberg method. **d-f,** Volcano plots showing cross-sectional differences in circulating cytokine and alarmin levels between no mCA controls (n=44,039) and mCA carriers (n=4,678) (**d**), large mCA (n=1,586) (**e**), or complex mCA (n=48) (**f**). Cytokine levels were measured using the Olink platform and expressed as Normalized Protein eXpression (NPX) values. The x-axis shows Cohen’s d effect size, and the y-axis shows −log₁₀(Wilcoxon p-value). Points are colored red (increased in mCA group, p<0.05), blue (decreased in mCA group, p<0.05), or gray (not significant, p≥0.05). Horizontal dashed line indicates p=0.05; vertical dashed line indicates no effect (Cohen’s d=0).

**Supplementary Fig. 4.**
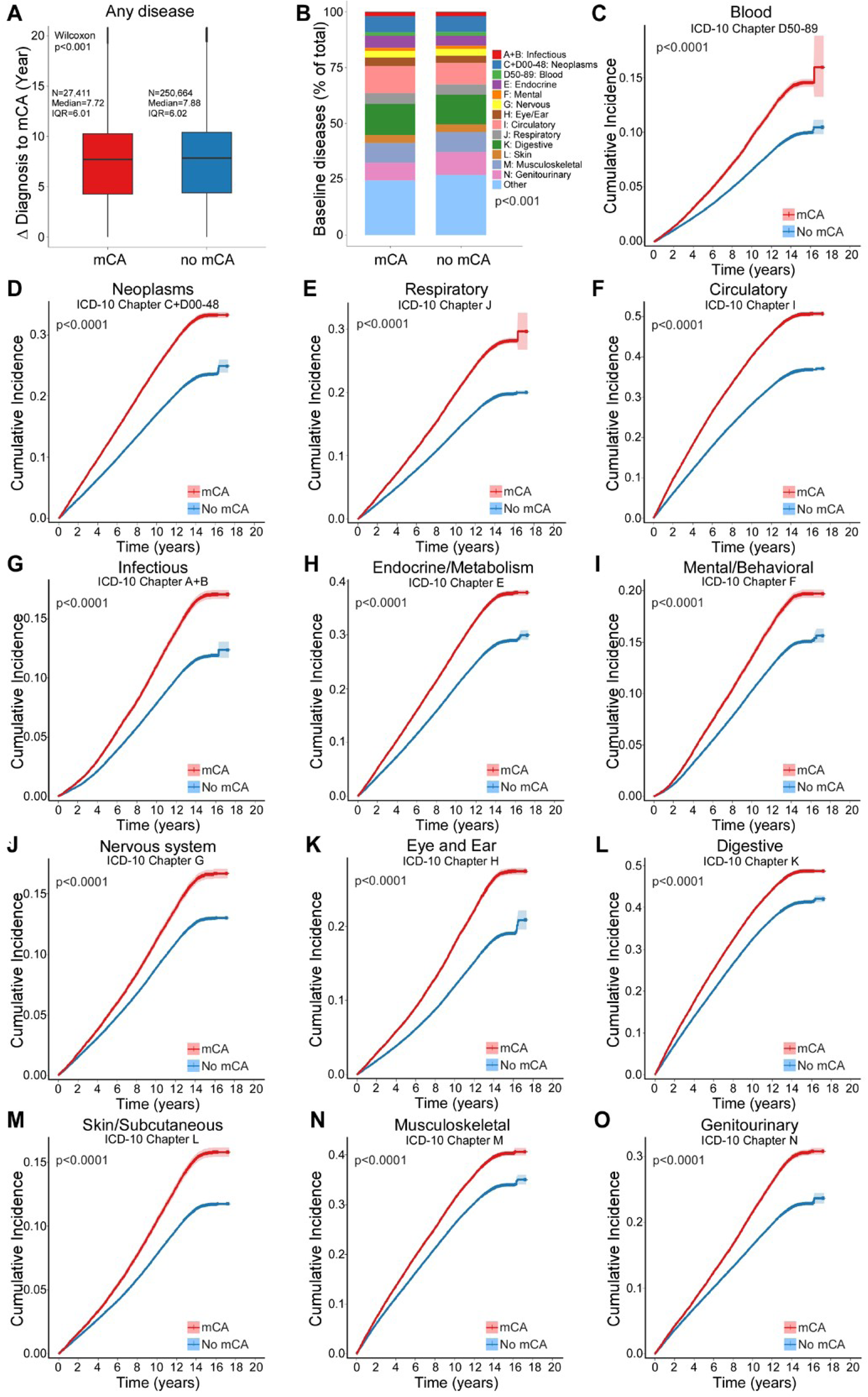
Baseline disease burden and cumulative incidence of diseases by ICD-10 chapter. **a**, Time interval from first disease diagnosis to mCA assessment among participants with at least one baseline diagnosis, comparing mCA carriers (n=27,411) vs. non-carriers (n=250,664). Box plots show median and interquartile range (IQR); P value was calculated using the Wilcoxon rank-sum test. **b**, Distribution of baseline diseases prevalence across ICD-10 chapters in mCA carriers vs. non-carriers; P value calculated using chi-square test. **c-o**, Cumulative incidence of incident diseases by ICD-10 chapter comparing mCA carriers (red) vs. non-carriers (blue): hematologic disorders (**c**), neoplasms (**d**), respiratory (**e**), circulatory (**f**), infectious (**g**), endocrine/metabolic (**h**), mental/behavioral (**i**), nervous system (**j**), eye and ear (**k**), digestive (**l**), skin/subcutaneous (**m**), musculoskeletal (**n**), and genitourinary (**o**). Shaded areas represent 95% CIs; P values were calculated using Gray’s test with death treated as a competing event.

**Supplementary Fig. 5.**
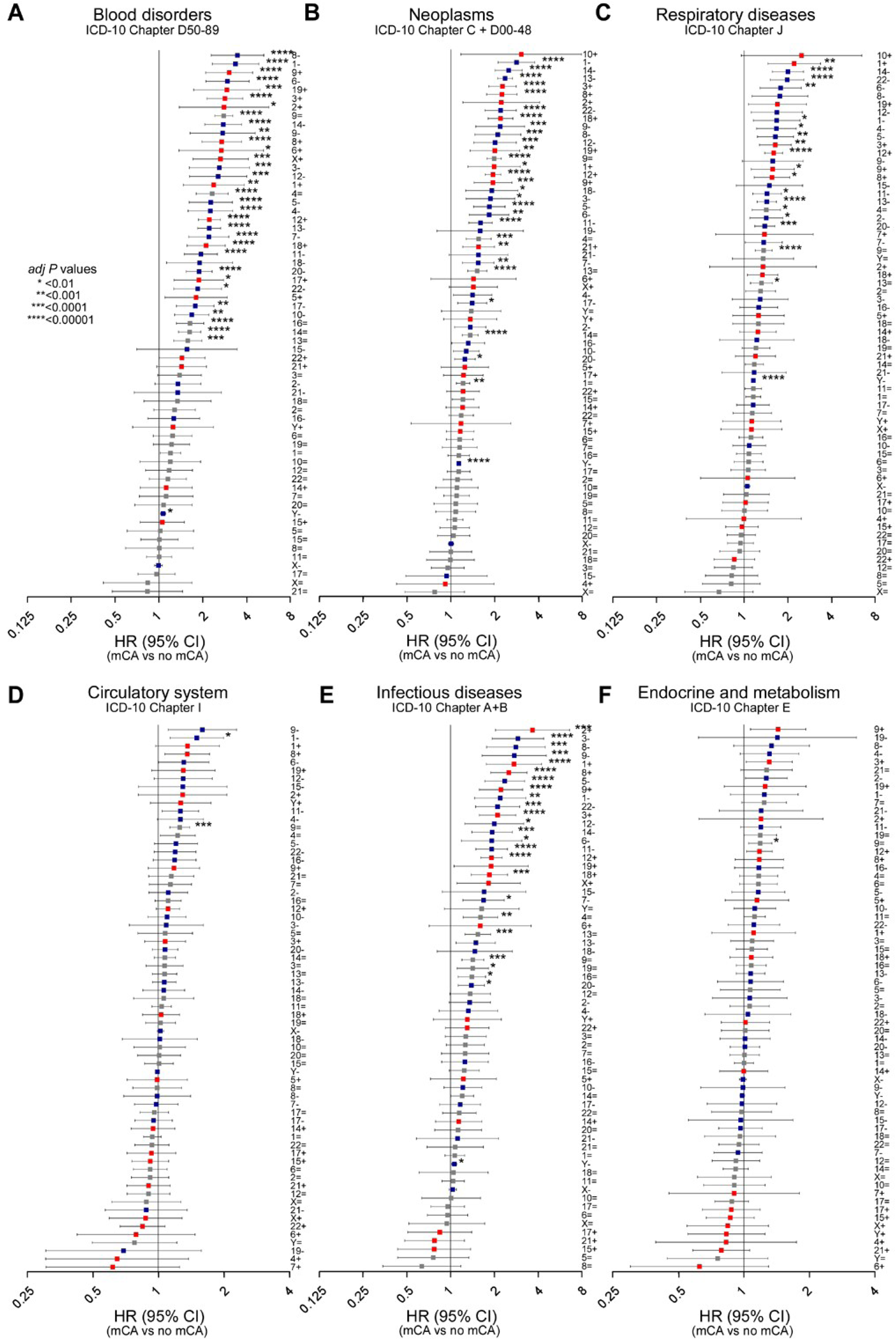
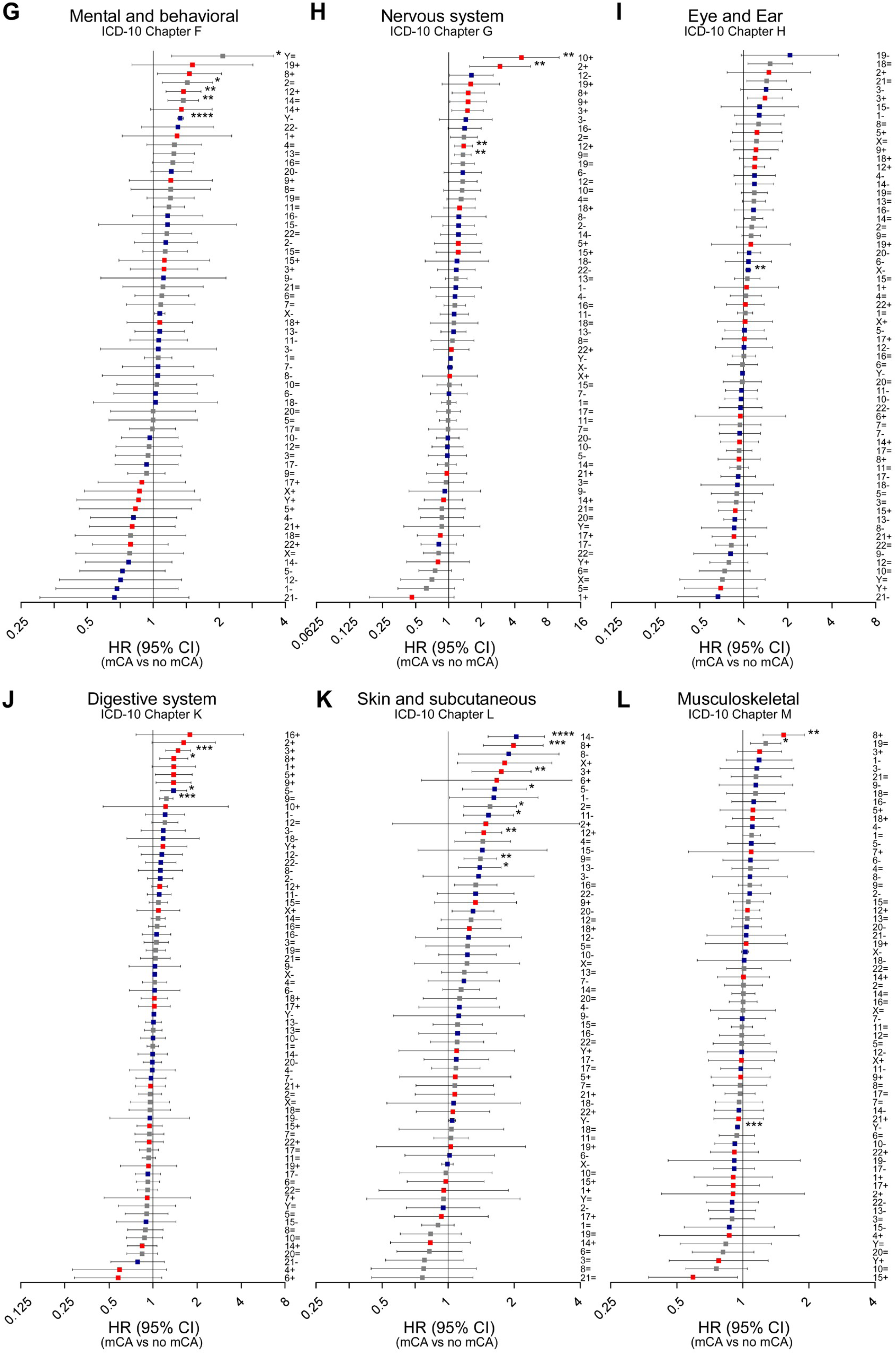
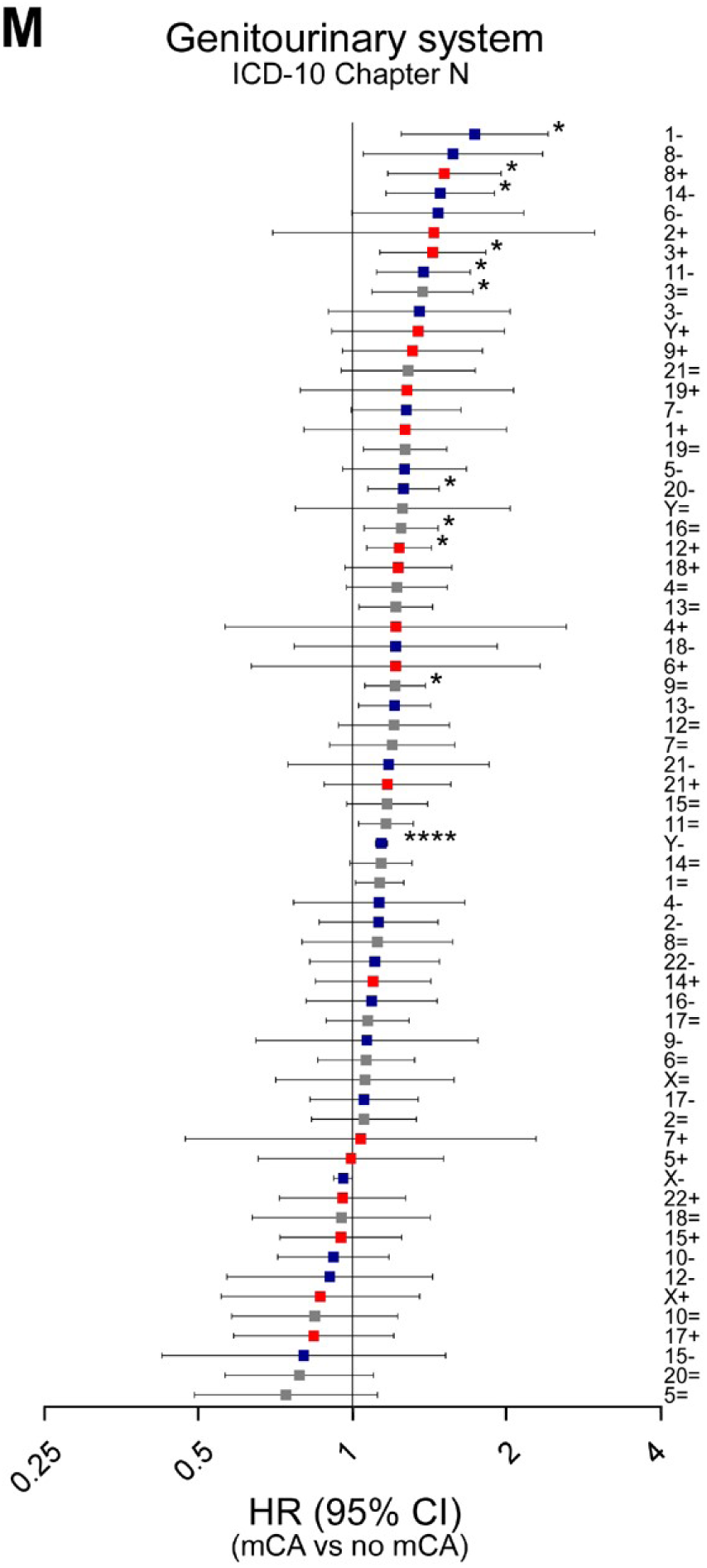
Incident disease risk by individual mCA subtype across ICD-10 chapters. **a-m**, Forest plots showing HRs with 95% CIs for incident disease risk by individual mCA subtype across ICD-10 disease chapters: hematologic disorders (**a**), neoplasms (**b**), respiratory (**c**), circulatory (**d**), infectious (**e**), endocrine/metabolic (**f**), mental/behavioral (**g**), nervous system (**h**), eye and ear (**i**), digestive (**j**), skin/subcutaneous (**k**), musculoskeletal (**l**), and genitourinary (**m**). Colors indicate CNV type (red, gain; blue, loss; gray, CNLOH). Fine-Gray competing risk regression was used with death as a competing event, adjusted for age, sex, and baseline disease burden. Analyses were restricted to mCA-disease combinations with ≥30 total participants, ≥5 events per group. Multiple testing was corrected using the Benjamini-Hochberg method.

**Supplementary Fig. 6.**
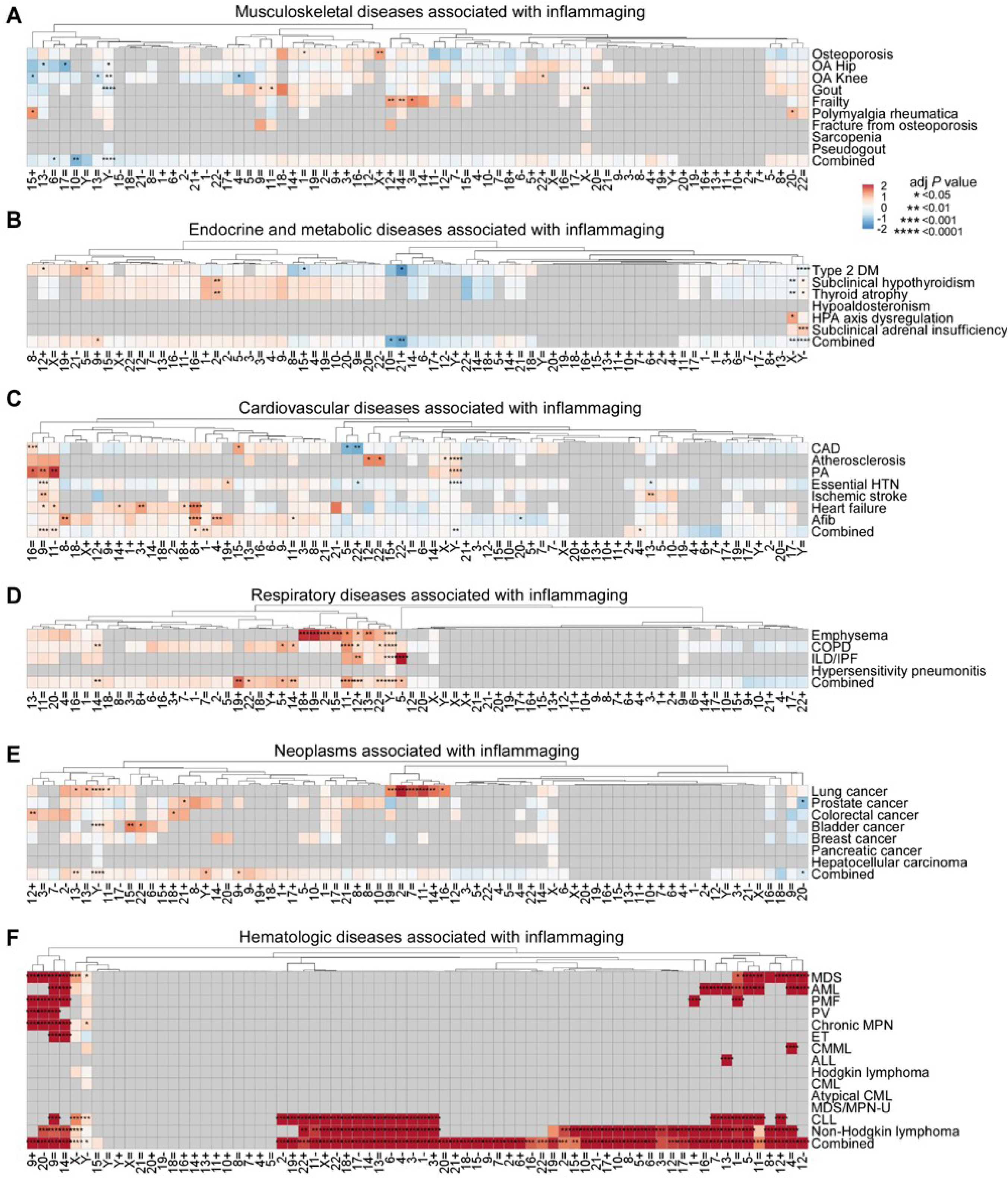
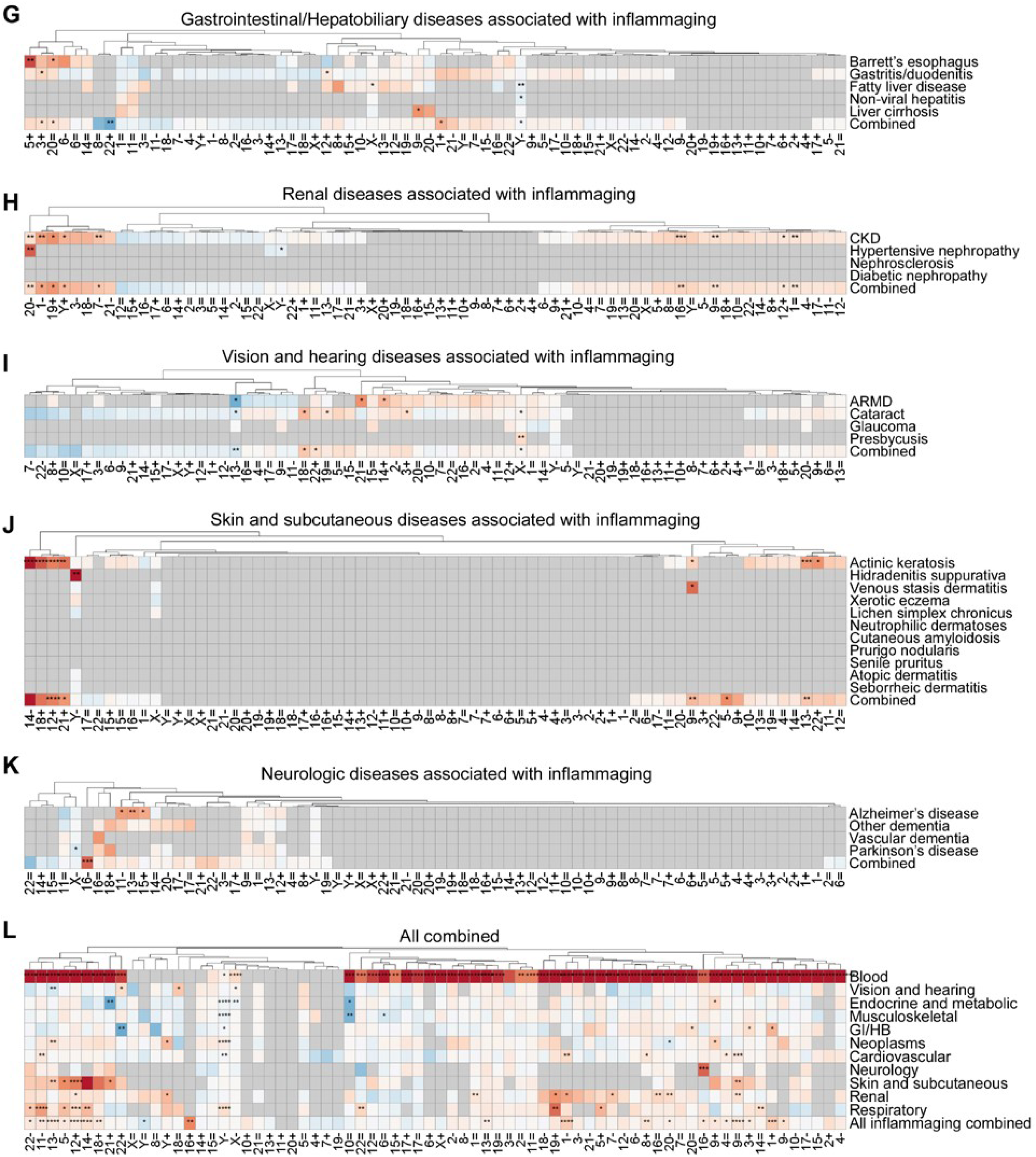
Incident inflammaging-related disease risk by individual mCA subtype. **a-l**, Heatmaps showing log₂-transformed HRs for incident inflammaging-related diseases by mCA subtype across organ system categories: musculoskeletal (**a**), endocrine and metabolic (**b**), cardiovascular (**c**), respiratory (**d**), neoplasms (**e**), hematologic (**f**), gastrointestinal and hepatobiliary (**g**), renal (**h**), vision and hearing (**i**), skin and subcutaneous (**j**), neurologic (**k**), and all categories combined (**l**). Red indicates increased risk (HR>1) and blue indicates decreased risk (HR<1). The “Combined” row at the bottom of each panel represents the category-level combined endpoint. Hierarchical clustering was applied to columns; the Combined row was excluded from row clustering. Fine-Gray competing risk regression was used with death as a competing event, adjusted for age, sex, and baseline disease burden. Analyses were restricted to mCA-disease combinations with ≥30 total events and ≥5 events per group. Multiple testing was corrected using the Benjamini-Hochberg method.

**Supplementary Fig. 7.**
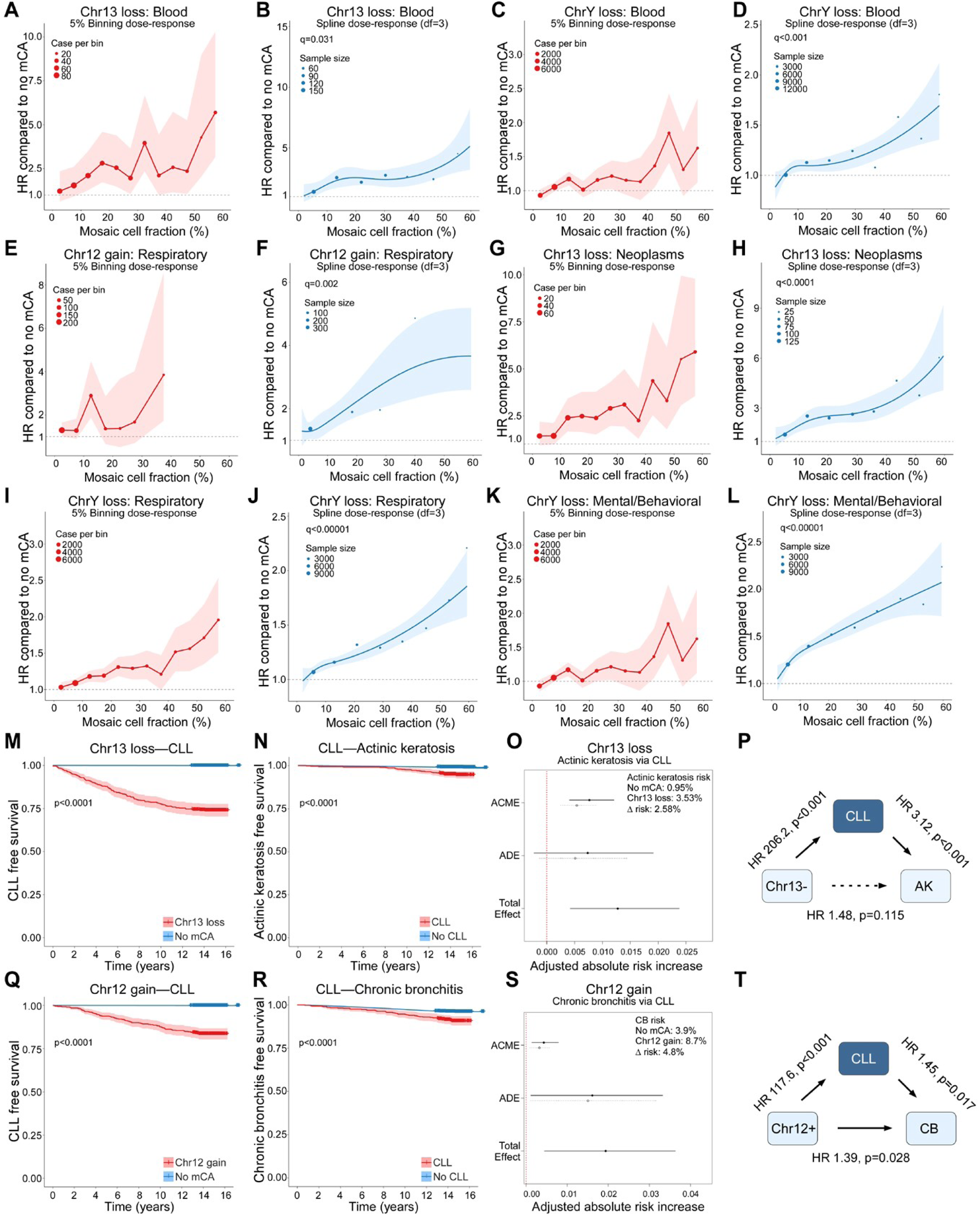
Additional dose-response relationships and mediation analyses. **a-l**, Dose-response relationships between MCF and incident disease risk for selected mCA-disease pairs using 5% binning (odd panels) and natural cubic spline regression (df=3) (even panels): 13- and hematologic disorders (**a-b**), Y- and hematologic disorders (**c-d**), 12+ and respiratory disorders (**e-f**), 13- and neoplasms (**g-h**), Y- and respiratory disorders (**i-j**), and Y- and mental/behavioral disorders (**k-l**). In binning plots, dot size reflects sample size per bin; in spline plots, shaded areas represent 95% CIs. Dashed horizontal lines indicate HR=1. **m-p**, Mediation analysis of 13- on actinic keratosis risk via CLL. CLL-free survival comparing 13- carriers vs. non-carriers (**m**) and actinic keratosis-free survival comparing individuals with vs. without CLL (**n**); shaded areas represent 95% CIs and P values were calculated using the log-rank test. Adjusted absolute risk increase showing average causal mediation effect (ACME), average direct effect (ADE), and total effect with 95% CIs (**o**). Path diagram summarizing statistically inferred direct and mediated effects; dashed arrow indicates non-significant direct effect (**p**). **q-t**, Mediation analysis of 12+ on chronic bronchitis risk via CLL, displayed as in m-p; solid arrows indicate significant associations. All models were adjusted for age, sex, and baseline disease burden.

**Supplementary Fig. 8.**
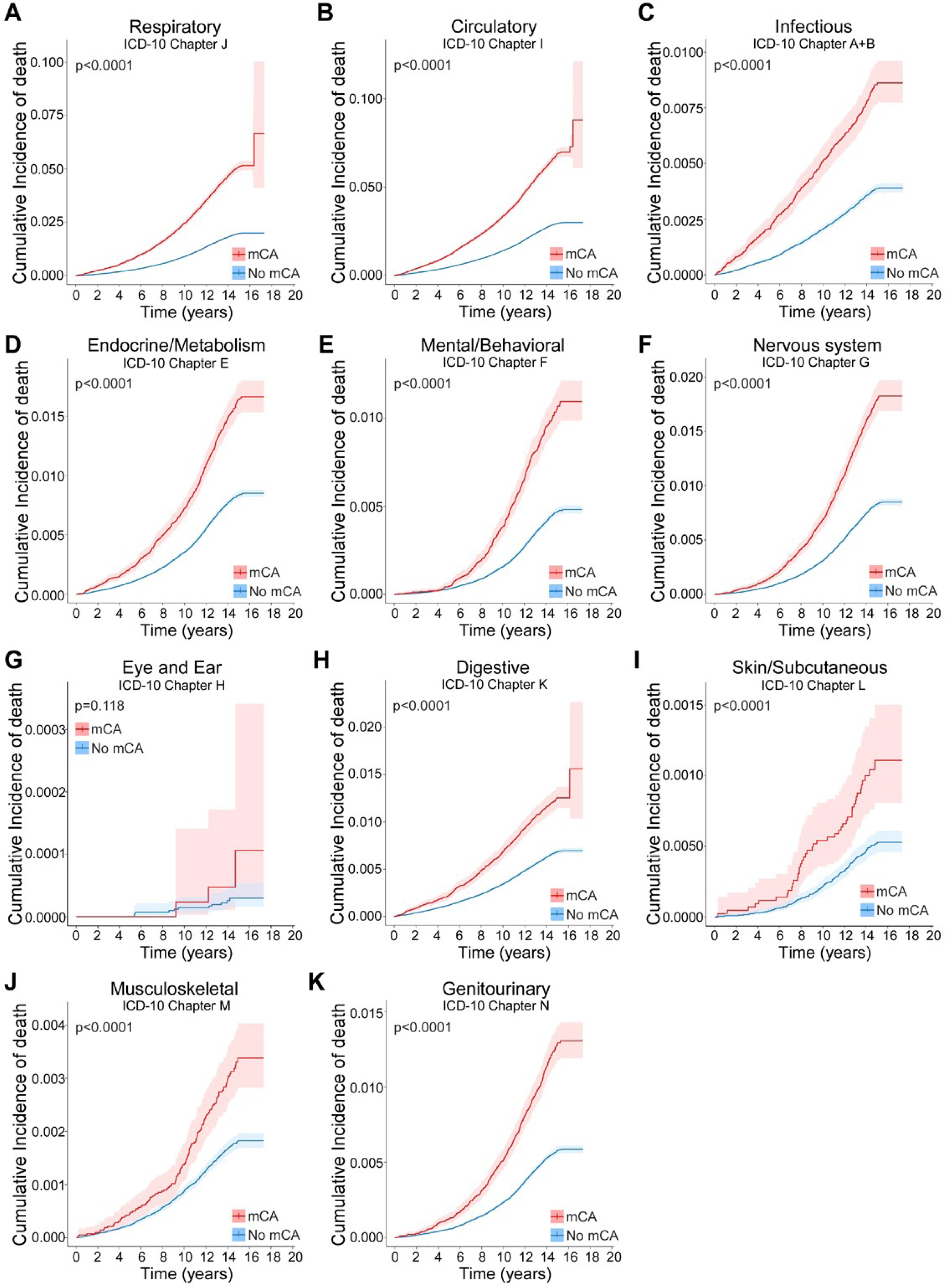
Cause-specific mortality by ICD-10 chapter. **a-k**, Cumulative incidence of death by ICD-10 chapter comparing mCA carriers (red) vs. non-carriers (blue): respiratory (**a**), circulatory (**b**), infectious (**c**), endocrine/metabolic (**d**), mental/behavioral (**e**), nervous system (**f**), eye and ear (**g**), digestive (**h**), skin/subcutaneous (**i**), musculoskeletal (**j**), and genitourinary (**k**). Shaded areas represent 95% CIs; P values were calculated using Gray’s test.

**Supplementary Fig. 9.**
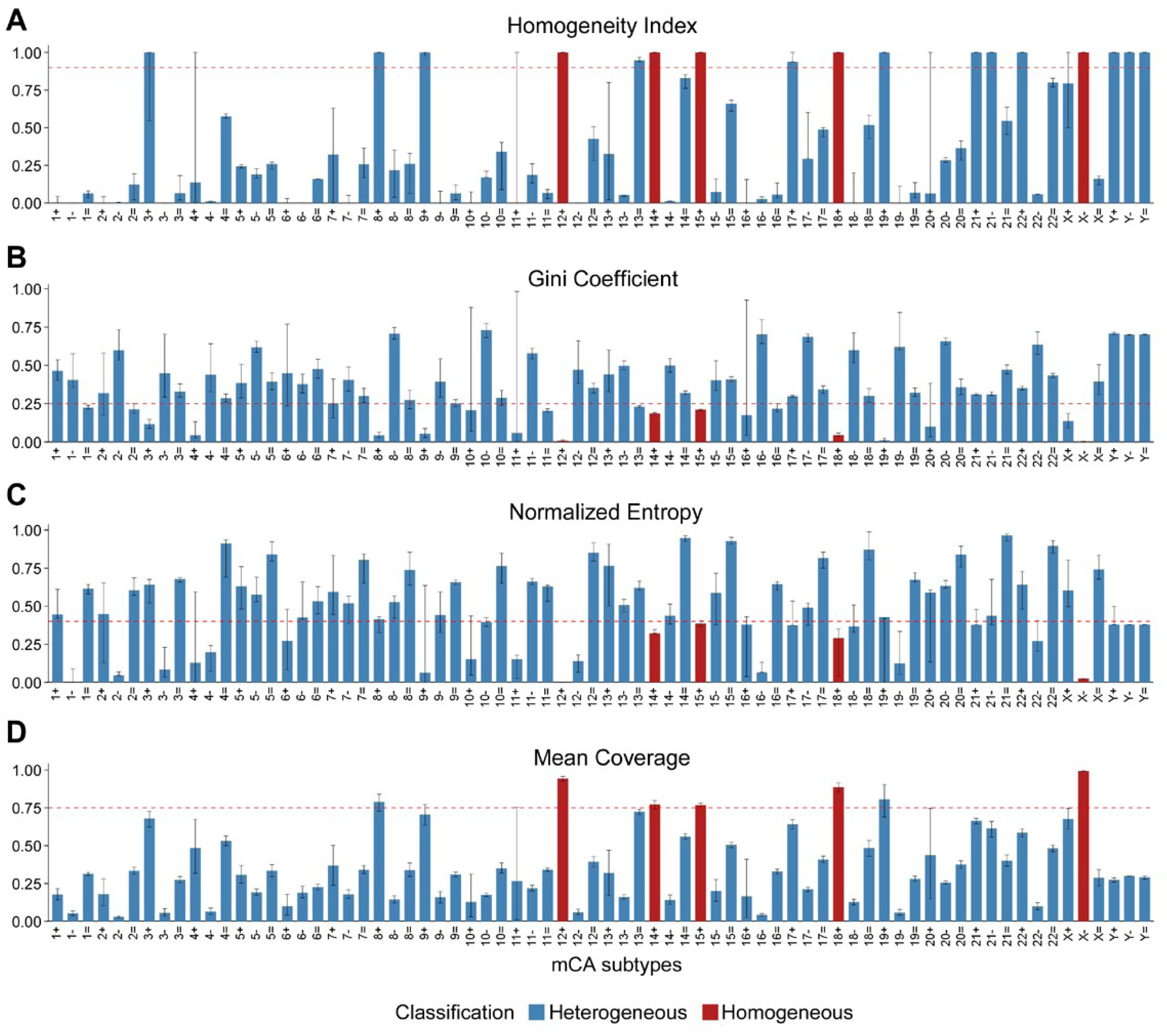
Classification of homogeneous vs. heterogeneous mCAs. **a-d**, Four quantitative metrics used to classify mCA subtypes as homogeneous or heterogeneous: homogeneity index (cross-sample consistency) (**a**), Gini coefficient (coverage distribution) (**b**), normalized coverage entropy (signal dispersion) (**c**), and mean coverage (bin-level representation) (**d**). Red dashed lines indicate pre-defined classification thresholds: homogeneity index >0.9, Gini coefficient <0.25, normalized entropy <0.4, and mean coverage >0.75. All thresholds were pre-specified prior to analysis. mCAs meeting all four criteria were classified as homogeneous (red bars: 12+, 14+, 15+, 18+, X−); all others were classified as heterogeneous (blue bars). Error bars represent 95% CIs derived from bootstrap resampling (1,000 iterations). Homogeneous mCAs represent whole-chromosome or whole-arm events with relatively uniform genomic boundaries across carriers, while heterogeneous mCAs exhibit variable breakpoints amenable to cytoband-level resolution.

**Supplementary Fig. 10.**
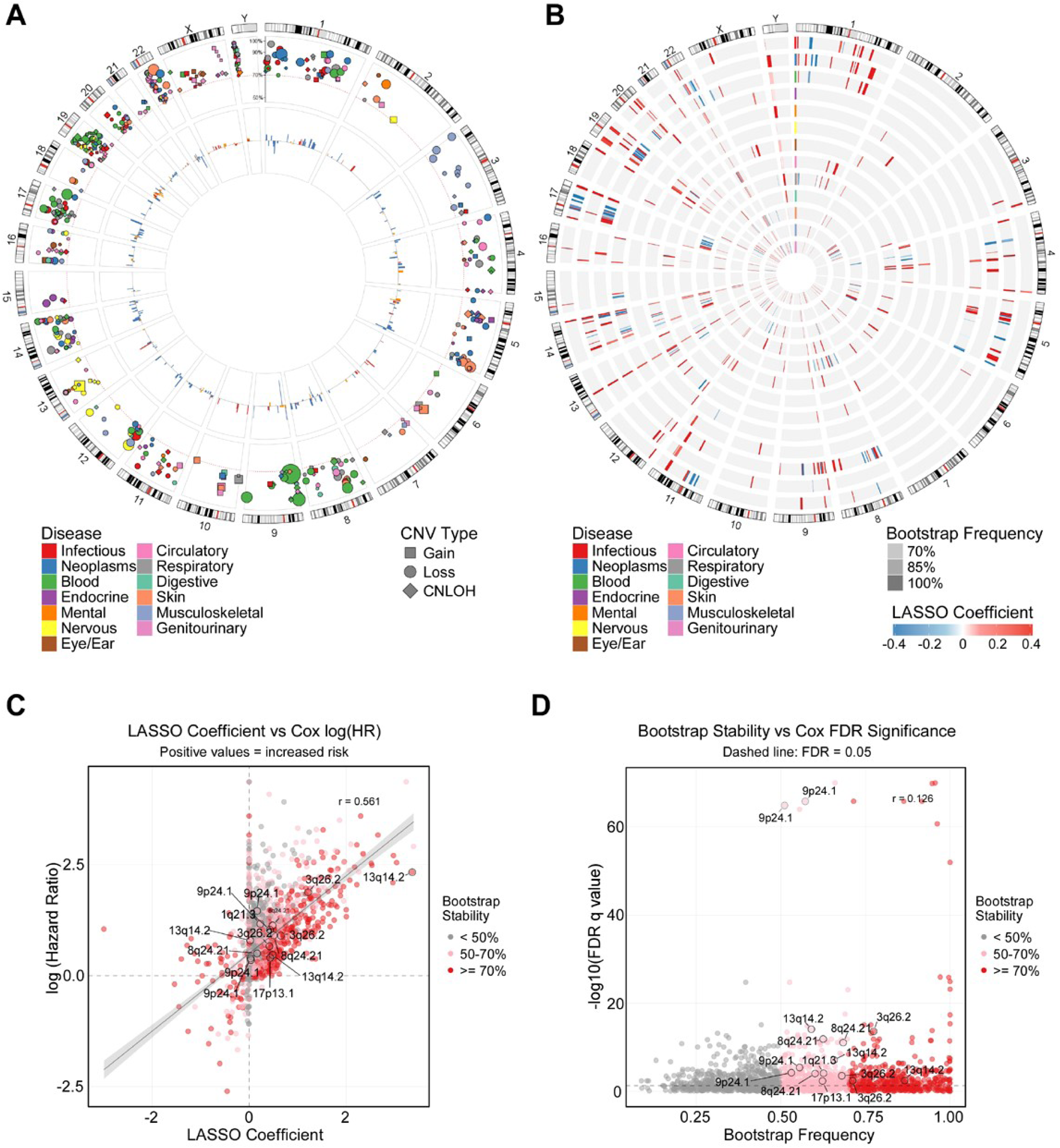
Validation of LASSO-penalized Cox regression via bootstrap stability analysis. **a**, Circos plot displaying cytobands with ≥70% bootstrap selection frequency across disease categories. Point size reflects LASSO coefficient magnitude; shape indicates CNV type; color denotes disease category. **b**, Heatmap circos plot showing the same cytobands, with concentric rings representing disease categories, color indicating coefficient direction (red, increased risk; blue, reduced risk), and opacity reflecting bootstrap frequency. **c**, Scatter plot of LASSO coefficients vs. Cox log(HR), demonstrating positive correlation (r=0.561) and 94% direction concordance. Points are colored by bootstrap stability category (red, ≥70%; pink, 50-70%; gray, <50%). **d**, Bootstrap selection frequency vs. Cox FDR significance (-log10 q-value), illustrating weak correlation (r=0.126) as FDR-significant associations span the full range of bootstrap stability. Features with low bootstrap stability but high FDR significance likely represent sets of correlated cytobands within the same genomic region, for which LASSO selects different cytobands across bootstrap iterations despite shared underlying biological signal.

**Supplementary Fig. 11.**
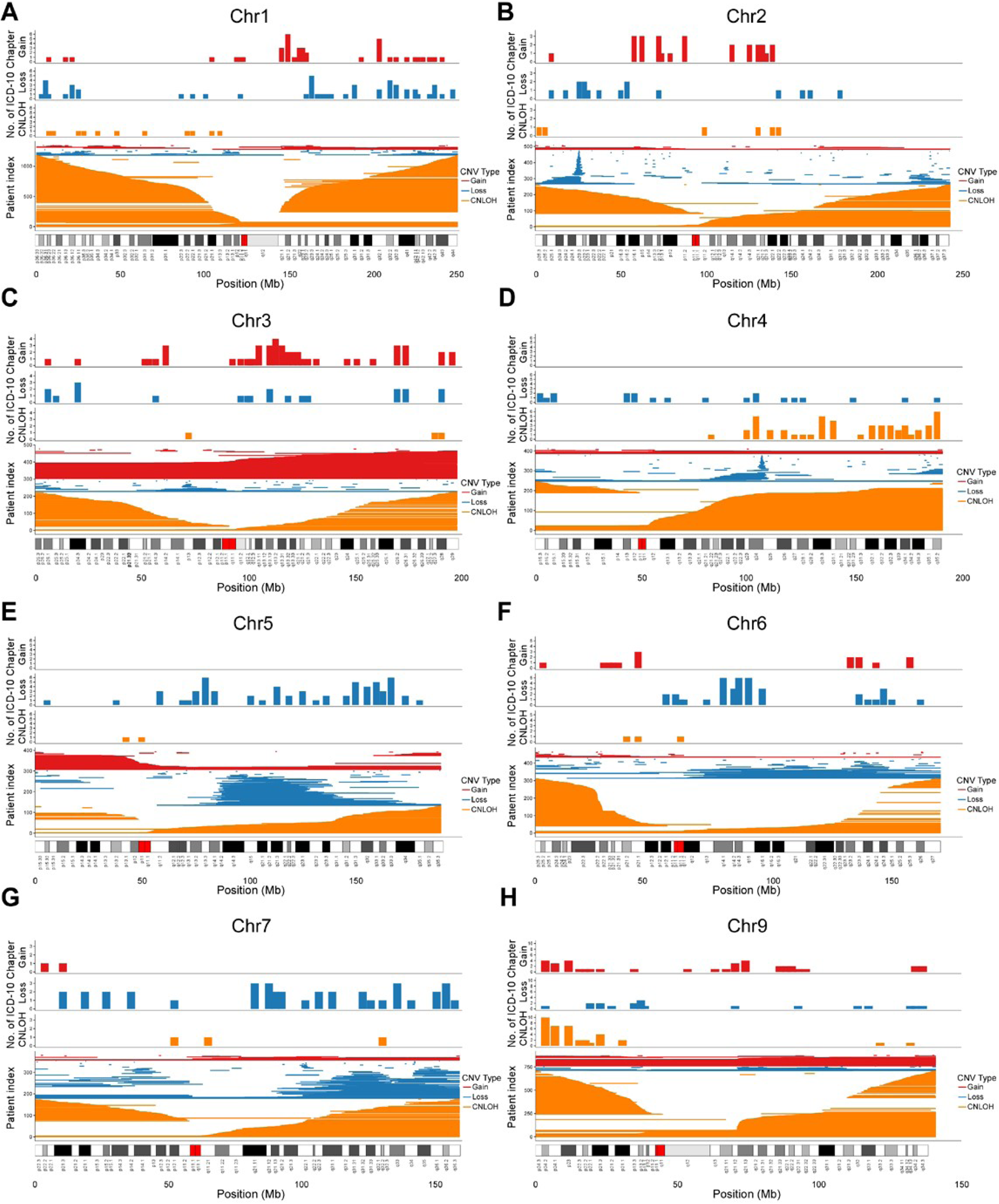
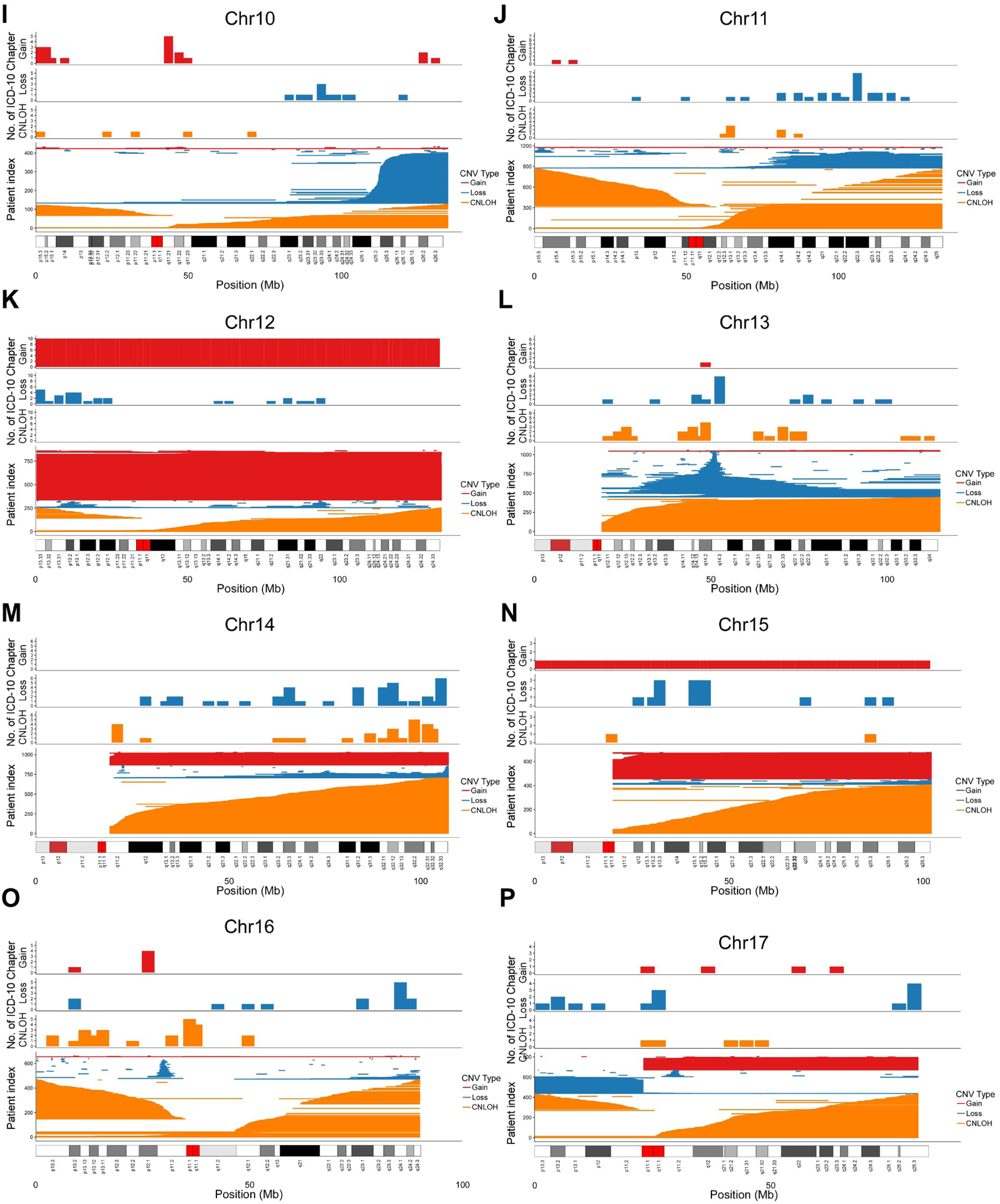
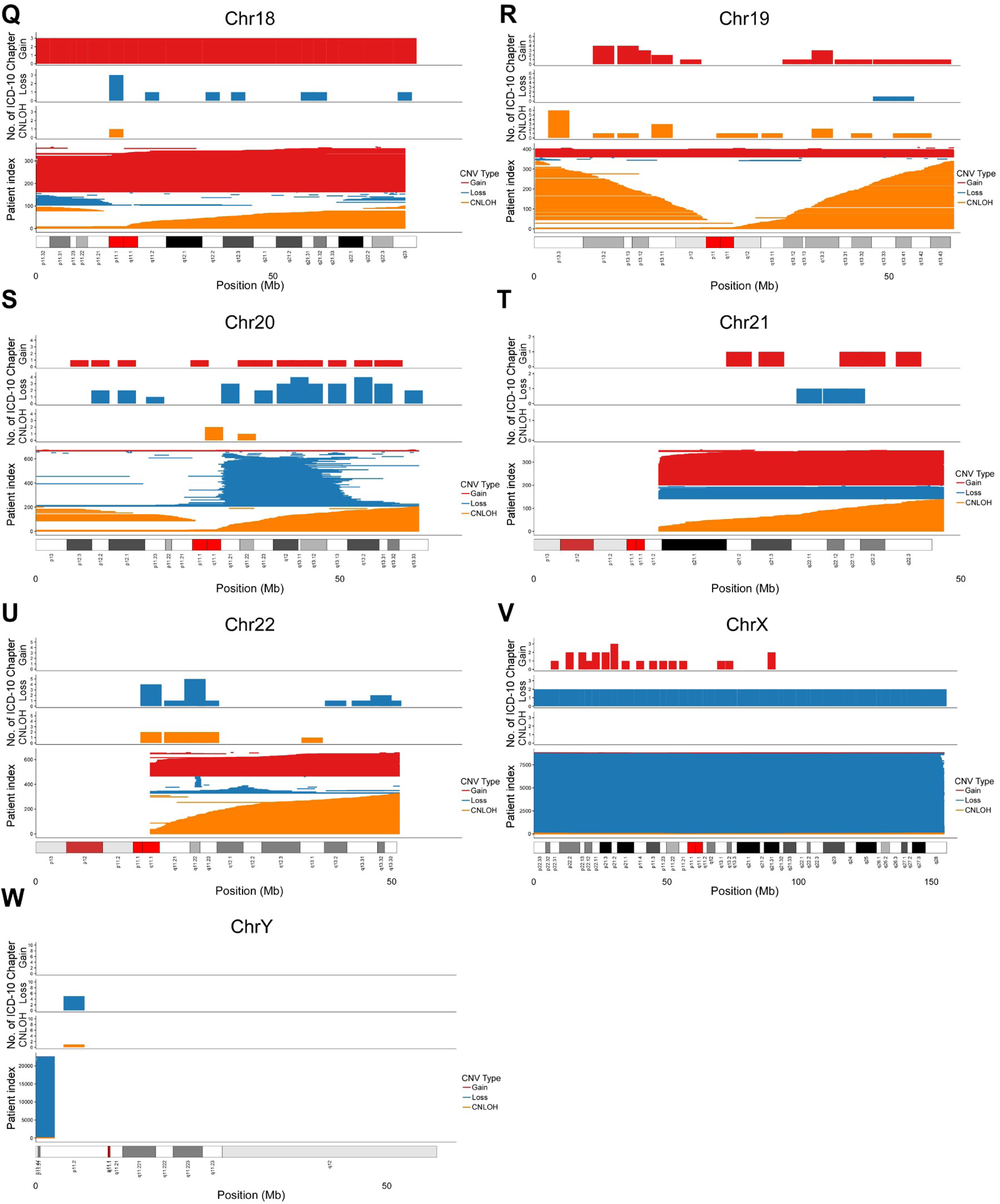
Cytoband-level disease associations and mCA distribution across all chromosomes. **a-w**, Chromosome-level summary plots for chr1-7 and chr9-22 (**a-u**; chr8 data are shown in Figure 6a), chrX (**v**), and chrY (**w**). For each chromosome, top tracks display the number of ICD-10 disease chapters with significant cytoband-level associations (adjusted q<0.05) for gain (red), loss (blue), and CNLOH (orange). Bottom tracks show mCA distribution across the chromosome; each horizontal line represents an individual mCA event, with colors indicating CNV type (red, gain; blue, loss; orange, CNLOH). Chromosomal ideograms with cytoband annotations are shown at the bottom of each panel. For heterogeneous mCAs, cytobands were selected using LASSO-penalized Cox regression and refitted using standard Cox proportional hazards models adjusted for age, sex, and baseline disease burden. For homogeneous mCAs (12+, 14+, 15+, 18+, X-), which preclude cytoband-level resolution, standard Cox proportional hazards regression was performed comparing carriers against participants with no detectable mCA, adjusting for age, sex, and baseline disease burden, and the resulting chromosome-wide hazard ratio was applied uniformly across all cytobands of the affected chromosome. Multiple testing was corrected using the Benjamini-Hochberg method.

**Supplementary Fig. 12.**
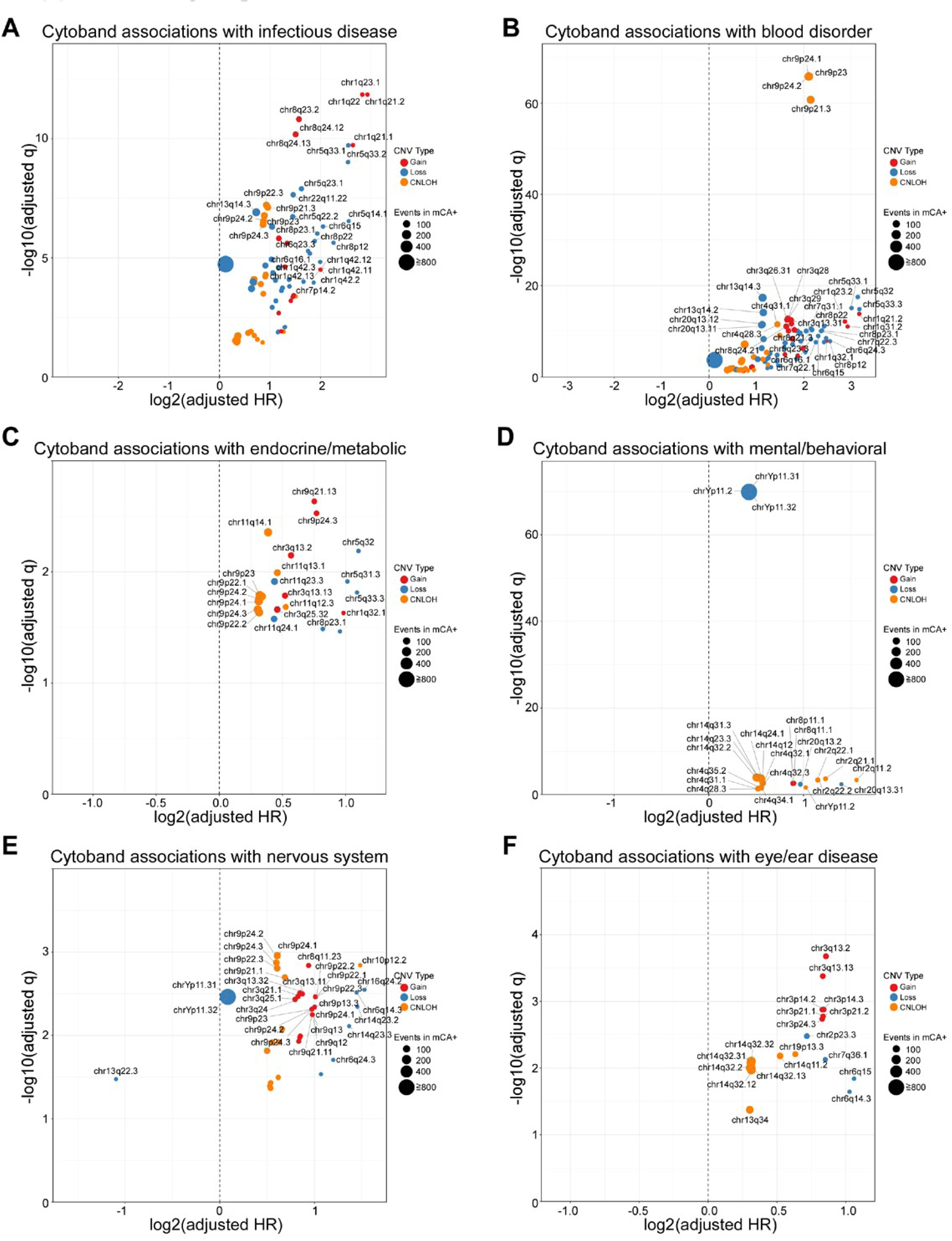
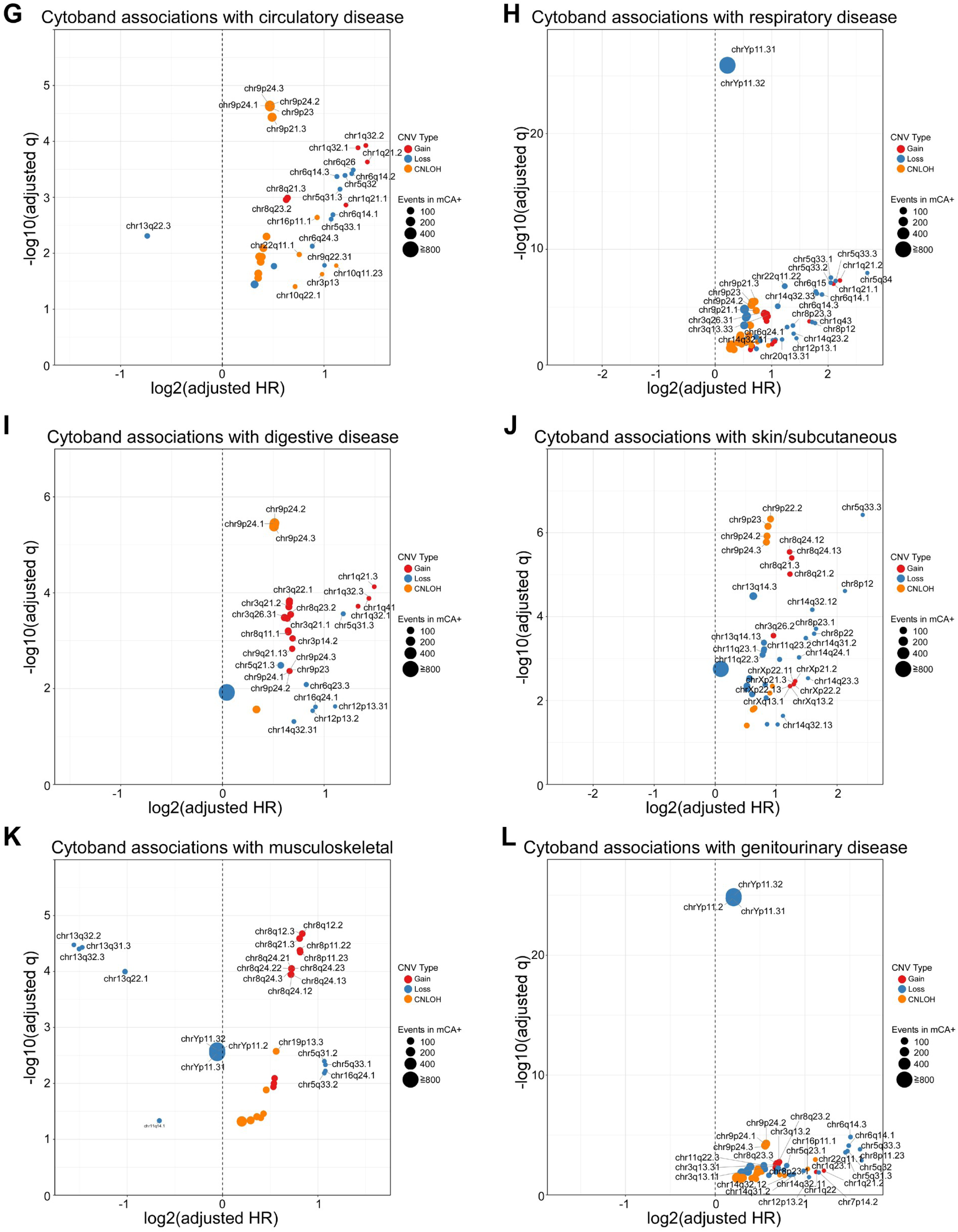
Cytoband-level disease associations by ICD-10 chapter. **a-l**, Volcano plots showing cytoband-level associations with incident disease risk across ICD-10 chapters: infectious diseases (**a**), hematologic disorders (**b**), endocrine/metabolic (**c**), mental/behavioral (**d**), nervous system (**e**), eye and ear (**f**), circulatory (**g**), respiratory (**h**), digestive (**i**), skin/subcutaneous (**j**), musculoskeletal (**k**), and genitourinary (**l**). The x-axis represents log₂-transformed HRs and the y-axis represents −log₁₀(FDR-adjusted q-values). Dot size reflects number of incident disease events among mCA carriers; colors indicate CNV type (red, gain; blue, loss; orange, CNLOH). Dashed vertical line indicates log₂(HR)=0. Cytobands were selected using LASSO-penalized Cox regression and refitted using standard Cox proportional hazards models adjusted for age, sex, and baseline disease burden. Cytoband-level HRs were estimated as described in Methods. Multiple testing was corrected using the Benjamini-Hochberg method; only cytobands with adjusted q<0.05 are displayed.

**Supplementary Fig. 13.**
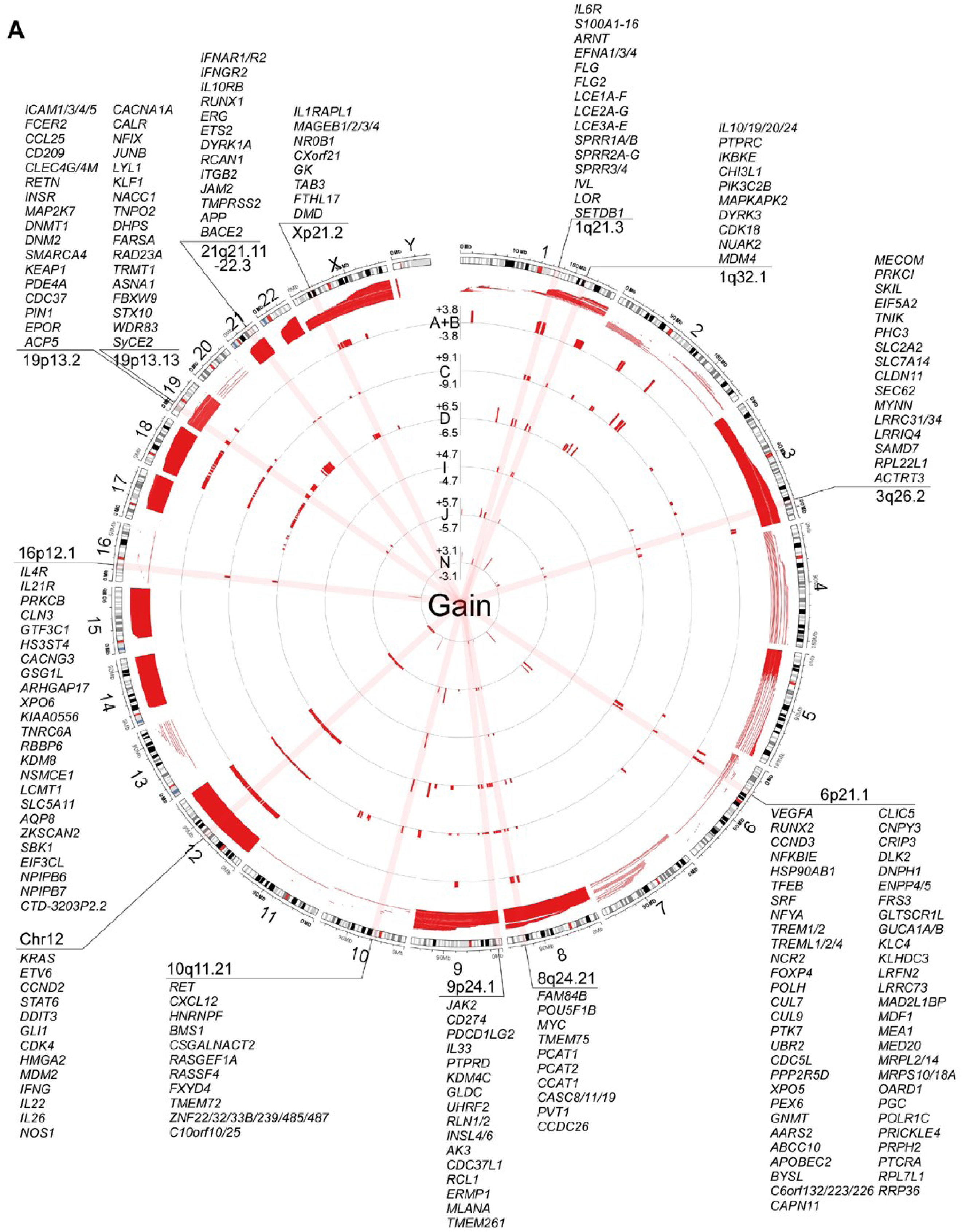
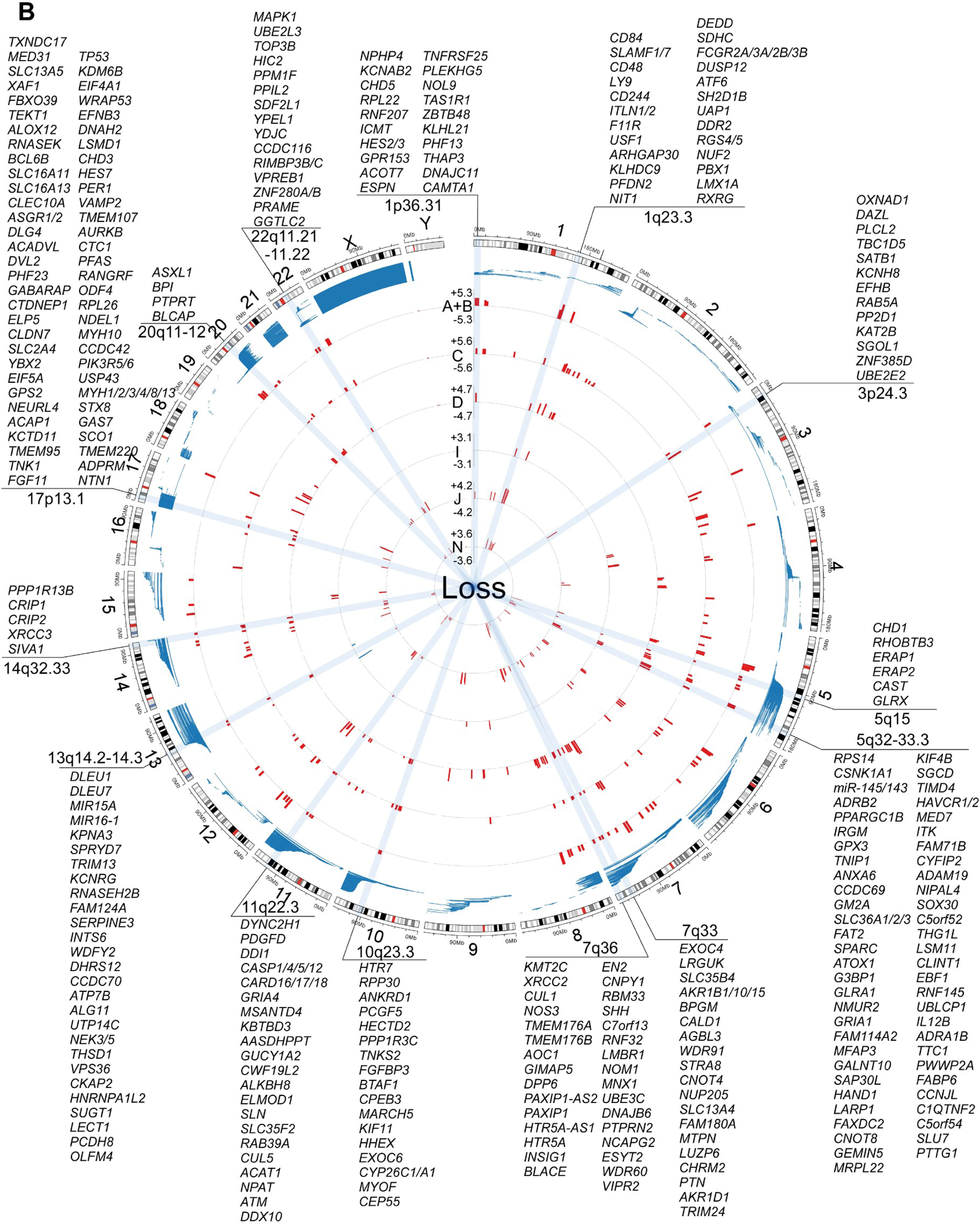
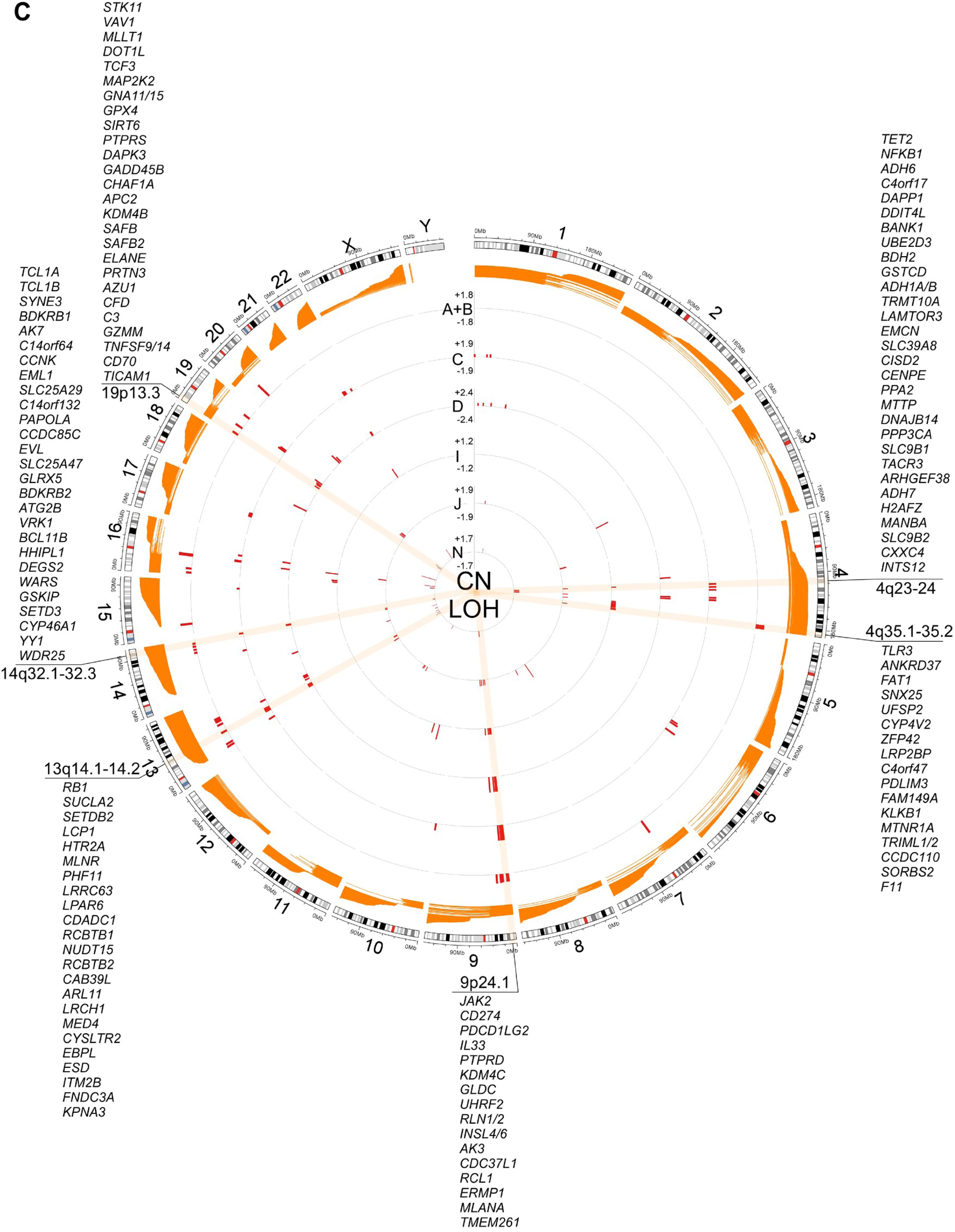
Genome-wide circos plots of disease-associated cytobands by CNV type. **a-c**, Circos plots showing cytobands significantly associated with incident disease risk (adjusted q<0.05) for gain (**a**), loss (**b**), and CNLOH (**c**). Outer track displays chromosomal ideogram. Inner tracks show log₂-transformed HRs for six selected ICD-10 disease chapters: infectious diseases (A+B), neoplasms (C), hematologic disorders (D), circulatory (I), respiratory (J), and genitourinary (N). Red bars indicate increased risk (HR>1); blue bars indicate decreased risk (HR<1). Cytobands were selected using LASSO-penalized Cox regression and refitted using standard Cox proportional hazards models adjusted for age, sex, and baseline disease burden. Multiple testing was corrected using the Benjamini-Hochberg method. Candidate genes within disease-associated cytobands (adjusted q<0.05) were retrieved from Ensembl GRCh37 using biomaRt. Protein-coding genes and microRNAs overlapping with significant cytoband coordinates are annotated.

**Supplementary Fig. 14.**
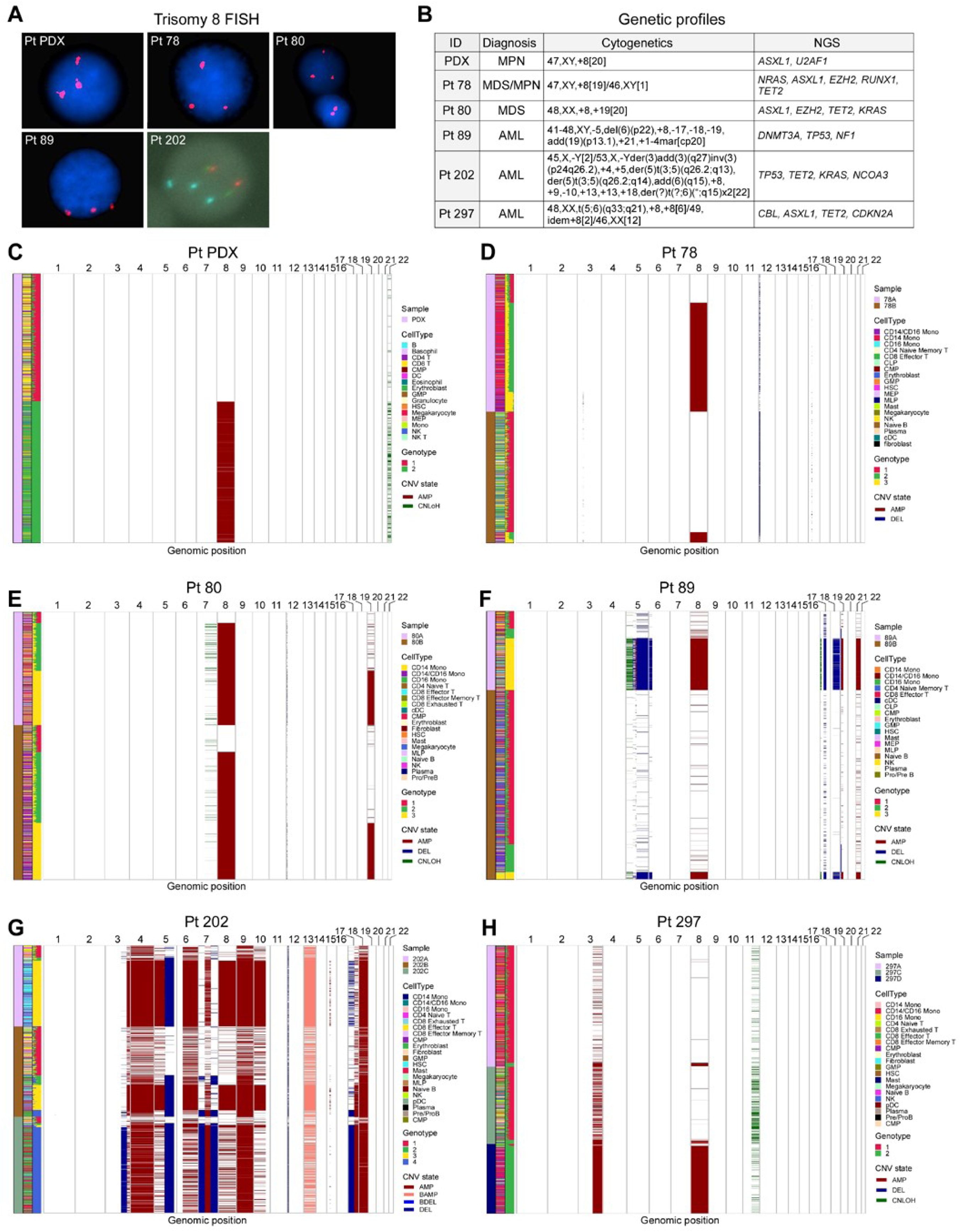
Single-cell-resolved chromosome copy number analysis in bone marrow mononuclear cells from patients with myeloid diseases. **a,** Representative fluorescence in situ hybridization (FISH) images confirming trisomy 8 in BM cells from patients with myeloid neoplasms. Chr8 centromeric probes appear as red signals (Pt PDX, Pt 78, Pt 80, Pt 89) or aqua signals (Pt 202). Three discrete signals indicate trisomy 8. Nuclei are counterstained with DAPI (blue). **b**, Clinical and genetic profiles of patients with chr8 gain. Cytogenetics were assessed by conventional karyotyping and somatic mutations were identified by a 98-gene myeloid NGS panel. **c-h**, Numbat analysis of scRNA-seq data revealing CNVs in BM cells from each patient (see Methods). Columns represent chromosome locations (p arm on the left, q arm on the right) and rows represent individual cells. CNV states are shown as gain (red), bi-allelic gain (pink), loss (dark blue), bi-allelic loss (blue), and CNLOH (green). Left annotations indicate sample source (A, pre-treatment; B, post-treatment; C/D, relapse), cell type, and genotype cluster; distinct CNV groups were identified based on shared CNV patterns across cells.

**Supplementary Fig. 15.**
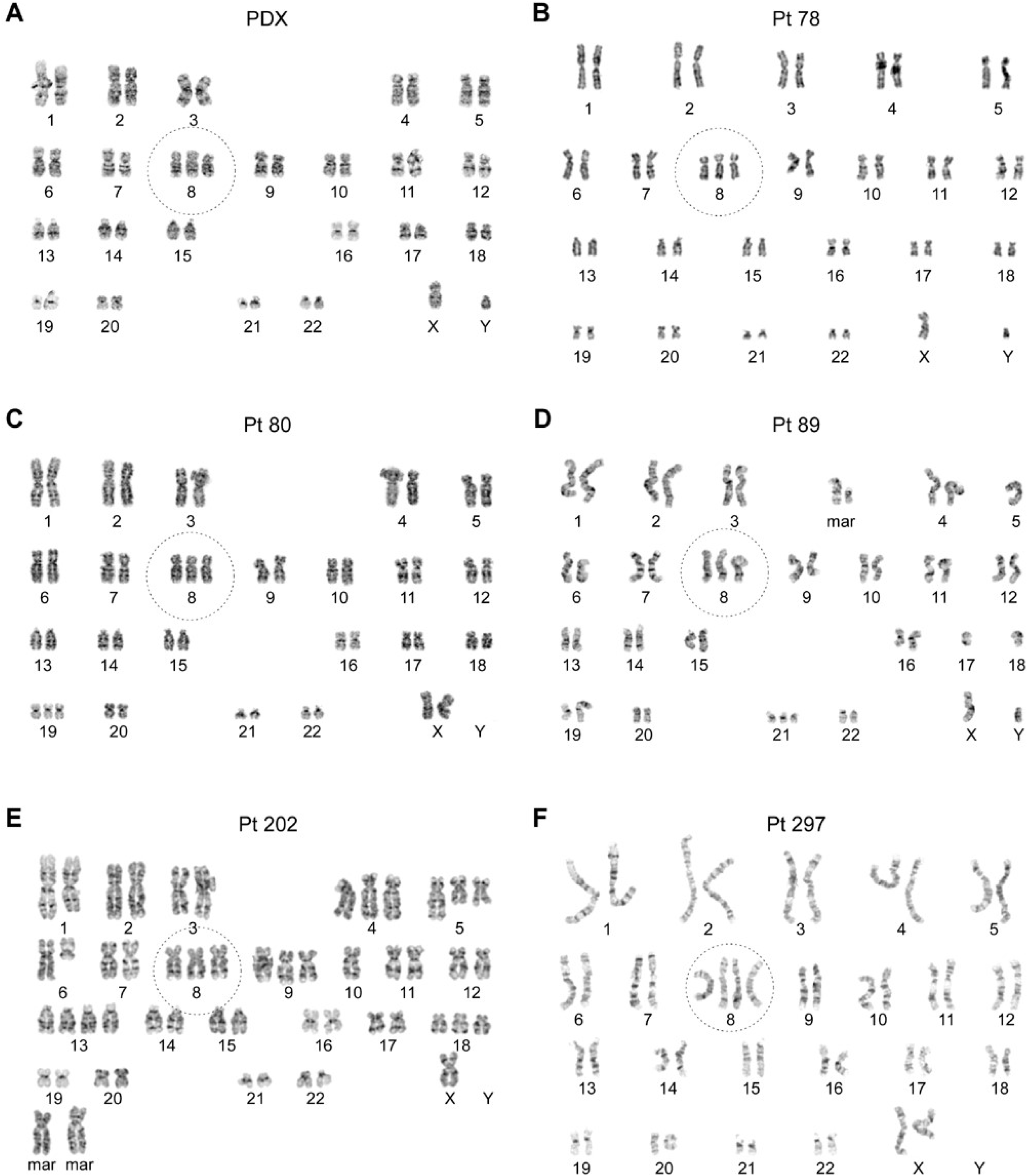
Cytogenetic validation of trisomy 8 in patients with myeloid diseases. **a-f**, G-banding (trypsin-Giemsa) karyotype analysis was performed by Laboratory Corporation of America (Burlington, NC, USA). Nomenclature follows the International System for Human Cytogenomic Nomenclature (ISCN 2016). Each panel displays the diagnostic karyotype from an independent patient included in the scRNA-seq analysis shown in Fig. 6 and **Supplementary Fig. 14**. Chr8 is highlighted with a dashed circle in each panel.

**Supplementary Fig. 16.**
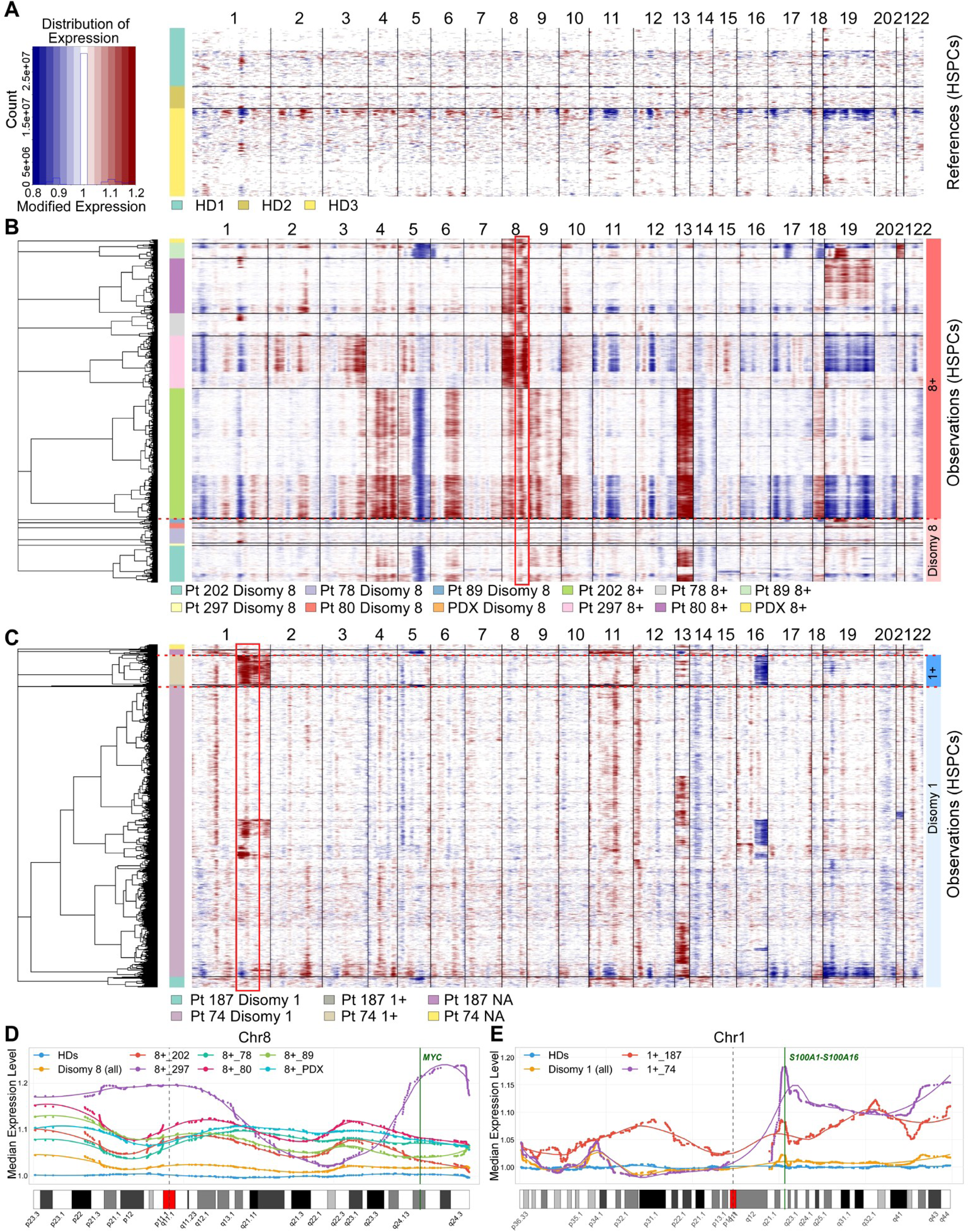
InferCNV analysis reveals focal transcriptional activation in aneuploid HSPCs. **a**, Genome-wide expression heatmap from InferCNV analysis of HSCs, CMPs, and GMPs in healthy donors (HD1, HD2, and HD3) serving as the reference population. Columns represent chromosomal positions (chromosomes 1–22), and rows represent individual cells. Color scale indicates modified expression values relative to the reference (blue, decreased expression; white, neutral; red, increased expression). **b**, InferCNV heatmap comparing cells with disomy 8 vs. cells with 8+ across multiple patients (Pt 78, Pt 80, Pt 89, Pt 202, Pt 297, and PDX). Hierarchical clustering groups cells by expression similarity. The prominent transcriptional upregulation (red box) is observed specifically on chr8 in 8+ cells but not in disomy 8 cells, with focal enrichment at the 8q21.3-24.23 region encompassing the *MYC* locus. **c**, InferCNV heatmap comparing cells with disomy 1 vs. cells with 1+ in two patients (Pt 74 with 1q gain and Pt 187 with whole chr1 gain). NA indicates cells with undetermined copy number status. Transcriptional activation is focally enriched within the 1q21.1–23.2 region (red box), which contains the *S100A1-S100A16* gene cluster, including *S100A9*. **d-e**, Quantitative analysis of chr8 (**d**) and chr1 (**e**) gene expression in HSCs across genomic positions. Smoothed LOESS curves show mean InferCNV-adjusted expression levels for healthy donors (HD, blue), disomy cells (orange), and individual samples with 8+ or 1+ (colored lines). Each dot represents a single gene’s mean expression. Green vertical lines mark key disease-associated loci: *MYC* at 8q24.21 and *S100A9* at 1q21.3. Gray dashed lines indicate centromere positions. Bottom panels show chromosome ideograms with cytoband annotations. 8+ cells demonstrate elevated expression across chr8, particularly in the 8q21.3–24.23 region, and 1+ cells show elevated expression predominantly on 1q, particularly in the 1q21.1–23.2 region, consistent with the focal amplification patterns observed in panels **b** and **c**. For panels **b-e**, copy number status was determined using Numbat, and InferCNV was used for visualization of transcriptional effects.

**Supplementary Fig. 17.**
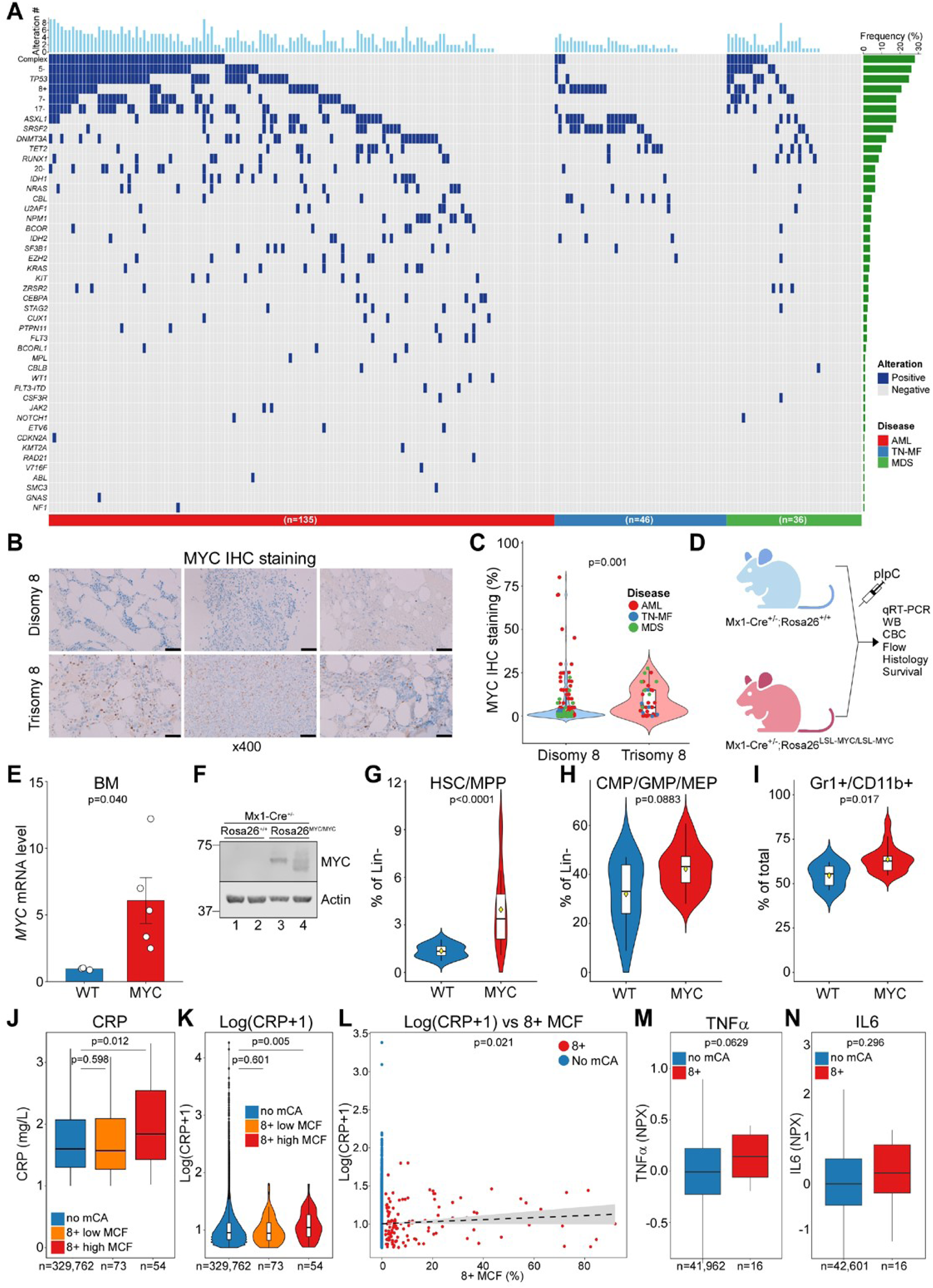
Genomic landscape of myeloid diseases with trisomy 8 and functional interrogation of MYC overexpression. **a**, Oncoplot showing somatic mutations and chromosomal alterations in patients with myeloid diseases. Columns represent individual patients; rows represent genes or chromosomal alterations. Top bar shows number of alterations per patient. Right bar shows mutation or mCA frequency (%). Patients are grouped by disease type: acute myeloid leukemia (AML; n=135), triple-negative myelofibrosis (TN-MF; n=46), and myelodysplastic syndrome (MDS; n=36). Blue indicates presence of alteration. **b**, Representative MYC immunohistochemistry (IHC) staining in BM samples from patients with disomy 8 vs. trisomy 8 across disease types (scale bar, 50 µm). **c**, Quantification of MYC IHC staining comparing disomy 8 (n=153) vs. trisomy 8 (n=39) among patients with available IHC data (n=192/217); P value calculated using Wilcoxon rank-sum test. **d**, Schematic of the Mx1-Cre^+/-^;Rosa26^LSL-MYC/LSL-MYC^ mouse model for conditional *MYC* overexpression following pIpC induction. **e-f**, Confirmation of MYC overexpression by qRT-PCR (**e**) and immunoblotting (**f**). **g-i**, Flow cytometric analysis of BM populations in WT vs. MYC mice: HSC/MPP (% of Lin-) (**g**), CMP/GMP/MEP (% of Lin-) (**h**), and Gr1+/CD11b+ myeloid cells (% of total) (**i**). Violin plots with embedded box plots show median and interquartile range; P values were calculated using Wilcoxon rank-sum test. **j**, Box plots showing raw CRP levels (mg/L) across three groups: no mCA (n=329,762), 8+ with MCF <10% (n=73), and 8+ with MCF ≥10% (n=54). Boxes represent the interquartile range (IQR, 25^th^-75^th^ percentiles), with the horizontal line indicating the median. Whiskers extend to 1.5×IQR. Outliers beyond the 95^th^ percentile are not displayed for visualization purposes. **k,** Violin plots showing the distribution of log-transformed CRP [log(CRP+1)] for the same groups. The width of each violin represents the kernel density estimate of the data distribution, with embedded box plots displaying the median and IQR. In **j** and **k**, group comparisons were performed using the Wilcoxon rank-sum test for raw CRP values and Welch’s t-test for log-transformed CRP. Individuals with 8+ and MCF ≥10% had significantly elevated CRP compared to no-mCA controls [median 1.84 (IQR 1.43-2.54) vs. 1.60 (IQR 1.30-2.07) mg/L; Wilcoxon p=0.012]. In contrast, 8+ with MCF <10% showed no significant difference in CRP compared to controls [median 1.57 (IQR 1.27-2.09) mg/L; Wilcoxon p=0.598]. **l,** Scatter plot showing the relationship between 8+ MCF and log-transformed CRP [log(CRP+1)]. Individuals without mCA are shown at MCF=0 (blue; n=5,000 randomly sampled for visualization), and 8+ carriers are shown at their actual MCF values (red; n=127). The dashed line represents the linear regression fit with 95% CI (shaded area). Multivariable linear regression was used to assess the association between MCF and log-transformed CRP, adjusted for age and sex. A significant positive association was observed (β=0.164, 95% CI: 0.024-0.304, p=0.021), indicating that higher 8+ clone burden is associated with increased levels of systemic inflammation. **m-n**, Box plots showing circulating TNFα (**m**) and IL6 (**n**) protein levels measured by Olink proteomics in no mCA controls (n=41,962 for TNFα; n=42,601 for IL6) vs. 8+ carriers (n=16). Values are expressed as Normalized Protein eXpression (NPX, log2 scale). Boxes represent IQR, horizontal lines indicate median, and whiskers extend to 1.5×IQR. P values were calculated using Wilcoxon rank-sum test.

**Supplementary Fig. 18.**
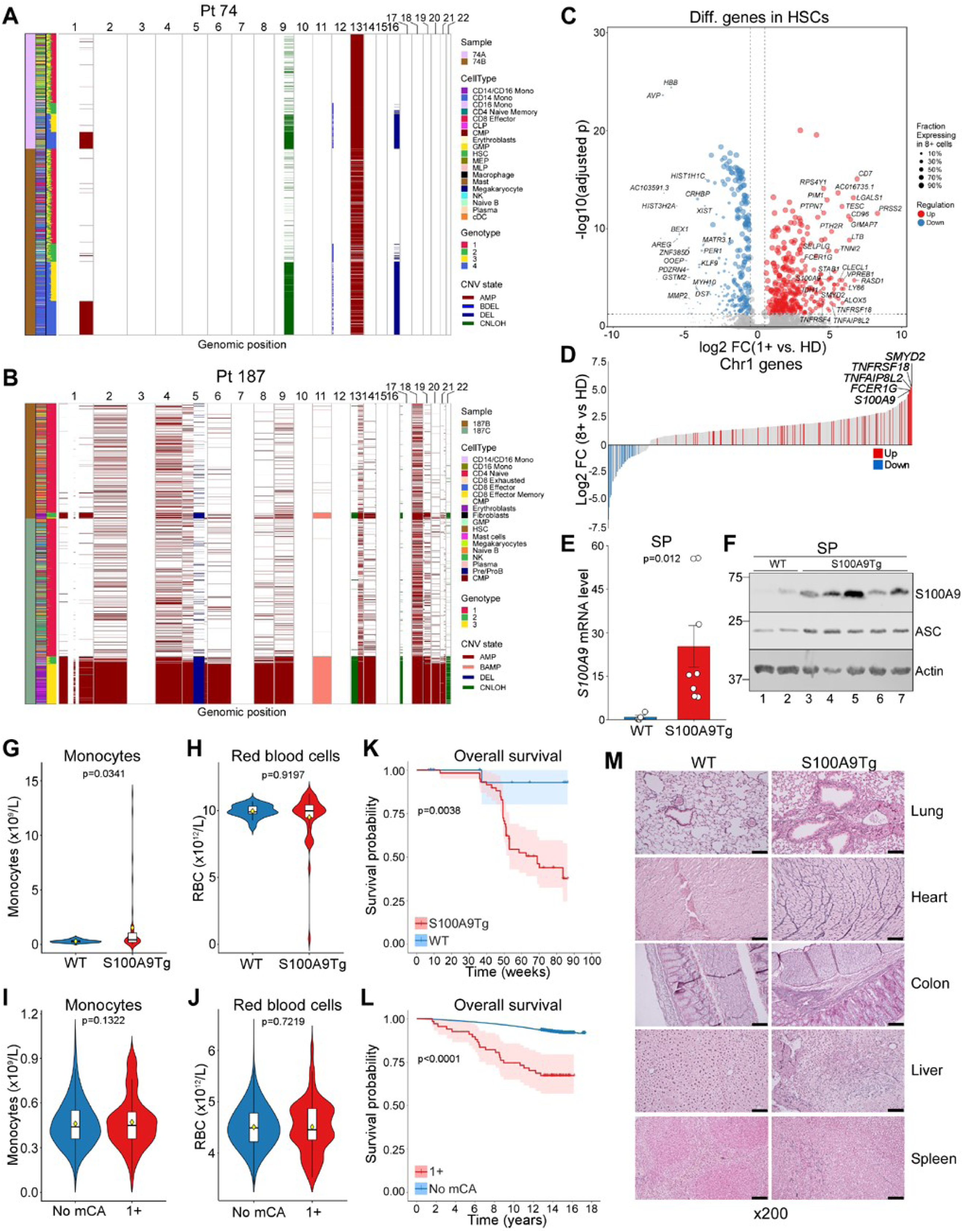
Functional validation of *S100A9* as a candidate gene in trisomy 1. **a-b**, Numbat analysis of scRNA-seq data revealing CNVs in BM cells from patients with 1+: Pt 74 (**a**) and Pt 187 (**b**). Columns represent chromosome locations (p arm on the left, q arm on the right) and rows represent individual cells. CNV states are shown as gain (red), biallelic gain (pink), loss (blue), and CNLOH (green). Left annotations indicate sample source (first column), cell type (cluster annotation; second column), and genotype cluster (third column). **c**, Differentially expressed genes in HSCs from patients with 1+ compared to healthy donors (HD). scRNA-seq was performed on 25,932 cells (1+, n=2,372; disomy 1, n=23,560; classified using Numbat, see Methods) from 4 samples (2 patients), and 16,504 cells from 3 HD. Dot size reflects fraction of 1+ cells expressing each gene; colors indicate upregulation (red) or downregulation (blue). **d**, Chr1 genes ranked by log₂ fold change in 1+ HSCs vs. HD. **e-f**, Confirmation of S100A9 overexpression by qRT-PCR (**e**) and immunoblotting (**f**) in spleen (SP) from S100A9 transgenic (S100A9Tg) vs. wild type (WT) mice. S100A9 expression levels in S100A9Tg mice are comparable to those observed in 1+ human HSPCs (panel **d**). **g-h**, Peripheral blood monocyte counts (**g**) and RBC counts (**h**) in S100A9Tg mice vs. WT controls. **i-j**, Monocyte counts (**i**) and RBC counts (**j**) in UKBB participants with 1+ vs. non-carriers. **k-l**, Overall survival in S100A9Tg (n=59) vs. WT (n=25) mice (**k**) and in UKBB participants with 1+ (n=67) vs. non-carriers (n=410,204) (**l**). **m**, Histological analysis of fibrosis using reticulin staining in lung, heart, colon, liver, and spleen from S100A9Tg vs. WT mice; original magnification x200; scale bar, 50µm. For **g-j**, violin plots with embedded box plots show median and interquartile range. For **e** and **g-j**, P values were calculated using the Wilcoxon rank-sum test. For **k** and **l**, shaded areas represent 95% CIs; P values were calculated using the log-rank test.

**Supplementary Fig. 19.**
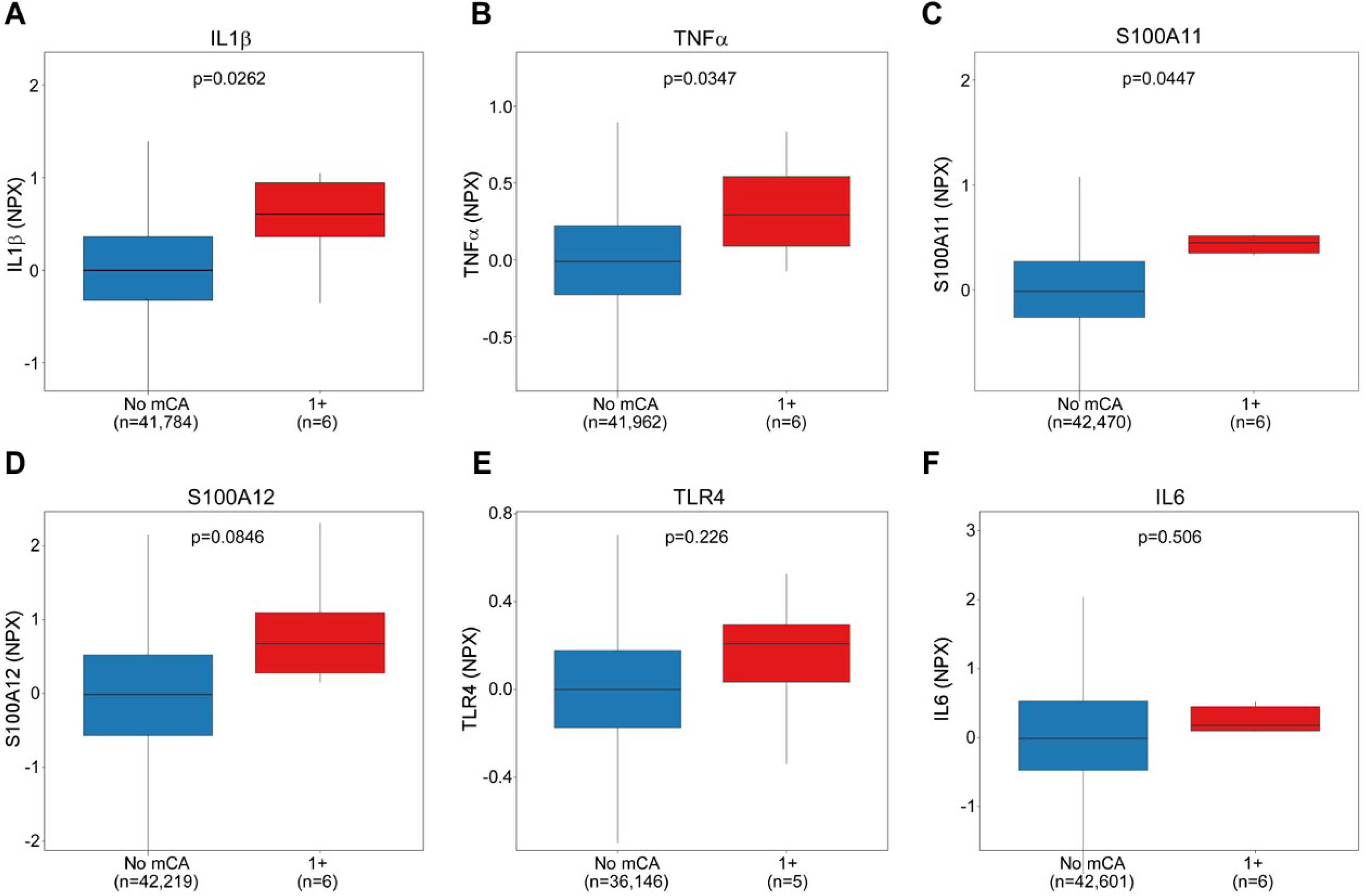
Levels of pro-inflammatory cytokines and alarmins in 1+ carriers. **a-f**, Box plots showing circulating protein levels measured by Olink proteomics in no mCA controls vs. 1+ carriers. Sample sizes for 1+ carriers: n=6 for all cytokines and alarmins except TLR4. Values are expressed as Normalized Protein eXpression (NPX, log2 scale). Boxes represent the IQR, horizontal lines indicate the median, and whiskers extend to 1.5×IQR. P values were calculated using the Wilcoxon rank-sum test. IL1β (p=0.0262) (**a**), TNFα (p=0.0347) (**b**), and S100A11 (p=0.0447) (**c**) showed significantly elevated levels in 1+ carriers, while S100A12 (p=0.0846) (d), TLR4 (p=0.226) (e), and IL6 (p=0.506) (f) were numerically higher but did not reach statistical significance. Note that the *S100A11* and *S100A12* genes are located on chromosome 1q21.3, which is associated with increased risk of multimorbidity (see Supplementary Fig. 11A and Supplementary Fig. 12A).

